# Elucidating the Epigenetic Landscape of Type 2 Diabetes: A Multi-Omics Analysis Revealing Novel CpG Sites and Their Association with Cardiometabolic Traits

**DOI:** 10.1101/2024.05.20.24307650

**Authors:** Ren-Hua Chung, Chun-Chao Wang, Djeane Debora Onthoni, Ben-Yang Liao, Tzu-Sheng Hsu, Eden R. Martin, Chao A. Hsiung, Wayne Huey-Herng Sheu, Hung-Yi Chiou

## Abstract

Type 2 Diabetes (T2D) is a complex, multifactorial disease with a significant global health burden. Genome-Wide Association Studies (GWAS) have identified numerous genetic variants associated with T2D, yet understanding their functional mechanisms remains challenging due to the polygenic nature of the disease and the prevalence of non-coding region variants. This study leverages a multi-omics approach integrating methylome-wide association studies (MWAS), Mendelian Randomization (MR), and functional analyses in human pancreatic cells and mouse models to elucidate the functional consequences of genetic variants on T2D. Using summary statistics calculated from large-scale GWAS for T2D and a DNA methylation (DNAm) prediction model, our MWAS tested the association of DNAm at CpGs in whole blood with T2D across the genome. We identified 87 significant and independent CpG sites associated with T2D risk in Europeans, including 13 novel CpG sites located in or near genes not previously associated with T2D, with these results being replicated in an additional dataset. Our analyses also revealed a significant overlap of these CpGs with cardiometabolic traits, underscoring the interconnectedness of metabolic diseases. Additionally, we demonstrated trans-ethnic effects of significant CpGs in East Asians, highlighting their global relevance. Functional analyses in human pancreatic alpha and beta cells identified potential regulatory roles of these CpGs in gene expression, particularly in genes involved in glucose metabolism. Notably, differential expression of the PPP1R3B gene, regulated by a significant CpG, between T2D cases and normal controls was observed in both alpha and beta cells, with mouse models confirming its role in glucose homeostasis. Our findings provide new insights into the epigenetic architecture of T2D, revealing novel genetic susceptibilities and highlighting potential targets for therapeutic intervention.

## Introduction

The study of genetics plays a pivotal role in unraveling the complex etiology of Type 2 Diabetes (T2D), a multifactorial disease with a significant global health impact. While genome-wide association studies (GWAS) have successfully identified numerous genetic variants associated with T2D ^1^, translating these findings into a clear understanding of the functional mechanisms impacting disease remains a significant challenge. This difficulty stems primarily from the complexity of distinguishing direct causal variants from those in linkage disequilibrium, and the polygenic nature of T2D where multiple genes and their interactions with environmental factors affect disease risk. Furthermore, most GWAS hits are located in non-coding regions of the genome, complicating the task of determining how these variants influence gene expression and contribute to T2D pathogenesis ^2^. Addressing this challenge necessitates integrated approaches that combine GWAS data with functional genomics and other omics technologies, aiming to elucidate the biological pathways impacted by these genetic variations ^3^.

A promising approach to overcome this challenge involves the imputation of gene expression levels using expression quantitative trait loci (eQTL) derived from GWAS. eQTL are genomic loci that explain variation in gene expression levels, which serve as powerful tools for interpreting the functional consequences of genetic variants identified in GWAS ^4^. Building upon this, the concept of transcriptome-wide association studies (TWAS) leverages genotype data and eQTL information to impute gene expression levels in individuals, linking these imputed expression levels to phenotypic traits ^5^. Recent advancements have demonstrated the utility of TWAS in uncovering novel gene-disease associations, with studies applying TWAS to identify genes implicated in complex diseases such as schizophrenia, T2D, and coronary artery disease, thereby offering new insights into disease etiology and highlighting potential therapeutic targets ^6,7^.

The principles underlying TWAS are effectively applicable to the study of DNA methylation (DNAm), a pivotal mechanism for regulation of gene expression. Specifically, CpG methylation, which entails the addition of a methyl group to cytosine residues within cytosine-guanine dinucleotides, influences gene activity, often leading to decreased expression when occurring in promoter regions, despite the complexity of these interactions ^8^. Moreover, abnormal DNAm levels are associated with various complex diseases, identified through epigenome-wide association studies (EWAS) ^9^. Importantly, DNAm is not just an epigenetic phenomenon but also a heritable one. Studies have demonstrated that CpG methylation possesses a significant heritable component, with heritability estimates ranging from 16% to 20% ^10^. Consequently, many methylation quantitative trait loci (meQTL), which are genetic variants affecting the DNAm levels, have been identified ^11^. These two key aspects ─ the heritability of DNAm and the identification of meQTL across CpGs ─ pave the way for conducting methylome-wide association studies (MWAS), which test the association between the imputed DNAm levels based on meQTL from GWAS and diseases.

Conducting a MWAS typically requires GWAS data and a DNAm prediction model, similar to TWAS. The pioneering work in this area was conducted by Fryett et al. ^10^, who constructed a methylation prediction model using data from the Understanding Society: UK Household Longitudinal Study ^12^. When this model was applied to a study of primary biliary cholangitis, more than 700 CpGs achieving a genome-wide significance level were identified. Despite these promising results, Fryett et al.’s research has not gained significant traction in the scientific community, and similar studies focusing on East Asian populations are notably absent.

In this study, we conducted a MWAS in Europeans to identify significant CpGs where DNAm levels are regulated by meQTL for T2D. The significant results were replicated using Mendelian Randomization (MR) in an independent dataset and validated with samples having measured methylation levels. We also evaluated the associations of the identified CpGs with other cardiometabolic traits using MWAS, and their trans-ethnic effects in East Asians.

Finally, methylation and gene-expression data in human pancreatic alpha and beta cells were used to evaluate the potential regulatory roles of the CpGs in gene expression, following by the functional analysis in mouse models.

## Methods

### Methylome-wide association analysis (MWAS) for type 2 diabetes

TWAS harness the relationship between eQTL and gene expression to predict gene expression levels and their correlation with phenotypes. PrediXcan ^13^, a tool widely adopted within the TWAS framework when raw SNP genotypes are available, divides an individual’s gene expression into three components: genetically regulated expression (GReX), reflecting inherent genetic contributions; alterations by traits, such as disease-modified expression levels; and environmental factors. PrediXcan’s primary objective is to assess GReX’s impact on specific traits or phenotypes, utilizing additive models trained on reference transcriptome datasets from resources like GTEx ^14^ to estimate genetically regulated expressions. These models enable the prediction of gene expression levels from multiple SNPs, which are then associated with actual phenotypic data. S-PrediXcan advances this methodology by inferring PrediXcan outcomes using only GWAS summary statistics, thus bypassing the need for individual-level data. Similarly, this paradigm extends to DNA methylation studies, where Fryett et al. ^10^ developed a methylation prediction model indicating the feasibility of predicting methylation levels using SNPs. This model, trained on a dataset of 1,120 samples, achieved notable prediction accuracy across 179,523 CpG sites. The PrediXcan and S-PrediXcan framework can be directly applied to perform an MWAS, adopting the prediction model for CpG methylation levels based on GWAS data. More details of the PrediXcan and S-PrediXcan algorithms as well as the DNAm prediction model are provided in the Supplementary Materials.

For our study, the Diabetes Meta-Analysis of Trans-Ethnic association studies (DIAMANTE) consortium provided GWAS summary statistics for T2D in Europeans calculated based on 80,154 T2D cases and 853,816 controls. Using the methylation prediction model constructed by Fryett et al. ^10^, along with the DIAMANTE GWAS summary statistics, we conducted an MWAS through S-PrediXcan. This involved association testing for imputed methylation levels at 179,523 CpGs. CpGs with association p-values < 2.78 × 10^−7^— a threshold set to consider multiple testing across 179,523 CpGs—were selected for further replication analysis. This process identified a total of 1,120 significant CpGs.

### Mendelian randomization analysis

TWAS methods such as PrediXcan and S-PrediXcan perform tests based on a two-stage regression-based framework. Initially, gene expression levels are imputed based on eQTL in a prediction model. Subsequently, the association between these imputed gene expression levels and the outcome of interest is tested. This approach mirrors the two-sample MR framework, suggesting that significant signals identified via S-PrediXcan might indicate potential MR effects ^15^. However, TWAS methods such as S-PrediXcan cannot distinguish causal effects from horizontal pleiotropy, where a genetic variant affects multiple phenotypes through different biological pathways ^16^. To address this limitation and validate the causality of our findings, we leveraged methods developed specifically for MR in our replication analysis. Ensuring the robustness of our results, we used distinct datasets for both GWAS data on T2D and meQTL information, which were independent from those used in the MWAS, in our MR analysis. This approach allowed us to replicate the MWAS findings and further evaluate the causal effects of CpGs on T2D.

A two-sample MR analysis was carried out using the inverse-variance weighted (IVW) ^17^ algorithm, the most widely used method in MR, alongside the MR-Egger ^18^ intercept test to evaluate the potential effect of pleiotropy. Both IVW and MR-Egger tests were conducted using the R package MendelianRandomization ^19^. To select instrumental variables for the significant CpGs, we searched the MeQTL EPIC database ^11^, which contains meQTL data for 724,499 CpGs profiled on the Illumina Infinium MethylationEPIC array in 2,358 blood samples from three UK cohorts. We chose clumped meQTL with low linkage disequilibrium for independence and applied a false discovery rate (FDR) < 0.05 to exclude weak instrumental variables, ensuring robust MR analysis.

Association statistics of the instrumental variables with T2D were obtained from the FinnGen study ^20^, another large-scale GWAS for T2D in Europeans, consisting of 38,657 T2D cases and 310,131 controls. In adhering to the key assumptions of two-sample MR, we ensured that the summary statistics for the Exposure-SNP (i.e., meQTL information) and Outcome-SNP (i.e., T2D associations) were derived from independent but related populations. This approach aligns with the principle of using non-overlapping datasets for the exposure and outcome in two-sample MR to avoid bias. We required a minimum of 3 instrumental meQTL per CpG to enable pleiotropy tests, resulting in 495 out of the 1,120 CpGs qualifying for MR analysis. Of these, 153 CpGs were successfully replicated, showing significant MR p-values < 1.01 × 10^−4^ — considering the 495 tests conducted — without evidence of pleiotropy effects (MR-Egger p-values > 0.05).

### Joint modeling of replicated CpGs

We assessed the independent and joint effects of the 153 replicated CpGs on T2D within the UK Biobank, which includes 25,025 prevalent T2D cases and 314,430 controls. Imputation of CpG levels for these 153 CpGs was performed using PrediXcan. Further information on QC procedures and the criteria for defining T2D in the UK Biobank can be found in the Supplementary Materials. LASSO regression, implemented in the glmnet package in R, was used to identify CpGs with independent and significant associations. The optimal hyperparameter *λ* was determined based on achieving a cross-validated error within one standard error of the minimum, as determined by 10-fold cross-validation. This process resulted in identifying 151 independent CpGs, with 87 of these maintaining significance in the joint model with 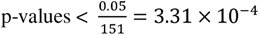.

### Validation of the CpGs using measured DNA methylation levels

We sought to validate our findings by comparing them against measured methylation levels, using the EWAS Catalog ^9^, which collects association statistics for CpG with traits from published EWAS. We downloaded the full summary statistics from the Generation Scotland (GS) cohort ^21^, calculated based on 757 incident T2D cases and 16,992 controls—the largest T2D EWAS dataset available in the catalog. This validation process successfully confirmed 10 of our identified CpGs, with 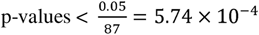.

### MWAS for cardiometabolic traits

We extended our analysis to investigate the effects of the 87 independent and significant CpGs on cardiometabolic traits, which are closely related to T2D. This included examining traits such as Body Mass Index (BMI), fasting glucose, HbA1c, systolic blood pressure (SBP), diastolic blood pressure (DBP), pulse pressure (PP), total cholesterol (TC), triglycerides (TG), low-density lipoprotein cholesterol (LDL-C), and high-density lipoprotein cholesterol (HDL-C). For this purpose, we accessed GWAS summary statistics for these traits in European populations from the GWAS Catalog ^22–26^. Using S-PrediXcan, we then conducted MWAS analyses for each trait, employing Fryett’s DNAm prediction model alongside the respective trait’s GWAS summary statistics.

### Trans-ethnic analysis of the significant CpGs

To explore the broader applicability of our findings, we assessed whether the 87 independent and significant CpGs have trans-ethnic effects on T2D among East Asians. We also used a two-sample MR analysis to investigate the causal influence of these CpGs on T2D. The meQTL summary statistics were derived from 2,150 samples profiled based on the Illumina Infinium MethylationEPIC BeadChip, with GWAS data from the Taiwan Biobank. Supplementary Materials provide in-depth details regarding the QC procedures for the Taiwan Biobank samples and the methodology for computing the meQTL summary statistics.

GWAS summary statistics for T2D were obtained from the Asian Genetic Epidemiology Network (AGEN) consortium, based on a cohort of 77,418 T2D cases and 356,122 controls ^27^. Of the 87 significant CpGs, 56 possessed 3 or more instrumental meQTLs. Within this subset, 16 CpGs were found to exhibit significant MR effects in the East Asian population, achieving MR p-values less than 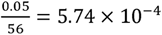 and demonstrating no evidence of pleiotropy (MR-Egger intercept p-values > 0.05).

### Expression quantitative trait methylation (eQTM) analysis

To investigate whether the 87 significant CpGs function as expression quantitative trait methylation (eQTM) loci in human pancreatic alpha and beta-cells, key to the development and progression of diabetes, we utilized whole-genome bisulfite sequencing (WGBS) and RNA sequencing (RNA-Seq) data from PANC-DB ^28^. The processing details for the WGBS and RNA-Seq data are elaborated in the Supplementary Materials. Our analysis was confined to European-descent samples without a diabetes history, comprising 17 alpha-cell samples and 10 beta-cell samples with both WGBS and RNA-Seq data.

Initially, we modeled the transcript per million (TPM) for each gene using a linear regression model, accounting for age and sex. We then calculated the residuals from this model to assess their correlation with methylation levels, employing Kendall’s τ coefficient. The significance of these correlations was determined using the non-parametric τ test. Given that CpGs often regulate gene expression from positions upstream, our focus was on CpGs located within 100 kb upstream of genes. Due to the small sample sizes, genes with correlation p-values < 0.2 were selected for subsequent pathway and differential gene expression analysis. In the alpha cells, we identified 23 CpGs that exhibited associations, albeit moderate, with the expression levels of 24 genes, evidenced by p-values < 0.2. Meanwhile, in the beta cells, 12 CpGs were found to have potential associations with the expression of 16 genes.

### Pathway analysis

To further understand the biological implications of the 24 and 16 genes potentially regulated by CpGs in alpha and beta cells, respectively, as identified in the eQTM analysis, we conducted a pathway analysis using GeneNetwork v2.0 ^29^. This analysis aimed to determine if these genes are significantly enriched in specific biological pathways. We utilized both the Reactome and KEGG pathway databases for this assessment. This approach allows for the elucidation of the potential functional roles and biological processes that these genes may be involved in, shedding light on their contributions to the pathophysiology of T2D.

### Differential gene expression analysis

To explore the expression dynamics of genes potentially regulated by the CpGs identified in the eQTM analysis, we conducted a differential gene expression analysis between T2D cases and controls without a diabetes history. This analysis utilized RNA-Seq data from PANC-DB, focusing on samples from pancreatic alpha and beta cells. Specifically, the dataset included 6 T2D patients and 26 controls for alpha cells, and 6 T2D patients and 22 controls for beta cells.

We first adjusted the TPM for each gene by age and sex using a linear regression model, then derived residuals for further analysis. These residuals were assessed for their association with T2D status employing a permutation test. Detailed methodology of the permutation test is elaborated in the Supplementary Materials. Genes that exhibited significant associations with T2D status, indicated by permutation p-values < 0.05, were earmarked for subsequent functional analysis, aiming to uncover the biological implications of these expression differences in the context of T2D.

### Functional analysis in mouse models

To delve deeper into the functional significance of the genes identified in the differential gene expression analysis, we searched the Mouse Genome Informatics (MGI) database. This comprehensive resource integrates data from the Mouse Genome Database (MGD) ^30^, Mouse Gene Expression Database (GXD) ^31^, and Mouse Tumor Biology Database (MMHCdb) ^32^. Our investigation focused on examining the phenotypic outcomes of mutational changes in these genes, aiming to infer potential implications for diabetes pathogenesis and progression. This approach allows us to extrapolate the biological impact of these genes based on phenotypic alterations observed in mouse models, providing valuable insights into their roles in disease mechanisms.

## Results

Our analysis workflow is illustrated in Figure 1, and the resources utilized throughout our study are detailed in Table S1. The MWAS and MR analyses primarily leveraged GWAS summary statistics from datasets comprising hundreds of thousands of samples. Through conducting a MWAS for T2D, followed by replication efforts via MR and joint modeling using LASSO regression on the UK Biobank dataset, we identified 87 significant, independent CpGs. Notably, 13 of these CpGs are located within or proximal to genes not previously highlighted as T2D candidate genes in the GWAS catalog or the Type 2 Diabetes Knowledge Portal ^33^. Detailed MWAS and MR statistics for these 13 novel CpGs are presented in Table 1, whereas comprehensive details regarding all 87 CpGs can be found in Supplementary Table S2. In Tables 1 and S2, the direction of effect generally concurs across MWAS and MR analyses, where a positive effect signifies that an increase in genetically predicted DNA methylation (DNAm) heightens T2D risk.

**Figure 1.**
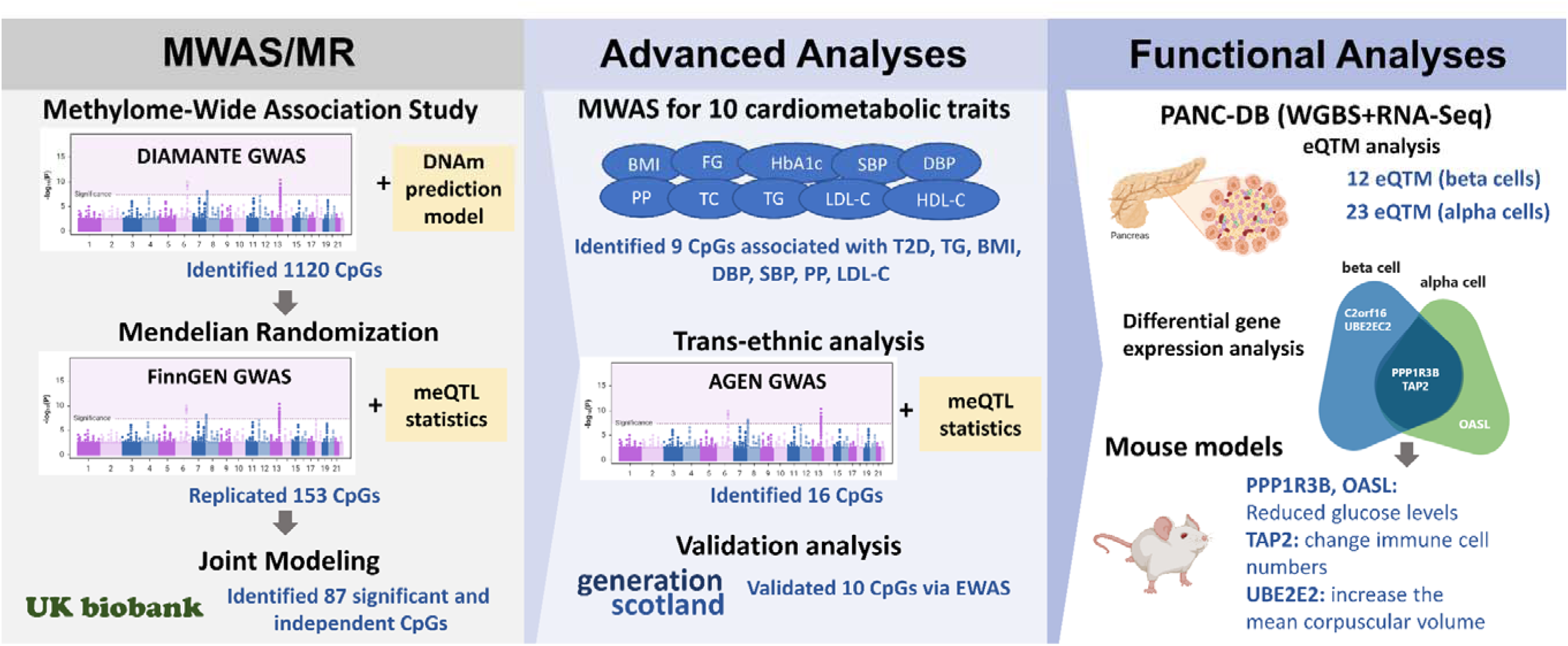
Study Overview and Key Findings. **MWAS/MR:** Methylome-Wide Association Study (MWAS) identified 1,120 CpGs associated with type 2 diabetes (T2D) in Europeans, while 153 of them were replicated through Mendelian Randomization (MR) in an independent dataset. Using UK Biobank, 87 of the 153 CpGs were jointly independent. **Advanced Analyses:** For the 87 CpGs, MWAS assessed their association with 10 cardiometabolic traits related to T2D, and 9 were significantly associated with T2D, triglycerides (TG), BMI, diastolic and systolic blood pressure (DBP and SBP), pulse pressure (PP), and low-density lipoprotein cholesterol (LDL-C). A trans-ethnic analysis identified 16 of the 87 CpGs demonstrating trans-ethnic effects in East Asians. A validation analysis using samples measured with DNA methylation levels identified 10 significant CpGs. **Functional Analyses:** Through whole-genome bisulfite sequencing (WGBS) and RNA-Seq analyses using data from PANC-DB, we identified 10 and 23 CpGs acting as potential expression quantitative trait methylation (eQTM) in pancreatic alpha and beta cells, respectively. Differential gene expression analysis identified genes regulated by the eQTM differentially expressed in alpha and beta cells. Mouse models supported the functional roles of PPP1R3B and OASL genes in glucose homeostasis.

**Table 1.**
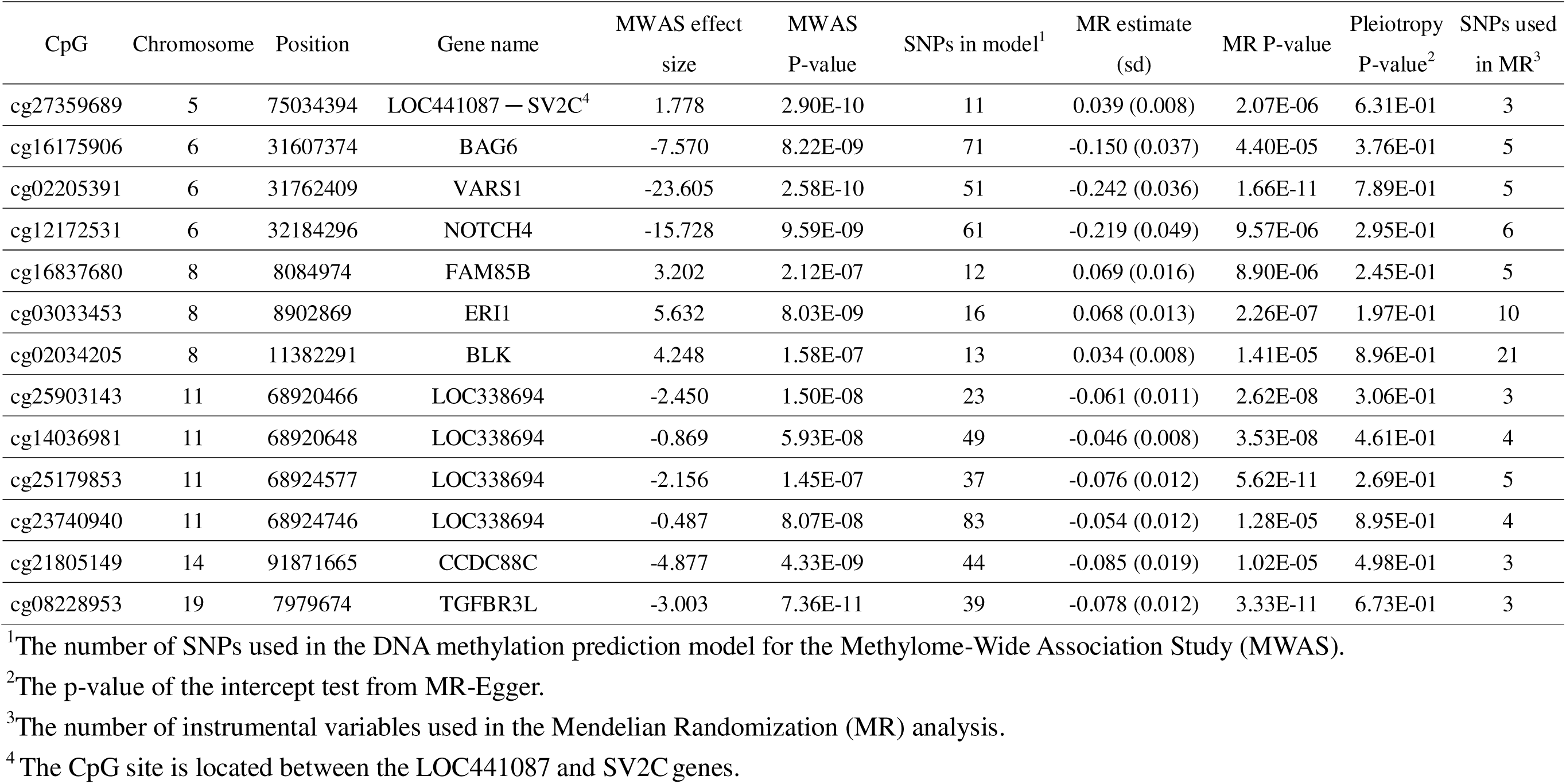
Significant and independent CpGs in novel genes for type 2 diabetes.

Supplementary Table S3. outlines the lead SNPs—those with the smallest GWAS p-values— associated with each of the 87 CpGs, identified either in the MWAS as prediction variables or in the MR analysis as instrumental variables. These lead SNPs generally exhibited significant GWAS p-values < 5 × 10^−8^. The median proximity of these SNPs to the corresponding CpGs was approximately 27 kb in the MWAS context and about 50 kb in the MR analysis. Notably, for CpGs located in novel genes (highlighted in bold in the Table), the average distance to their lead SNPs was approximately 108 kb in MWAS and about 231 kb in MR analysis, indicating the intricate influence of GWAS-identified SNPs on the epigenetic regulation.

Out of the 87 identified CpG sites, 10 were successfully validated through measured DNAm levels from the GS study. Their summary statistics are shown in Supplementary Table S4. The validation of these CpGs, despite the relatively modest cohort size of 757 T2D cases, suggests that the genetically regulated aspects of methylation at these 10 sites have a substantial impact.

Figure 2 presents an upset plot of the MWAS results for the 87 CpGs across the 10 cardiometabolic traits, illustrating both the number of significant CpGs (with p-values < 5.74 × 10^−4^ considering 87 tests) associated with each trait and the intersections among these significant CpGs across different traits. Notably, TG showed the most substantial overlap with T2D, featuring 41 significant CpGs, followed by BMI with 33, and HDL-C with 31 significant CpGs within the set of 87. Additionally, TG, BMI, DBP, SBP, PP, and LDL-C collectively exhibited the most considerable overlap with T2D, evidenced by 9 CpGs being significant across these traits. FG was unique in having 6 distinct significant CpGs that overlapped with T2D but not with the other traits. Supplementary Table S5 contains comprehensive details of the MWAS findings.

**Figure 2.**
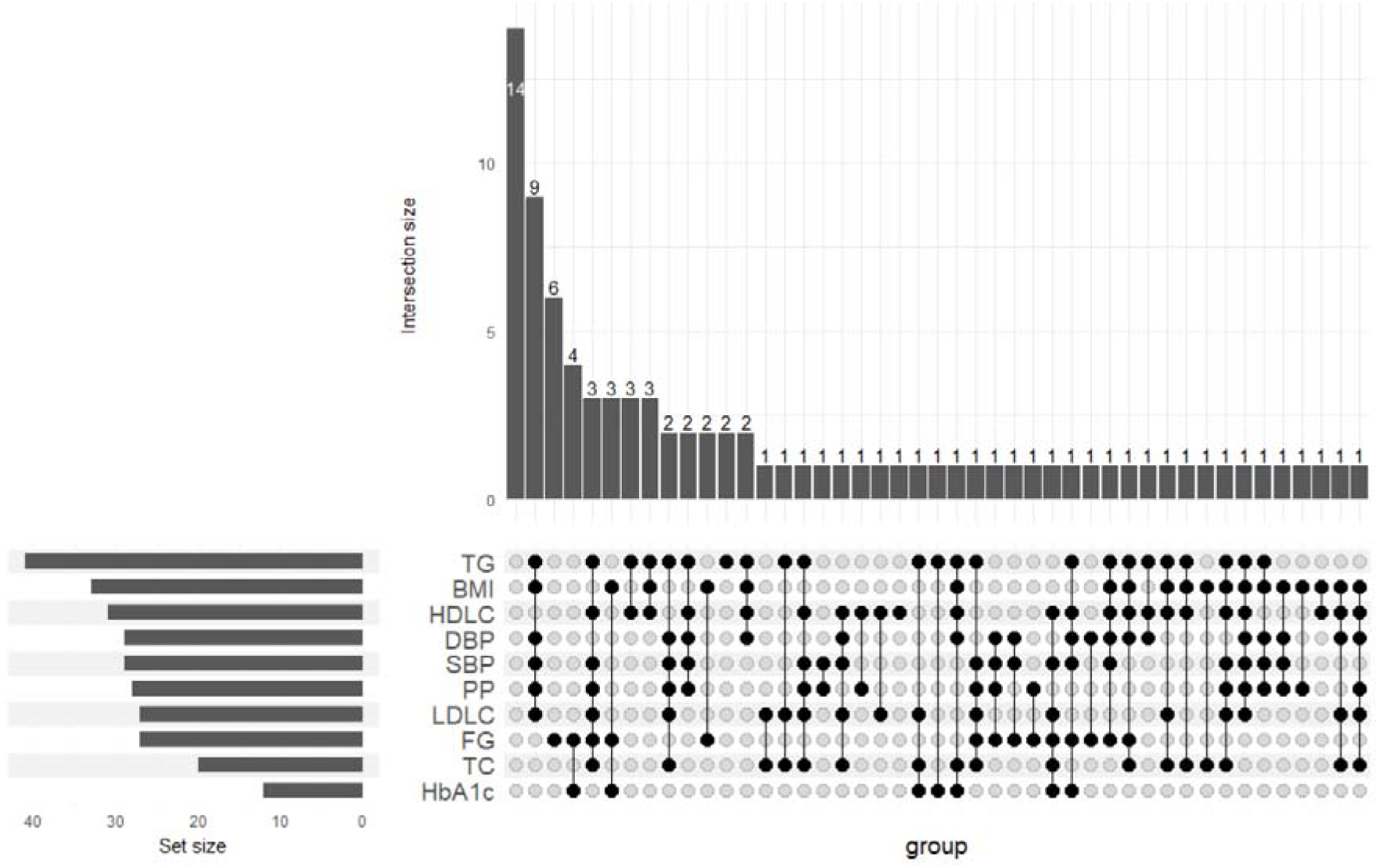
Intersection Analysis of MWAS Findings for 87 CpGs Across Cardiometabolic Traits. This figure presents an upset plot visualizing the distribution of significant CpG sites identified through MWAS among 10 cardiometabolic traits associated with T2D. The “Set size” section quantifies the significant CpGs associated with each individual trait. The “Intersection size” part details the count of CpGs that are significant across multiple traits. Traits associated with each significant CpG group are denoted by black dots under the “group” category. Key traits include triglycerides (TG), body mass index (BMI), high-density lipoprotein cholesterol (HDLC), diastolic blood pressure (DBP), systolic blood pressure (SBP), pulse pressure (PP), low-density lipoprotein cholesterol (LDLC), fasting glucose (FG), total cholesterol (TC), and HbA1c.

Table 2 shows the 16 CpGs demonstrating trans-ethnic effects in East Asians based on the two-sample MR analysis. Among these CpGs, two are located in genes newly identified in our study for T2D: VARS1 and FAM85B. Additionally, Table 2 compares the directions of effects of these CpGs between European and East Asian populations. Notably, all 16 CpGs showed consistent directions of effect across both groups, underscoring their potential universal biological relevance to T2D.

**Table 2.**
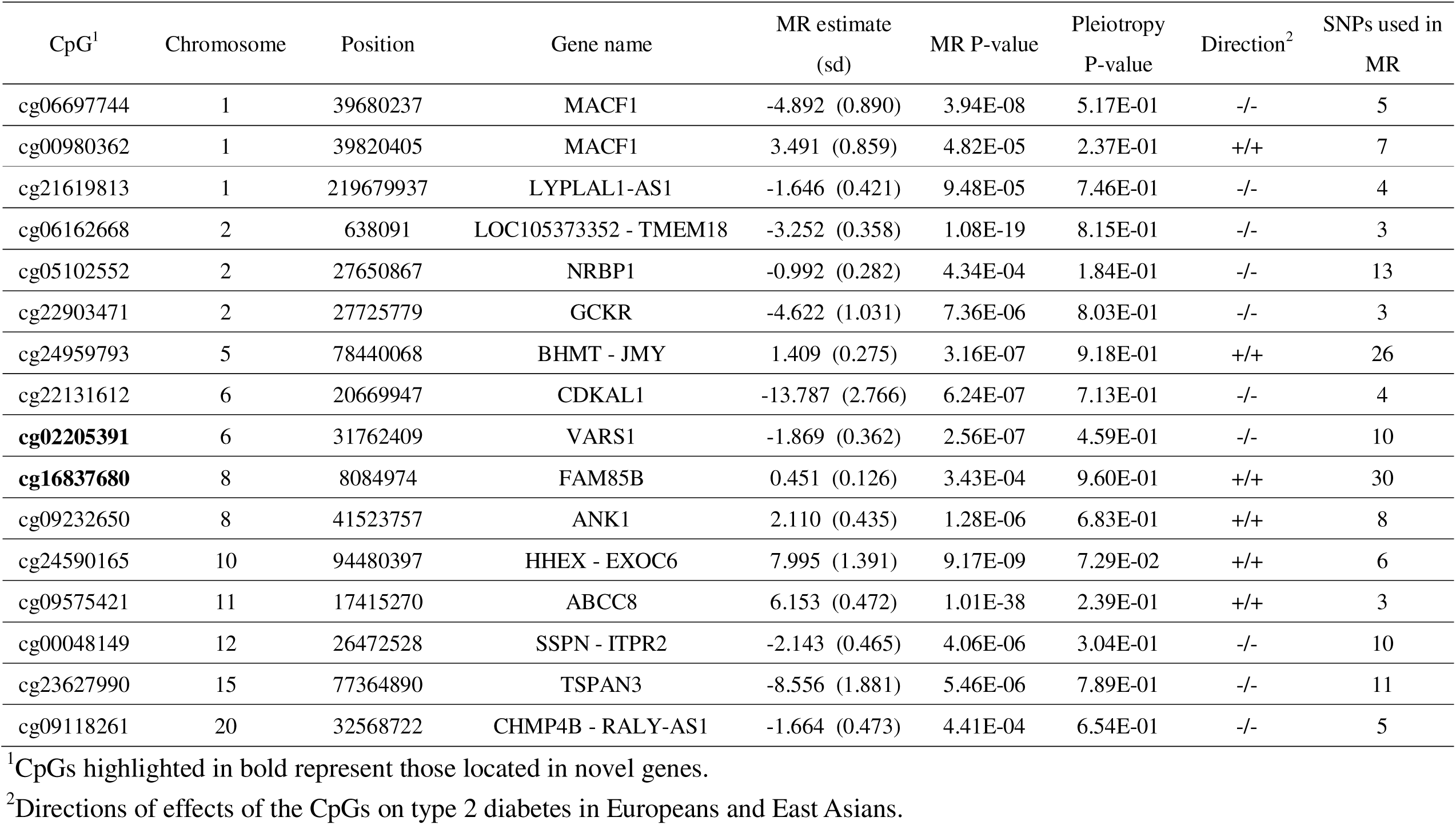
CpGs with trans-ethnic effects in East Asians.

We assessed whether the 87 CpGs could be potential eQTM in alpha and beta pancreatic cells. In alpha cells, 23 CpGs were found to be somewhat associated with the expression of 24 genes, evidenced by p-values < 0.2. Meanwhile, in beta cells, 12 CpGs were associated with the expression of 16 genes. Notably, five genes—KRTCAP3, UBE2E2, TAP2, PPP1R3B, and AXIN1—showed overlap in findings between alpha and beta cells, with their association statistics detailed in Table 3. Comprehensive association statistics for all 87 CpGs in alpha and beta cells are presented in Supplementary Tables S6 and S7, respectively.

**Table 3.** eQTM statistics for CpGs and the genes they regulate, with overlap in alpha and beta cells.

| CpG |  |  |  | Regulated Gene |  |  |  | eQTM Statistics |  | Cell Type |
| --- | --- | --- | --- | --- | --- | --- | --- | --- | --- | --- |
| CpG | Chromosome | Position | Gene | Gene | Chromosome | START | END | beta | p-value |  |
| cg01969012 | 2 | 27650506 | NRBP1 | KRTCAP3 | 2 | 27666921 | 27666923 | 0.294 | 1.05E-01 | alpha |
| cg01969012 | 2 | 27650506 | NRBP1 | KRTCAP3 | 2 | 27666921 | 27666923 | 0.568 | 2.42E-02 | beta |
| cg21007371 | 3 | 23244130 | UBE2E2 | UBE2E2 | 3 | 23267264 | 23267266 | -0.279 | 1.50E-01 | alpha |
| cg21007371 | 3 | 23244130 | UBE2E2 | UBE2E2 | 3 | 23267264 | 23267266 | 0.442 | 8.33E-02 | beta |
| cg06598146 | 6 | 32711632 | HLA-DQA2 | TAP2 | 6 | 32796683 | 32796685 | 0.379 | 3.77E-02 | alpha |
| cg14255617 | 6 | 32729118 | HLA-DQB2 | TAP2 | 6 | 32796683 | 32796685 | 0.449 | 7.25E-02 | beta |
| cg03033453 | 8 | 8902869 | ERI1 | PPP1R3B | 8 | 8998304 | 8998306 | -0.406 | 2.34E-02 | alpha |
| cg03033453 | 8 | 8902869 | ERI1 | PPP1R3B | 8 | 8998304 | 8998306 | -0.454 | 7.81E-02 | beta |
| cg01474600 | 16 | 315564 | FAM234A | AXIN1 | 16 | 338122 | 338124 | 0.241 | 1.98E-01 | alpha |
| cg01474600 | 16 | 315564 | FAM234A | AXIN1 | 16 | 338122 | 338124 | -0.360 | 1.51E-01 | beta |

The identified genes based on the eQTM analyses were categorized into 4 clusters for alpha cells and 3 for beta cells, as illustrated in Figure 3, using GeneNetwork v2.0 for analysis. Supplementary Tables S8 and S9 list pathways with a False Discovery Rate (FDR) < 0.2 in alpha and beta cells, respectively. Among these, pathways pertinent to glucose metabolism were identified, including REACTOME:R-HSA-70263 (Gluconeogenesis) with an FDR of 0.15 in alpha cells and REACTOME:R-HSA-112399 (Insulin Receptor Substrate (IRS)-mediated signalling) with an FDR of 0.10 in beta cells, suggesting a functional link to glucose regulation mechanisms.

**Figure 3:**
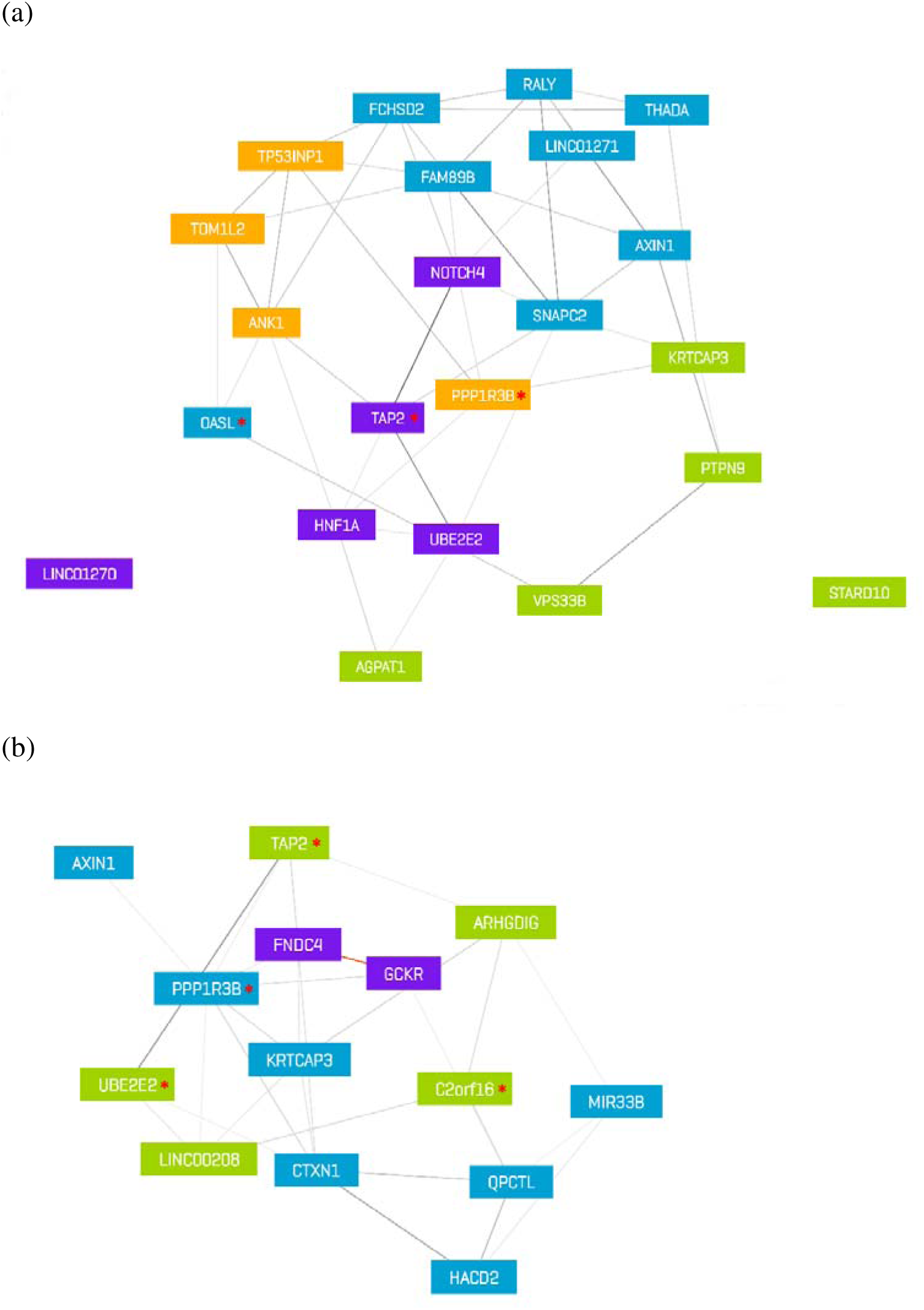
Gene Regulatory Networks Influenced by CpG Sites in Pancreatic Cell Types. Panel (a) displays the network for alpha cells, highlighting genes affected by CpG site regulation. Panel (b) illustrates the corresponding network for beta cells. Genes designated with an asterisk (*) represent those exhibiting differential expression between type 2 diabetes (T2D) cases and control subjects.

In Figure 3, genes that exhibited differential expression with p-values < 0.05 between T2D patients and controls in pancreatic alpha and beta cells are highlighted. Specifically, three genes—PPP1R3B, TAP2, and OASL—were identified as significant in alpha cells, whereas four genes—C2orf16, TAP2, PPP1R3B, and UBE2E2—showed significance in beta cells. The detailed p-values for differential expression are also provided in Supplementary Tables S6 and S7. Supplementary Figures S1 to S7 display the plots for the residuals of gene expression levels, adjusted for age and sex, for these seven genes between the two groups in both cell types.

The results of functional analyses conducted on these genes in mouse models are detailed in Supplementary Table S10. Notably, mutations in the mouse Ppp1r3b and Oasl2 genes were associated with reduced circulating glucose levels, indicating a potential role in glucose metabolism. Mutations in the mouse Tap2 gene were linked to changes in immune cell numbers, highlighting its involvement in immune responses. Furthermore, mutations in Ube2e2 were observed to increase the mean corpuscular volume, suggesting an impact on erythrocyte properties. These findings underscore the biological significance of these genes in both glucose metabolism and broader physiological processes.

## Discussion

The findings from our comprehensive multi-omics analysis provide significant insights into the influence of GWAS-identified SNPs on the epigenetic architecture for T2D and their association with cardiometabolic traits. Our results demonstrated that these GWAS SNPs influenced DNAm in blood samples at CpGs that could be located tens or hundreds of base pairs away. Notably, certain CpGs we identified are likely to regulate gene expression in human pancreatic alpha and beta cells, and DNAm levels at these CpGs, regulated by T2D SNPs, may lead to differential gene expression of PPP1R3B, TAP2, and OASL in alpha cells and C2orf16, TAP2, PPP1R3B, and UBE2E2 in beta cells, when comparing normal controls to T2D patients. Evidence from mouse models further corroborates the critical roles of PPP1R3B and OASL in glucose homeostasis, reinforcing the biological relevance of our findings. Additionally, research on UBE2E2 in mice indicates its essential function in glucose homeostasis and β-cell mass regulation via ubiquitin modifications, with its overexpression leading to reduced insulin secretion and β-cell proliferation ^34^.

The identification of 13 novel CpGs located in or near genes not previously associated with T2D through GWAS highlights the untapped potential of epigenetic modifications in revealing new genetic susceptibilities to metabolic diseases. The presence of these CpGs in novel genes such as NOTCH4, and its subsequent validation in the GS study suggests that genetically regulated methylation levels play a substantial role in T2D development. The Notch signaling pathway, crucial for cell fate decisions and involved in the modulation of essential transcriptional regulators such as Hes1, plays a significant role in pancreatic development and the differentiation of pancreatic progenitor cells ^35^. The findings that specific Notch pathway components like DLL1 and DLL4 in β-cells and JAGGED1 in α-cells have distinct roles in adult pancreatic islets, affecting glucose tolerance and insulin secretion, further delineate the intricate involvement of Notch signaling in T2D ^36^. The linkage of NOTCH4’s role in T2D through the modulation of PI3K/AKT-dependent insulin signaling to Notch signaling emphasizes its critical function in regulating glucose homeostasis and offers promising insights into developing targeted therapies for T2D ^37–40^, suggesting a broader mechanism where genetic and epigenetic factors intersect in the disease’s pathophysiology.

The upset plot showing the intersections of significant CpGs among 10 cardiometabolic traits provides valuable insights into the shared epigenetic basis of these conditions. The substantial overlap of significant CpGs between T2D and traits such as TG, BMI, blood pressure, and LDL-C emphasizes the interconnectedness of metabolic diseases at the epigenetic level. The role of DNAm in T2D appears intricately linked to the regulation of inflammatory markers, such as Monocyte Chemoattractant Protein-1 (MCP-1), crucial for the migration and infiltration of various immune cells ^41^. Findings from a study suggest that in individuals with T2D, there is a notable hypomethylation of the MCP-1 promoter CpG sites, which correlates with elevated MCP-1 serum levels. This hypomethylation is associated with increased levels of blood glucose and TG, conditions prevalent in T2D, indicating a potential mechanism through which metabolic dysregulations influence inflammatory processes ^42^. Additionally, the negative correlation between MCP-1 promoter methylation and markers of metabolic syndrome, such as BMI, blood pressure, and lipid traits ^42^, underscores the complex interplay between epigenetic modifications and metabolic health, suggesting that DNA methylation could modulate the severity or progression of T2D through its impact on inflammatory pathways and metabolic markers.

The identification of 16 CpGs with trans-ethnic effects in East Asians underscores the universal relevance of these epigenetic markers across populations. The consistent direction of effects in both Europeans and East Asians enhances the generalizability of our findings and suggests that these CpGs could serve as global biomarkers for T2D risk.

Additionally, the evaluation of these 87 CpGs as potential eQTM in human pancreatic alpha and beta cells provides a deeper understanding of how these CpGs influence gene expression in cell types crucial for glucose homeostasis. The pathway analysis in alpha and beta cells reveals associations with pathways related to glucose genesis, such as gluconeogenesis and IRS-mediated signaling. The presence of gluconeogenesis-related genes in alpha cells suggests a nuanced role for these cells in glucose production, extending beyond glucagon secretion to possibly direct regulation of hepatic glucose output. On the other hand, the enrichment of IRS-mediated signaling genes in beta cells underscores the critical function of these cells in insulin sensitivity and signaling, essential for glucose uptake and regulation. These discoveries highlight the complex interplay between pancreatic cell types in managing blood glucose levels.

The findings that PPP1R3B, key in glycogen metabolism, is differentially expressed both in pancreatic alpha and beta cells, along with mouse models showing its mutations can improve glucose tolerance and lower glucose levels, provide new paths for diabetes treatment. This gene, crucial for glycogen synthesis, plays a significant role in glucose storage and its bloodstream availability, suggesting its involvement in both glucagon and insulin pathways which could affect glycogen storage, gluconeogenesis, and insulin sensitivity, offering a therapeutic target for T2D ^43^. Similarly, the unique expression of OASL in alpha cells and its mutation-induced glucose level reduction highlight a novel link between immune mechanisms and metabolic regulation. OASL, through its modulation of the type I interferon (IFN) response, plays a significant role in the innate immune system’s response to various pathogens, including its dual role in enhancing antiviral defenses and inhibiting autophagic mechanisms ^44^. While type I IFN signaling, to which OASL contributes, is involved more directly in type 1 diabetes (T1D) in terms of immune system regulation ^45^, its role in modulating the immune response suggests potential implications for T2D as well, given the chronic inflammation and innate immune system dysfunction associated with T2D. These discoveries underscore the potential of targeting PPP1R3B and OASL for diabetes management, emphasizing the need to understand gene functions across cell types for effective interventions.

While our study has uncovered significant associations and novel CpGs, it is not without limitations. The sample size for some analyses, such as the GS study and PANC-DB, was relatively small, which may affect the generalizability of those findings. Future studies with larger cohorts are necessary to validate our results and to explore the functional consequences of methylation changes in greater detail. Moreover, while the MWAS and MR associations were observed in whole blood samples, we have used human pancreatic alpha and beta cells to delve into their specific functional roles in relation to the pathology of T2D. However, investigating the effects in other tissue types, including adipose tissue, liver, and skeletal muscles, could provide further insights into the systemic nature of T2D and its epigenetic regulation.

In conclusion, our study underscores the importance of epigenetic mechanisms in the etiology of T2D and related cardiometabolic diseases. The identification of novel CpGs and their association with T2D risk opens new avenues for research into the genetic and epigenetic underpinnings of metabolic diseases. This could potentially lead to the development of novel diagnostic and therapeutic strategies that target the epigenetic landscape of T2D.

## Data Availability

The Taiwan Biobank data can be applied through the Taiwan Biobank at https://www.twbiobank.org.tw/. The sources for publicly available datasets used in this study are provided in Table S1 in the Supplementary Tables.

## Acknowledgements

We thank the participants from the UK and Taiwan Biobanks. This manuscript used data acquired from the Human Pancreas Analysis Program (HPAP-RRID:SCR_016202) Database (https://hpap.pmacs.upenn.edu), a Human Islet Research Network (RRID:SCR_014393) consortium (UC4-DK-112217, U01-DK-123594, UC4-DK-112232, and U01-DK-123716). This study was supported by grants PH-112-GP-04 and PH-112-PP-10 from the National Health Research Institutes and MOST 111-2221-E-400-004 from the National Science and Technology Council in Taiwan.

## Notes

### Competing Interest Statement

The authors have declared no competing interest.

### Author Declarations

Ethics committee/IRB of National Health Research Institutes gave ethical approval for this work.

