## Supplementary Tables for "Elucidating the Epigenetic Landscape of Type 2 Diabetes: A Multi-Omics Analysis Revealing Novel CpG Sites and Their Association with Cardiometabolic Traits"

**Supplmentary Table S1. Datasets/models used in our analysis**

| Dataset/Model | Type | Sample size | Ethnicity | Purpopose | Reference |
|---|---|---|---|---|---|
| Understanding Society: UK Household Longitudinal Studay | DNAm prediction model trained on GWAS and DNAm data | 1,120 | European | MWAS | https://pubmed.ncbi.nlm.nih.gov/35930604/ |
| DIAMANTE | GWAS summary statistics | 80,154 T2D cases, 853,816 controls | European | MWAS | https://pubmed.ncbi.nlm.nih.gov/30297969/ |
| FinnGEN | GWAS summary statistics | 38,657 T2D cases, 310,131 controls | European | MR | https://r9.finngen.fi/ |
| BMI GWAS | GWAS summary statistics | 806,834 with BMI measurements | European | MWAS | https://pubmed.ncbi.nlm.nih.gov/30239722/ |
| Fasting glucose GWAS | GWAS summary statistics | 200,622 with fasting glucose measurements | European | MWAS | https://pubmed.ncbi.nlm.nih.gov/34059833/ |
| HbA1c GWAS | GWAS summary statistics | 146,806 with HbA1c measurements | European | MWAS | https://pubmed.ncbi.nlm.nih.gov/34059833/ |
| Systolic blood pressure (SBP) GWAS | GWAS summary statistics | 757,601 with SBP measurements | European | MWAS | https://pubmed.ncbi.nlm.nih.gov/30224653/ |
| Diastolic blood pressure (DBP) GWAS | GWAS summary statistics | 757,601 with DBP measurements | European | MWAS | https://pubmed.ncbi.nlm.nih.gov/30224653/ |
| Pulse pressure (PP) GWAS | GWAS summary statistics | 757,601 with PP measurements | European | MWAS | https://pubmed.ncbi.nlm.nih.gov/30224653/ |
| Total cholesterol GWAS | GWAS summary statistics | 1,320,016 with total cholesterol measurements | European | MWAS | https://pubmed.ncbi.nlm.nih.gov/34887591/ |
| Triglycerides (TG) GWAS | GWAS summary statistics | 1,320,016 with TG measurements | European | MWAS | https://pubmed.ncbi.nlm.nih.gov/34887591/ |
| LDL-C GWAS | GWAS summary statistics | 1,320,016 with LDL-C measurements | European | MWAS | https://pubmed.ncbi.nlm.nih.gov/34887591/ |
| HDL-C GWAS | GWAS summary statistics | 1,320,016 with HDL-C measurements | European | MWAS | https://pubmed.ncbi.nlm.nih.gov/34887591/ |
| AGEN GWAS | GWAS summary statistics | 77,418 T2D cases, 356,122 controls | East Asian | MR | https://pubmed.ncbi.nlm.nih.gov/32499647/ |
| UK Biobank | GWAS raw genotypes | 25,025 T2D prevalent cases, 314,430 controls | European | Joint analysis | https://www.ukbiobank.ac.uk/ |
| Taiwan Biobank | Methylation and GWAS chip | 2,150 samples | East Asian | MR | https://www.twbiobank.org.tw/ |
| EWAS Catalog | EWAS summary statistics | 757 incident cases, 16,992 controls | European | Validation | https://pubmed.ncbi.nlm.nih.gov/35592546/ |
| MeQTL EPIC | meQTL database | 2,358 samples | European | MR | https://pubmed.ncbi.nlm.nih.gov/37525248/ |
| PANC-DB | WGBS and RNA-Seq data in human pancreatic alpha and beta cells | eQTM analysis: 17 and 10 alpha and beta-cell samples, respectively. Differential gene expression analysis: 6 T2D cases and 26 controls for alpha cells, 6 T2D cases and 22 controls for beta cells | European | eQTM and differential gene expression analyses | https://pubmed.ncbi.nlm.nih.gov/31127054/ |

MWAS: Methylome-wide association studies; MR: Mendelian randomization; WGBS: whole-genome bisulfite sequencing;

**Supplmentary Table S2. Summary statistics for significant and independent CpGs identified by MWAS and MR for T2D**

| CpG | Chromosome | Position | Gene name | MWAS effect size | MWAS P-value | SNPs in model[1] | MR estimate | MR sd | MR P-value | Pleiotropy P-value | SNPs used in MR[2] |
|---|---|---|---|---|---|---|---|---|---|---|---|
| cg03179008 | 1 | 39620300 | MACF1 | 2.699 | 8.51E-21 | 50 | 0.066 | 0.011 | 4.85E-10 | 5.57E-01 | 5 |
| cg06697744 | 1 | 39680237 | MACF1 | -7.514 | 2.95E-16 | 35 | -0.081 | 0.016 | 3.92E-07 | 1.45E-01 | 3 |
| cg08719237 | 1 | 39728532 | MACF1 | 4.017 | 7.67E-22 | 39 | 0.080 | 0.013 | 1.25E-09 | 2.96E-01 | 3 |
| cg00980362 | 1 | 39820405 | MACF1 | 4.088 | 3.71E-25 | 35 | 0.085 | 0.016 | 9.16E-08 | 2.56E-01 | 3 |
| cg21619813 | 1 | 219679937 | LYPLAL1-AS1 | -2.697 | 1.56E-15 | 27 | -0.112 | 0.015 | 2.10E-13 | 3.44E-01 | 4 |
| cg06162668 | 2 | 638091 | LOC105373352(dist=75999),TMEM18(dist=25787) | -2.183 | 8.11E-09 | 39 | -0.049 | 0.010 | 8.40E-07 | 3.92E-01 | 8 |
| cg01969012 | 2 | 27650506 | NRBP1 | -1.520 | 3.96E-12 | 17 | -0.045 | 0.010 | 1.21E-05 | 3.60E-01 | 7 |
| cg05102552 | 2 | 27650867 | NRBP1 | -2.216 | 1.65E-11 | 12 | -0.048 | 0.008 | 8.32E-09 | 2.05E-01 | 9 |
| cg22903471 | 2 | 27725779 | GCKR | -1.464 | 5.65E-08 | 24 | -0.057 | 0.010 | 1.90E-09 | 9.24E-01 | 6 |
| cg10337841 | 2 | 27747795 | GCKR(dist=1245),C2orf16(dist=51594) | -5.864 | 8.02E-11 | 28 | -0.087 | 0.015 | 1.04E-08 | 5.24E-01 | 3 |
| cg25428544 | 2 | 43428588 | LINC02580(dist=99005),ZFP36L2(dist=20953) | 2.307 | 6.14E-08 | 24 | 0.114 | 0.019 | 8.85E-10 | 6.86E-01 | 3 |
| cg11998686 | 2 | 43687897 | THADA;THADA;THADA | 1.369 | 6.18E-27 | 93 | 0.093 | 0.011 | 1.70E-16 | 8.05E-01 | 3 |
| cg05293897 | 2 | 226913713 | NYAP2(dist=345343),LOC646736(dist=93797) | -0.629 | 8.39E-10 | 68 | -0.104 | 0.013 | 5.17E-16 | 5.33E-01 | 5 |
| cg04891645 | 2 | 227050097 | LOC646736(dist=5319),MIR5702(dist=473329) | -5.279 | 6.62E-24 | 27 | -0.175 | 0.017 | 4.67E-25 | 8.71E-01 | 5 |
| cg10397322 | 3 | 23244051 | UBE2E2 | -3.892 | 1.44E-15 | 41 | -0.075 | 0.013 | 2.53E-08 | 2.19E-01 | 8 |
| cg18257541 | 3 | 23244062 | UBE2E2 | -5.097 | 3.06E-17 | 40 | -0.076 | 0.013 | 2.12E-09 | 2.82E-01 | 5 |
| cg10632094 | 3 | 23244068 | UBE2E2 | -3.884 | 7.26E-16 | 44 | -0.073 | 0.014 | 3.54E-07 | 8.87E-02 | 6 |
| cg21007371 | 3 | 23244130 | UBE2E2 | -3.937 | 3.45E-10 | 31 | -0.108 | 0.019 | 2.01E-08 | 5.99E-01 | 5 |
| cg16258503 | 3 | 63850278 | ATXN7;THOC7 | 179.009 | 4.58E-11 | 6 | 0.068 | 0.018 | 1.00E-04 | 7.13E-02 | 3 |
| cg16021135 | 3 | 123130669 | ADCY5;ADCY5 | 2.865 | 2.88E-23 | 17 | 0.053 | 0.012 | 1.14E-05 | 1.57E-01 | 4 |
| cg12781915 | 3 | 185544099 | IGF2BP2;IGF2BP2 | -4.802 | 4.81E-19 | 34 | -0.164 | 0.020 | 1.65E-16 | 6.27E-01 | 4 |
| cg01899937 | 4 | 185723505 | ACSL1 | 2.876 | 1.75E-07 | 60 | 0.101 | 0.018 | 2.53E-08 | 6.64E-02 | 3 |
| cg20885688 | 5 | 74956618 | ANKDD1B | -1.582 | 2.28E-11 | 18 | -0.055 | 0.011 | 6.20E-07 | 2.92E-01 | 6 |
| **cg27359689** | **5** | **75034394** | **LOC441087(dist=7599),SV2C(dist=344814)** | **1.778** | **2.90E-10** | **11** | **0.039** | **0.008** | **2.07E-06** | **6.31E-01** | **3** |
| cg17746515 | 5 | 78439992 | BHMT(dist=11881),JMY(dist=91962) | 3.279 | 1.06E-07 | 43 | 0.067 | 0.011 | 4.86E-09 | 5.66E-01 | 5 |
| cg24959793 | 5 | 78440068 | BHMT(dist=11957),JMY(dist=91886) | 3.868 | 2.62E-11 | 33 | 0.053 | 0.011 | 1.88E-06 | 4.50E-01 | 4 |
| cg19277069 | 5 | 102089501 | PAM(dist=986) | -13.212 | 7.08E-08 | 8 | 0.134 | 0.032 | 2.24E-05 | 3.96E-01 | 3 |
| cg26671988 | 5 | 102089760 | PAM(dist=727) | -1.285 | 1.66E-08 | 75 | -0.087 | 0.018 | 1.16E-06 | 6.37E-01 | 4 |
| cg22131612 | 6 | 20669947 | CDKAL1 | -7.156 | 5.24E-48 | 38 | -0.322 | 0.059 | 4.47E-08 | 8.40E-01 | 4 |
| **cg16175906** | **6** | **31607374** | **BAG6** | **-7.570** | **8.22E-09** | **71** | **-0.150** | **0.037** | **4.40E-05** | **3.76E-01** | **5** |
| **cg02205391** | **6** | **31762409** | **VARS1** | **-23.605** | **2.58E-10** | **51** | **-0.242** | **0.036** | **1.66E-11** | **7.89E-01** | **5** |
| cg25426302 | 6 | 32120826 | PPT2;PRRT1 | -36.701 | 4.56E-09 | 28 | -0.166 | 0.027 | 1.28E-09 | 3.58E-01 | 6 |
| **cg12172531** | **6** | **32184296** | **NOTCH4** | **-15.728** | **9.59E-09** | **61** | **-0.219** | **0.049** | **9.57E-06** | **2.95E-01** | **6** |
| cg15710545 | 6 | 32578114 | HLA-DRB1(dist=20501),HLA-DQA1(dist=27069) | -2.697 | 1.44E-11 | 129 | -0.072 | 0.014 | 1.98E-07 | 8.43E-01 | 17 |
| cg06598146 | 6 | 32711632 | HLA-DQA2 | 5.007 | 3.85E-10 | 79 | 0.165 | 0.030 | 4.24E-08 | 6.35E-01 | 8 |
| cg14255617 | 6 | 32729118 | HLA-DQB2 | -1.758 | 2.64E-07 | 82 | -0.171 | 0.022 | 8.62E-15 | 6.79E-01 | 13 |
| cg00699601 | 6 | 32762284 | HLA-DQB2(dist=30975),HLA-DOB(dist=18256) | 0.833 | 8.48E-11 | 69 | 0.153 | 0.032 | 1.41E-06 | 3.50E-01 | 18 |
| cg12989544 | 6 | 153429875 | RGS17 | 1.735 | 2.52E-07 | 9 | 0.056 | 0.011 | 2.53E-07 | 3.82E-01 | 3 |
| **cg16837680** | **8** | **8084974** | **FAM85B** | **3.202** | **2.12E-07** | **12** | **0.069** | **0.016** | **8.90E-06** | **2.45E-01** | **5** |
| **cg03033453** | **8** | **8902869** | **ERI1** | **5.632** | **8.03E-09** | **16** | **0.068** | **0.013** | **2.26E-07** | **1.97E-01** | **10** |
| cg03502219 | 8 | 9978949 | MSRA | -2.332 | 8.41E-11 | 23 | -0.049 | 0.011 | 1.12E-05 | 3.54E-01 | 11 |
| cg01048752 | 8 | 10001523 | MSRA | 4.868 | 8.95E-09 | 21 | 0.060 | 0.014 | 3.26E-05 | 5.07E-02 | 12 |
| cg18477422 | 8 | 10773493 | XKR6 | -1.843 | 5.24E-10 | 22 | -0.071 | 0.016 | 1.64E-05 | 1.17E-01 | 4 |
| cg21725652 | 8 | 10918152 | XKR6 | 4.967 | 2.42E-07 | 47 | 0.062 | 0.010 | 1.53E-10 | 8.57E-01 | 13 |
| cg18317842 | 8 | 10921502 | XKR6 | -4.536 | 3.99E-08 | 18 | -0.065 | 0.012 | 6.45E-08 | 8.75E-01 | 8 |
| cg09697033 | 8 | 10929599 | XKR6 | 3.736 | 2.51E-10 | 30 | 0.095 | 0.021 | 5.52E-06 | 4.48E-01 | 3 |
| cg25014118 | 8 | 10948462 | XKR6 | 0.836 | 1.61E-10 | 37 | 0.031 | 0.004 | 5.30E-13 | 2.28E-01 | 43 |
| **cg02034205** | **8** | **11382291** | **BLK** | **4.248** | **1.58E-07** | **13** | **0.034** | **0.008** | **1.41E-05** | **8.96E-01** | **21** |
| cg00448191 | 8 | 41504890 | NKX6-3 | 2.670 | 6.51E-14 | 56 | 0.111 | 0.025 | 7.66E-06 | 9.76E-01 | 5 |
| cg20677018 | 8 | 41508391 | NKX6-3(dist=55) | 4.278 | 1.79E-22 | 24 | 0.130 | 0.019 | 3.18E-12 | 1.43E-01 | 3 |
| cg17274126 | 8 | 41522935 | ANK1 | -10.425 | 4.87E-23 | 23 | -0.068 | 0.016 | 2.73E-05 | 2.78E-01 | 3 |

| | | | | | | | | | | | |
|---|---|---|---|---|---|---|---|---|---|---|---|
| cg09232650 | 8 | 41523757 | ANK1 | 0.714 | 2.62E-26 | 15 | 0.074 | 0.018 | 3.95E-05 | 4.78E-01 | 4 |
| cg16467757 | 8 | 95888145 | INTS8 | 2.426 | 7.07E-10 | 8 | 0.083 | 0.016 | 2.10E-07 | 9.42E-01 | 3 |
| cg22283921 | 8 | 95961819 | TP53INP1 | -1.751 | 6.62E-13 | 44 | -0.058 | 0.010 | 3.25E-09 | 6.81E-01 | 12 |
| cg05289649 | 10 | 80925798 | ZMIZ1 | 0.808 | 1.29E-10 | 94 | 0.044 | 0.008 | 1.65E-08 | 2.86E-01 | 4 |
| cg21213154 | 10 | 80927984 | ZMIZ1 | 3.095 | 3.76E-10 | 18 | 0.060 | 0.008 | 2.10E-12 | 4.55E-01 | 3 |
| cg24590165 | 10 | 94480397 | HHEX(dist=24993),EXOC6(dist=106191) | 3.845 | 2.38E-48 | 63 | 0.165 | 0.016 | 4.67E-25 | 6.05E-01 | 3 |
| cg09575421 | 11 | 17415270 | ABCC8 | 5.894 | 9.51E-24 | 13 | 0.100 | 0.024 | 4.23E-05 | 1.37E-01 | 5 |
| cg01366692 | 11 | 65291439 | SCYL1 | 9.151 | 3.76E-12 | 13 | 0.105 | 0.026 | 5.21E-05 | 1.87E-01 | 3 |
| cg14286243 | 11 | 68867220 | TPCN2(dist=9155),LOC338694(dist=47476) | -1.064 | 5.95E-09 | 39 | -0.039 | 0.009 | 1.05E-05 | 7.17E-01 | 3 |
| **cg25903143** | **11** | **68920466** | **LOC338694** | **-2.450** | **1.50E-08** | **23** | **-0.061** | **0.011** | **2.62E-08** | **3.06E-01** | **3** |
| **cg14036981** | **11** | **68920648** | **LOC338694** | **-0.869** | **5.93E-08** | **49** | **-0.046** | **0.008** | **3.53E-08** | **4.61E-01** | **4** |
| **cg25179853** | **11** | **68924577** | **LOC338694** | **-2.156** | **1.45E-07** | **37** | **-0.076** | **0.012** | **5.62E-11** | **2.69E-01** | **5** |
| **cg23740940** | **11** | **68924746** | **LOC338694** | **-0.487** | **8.07E-08** | **83** | **-0.054** | **0.012** | **1.28E-05** | **8.95E-01** | **4** |
| cg04521279 | 11 | 72424620 | ARAP1 | -8.848 | 6.25E-21 | 15 | -0.198 | 0.023 | 9.99E-18 | 8.98E-01 | 3 |
| cg08110170 | 11 | 72474129 | STARD10 | -12.332 | 1.41E-23 | 7 | -0.137 | 0.031 | 7.97E-06 | 6.93E-01 | 4 |
| cg03878208 | 11 | 72483293 | STARD10 | -5.924 | 6.04E-21 | 22 | -0.133 | 0.027 | 7.01E-07 | 9.05E-01 | 4 |
| cg00048149 | 12 | 26472528 | SSPN(dist=84820),ITPR2(dist=15757) | -9.230 | 3.12E-09 | 25 | -0.086 | 0.014 | 5.95E-10 | 5.23E-01 | 5 |
| cg07742264 | 12 | 121419635 | HNF1A | -11.853 | 1.30E-14 | 12 | -0.246 | 0.023 | 1.40E-27 | 2.57E-01 | 3 |
| cg25150715 | 12 | 121454808 | C12orf43 | 4.825 | 2.31E-08 | 8 | 0.200 | 0.019 | 4.23E-27 | 7.58E-02 | 5 |
| **cg21805149** | **14** | **91871665** | **CCDC88C** | **-4.877** | **4.33E-09** | **44** | **-0.085** | **0.019** | **1.02E-05** | **4.98E-01** | **3** |
| cg08686008 | 15 | 41876221 | TYRO3(dist=434) | 0.432 | 2.27E-07 | 42 | 0.026 | 0.005 | 5.72E-07 | 9.07E-01 | 15 |
| cg01268058 | 15 | 75738663 | SIN3A | 1.636 | 7.65E-08 | 59 | 0.060 | 0.010 | 2.54E-10 | 6.24E-01 | 6 |
| cg23627990 | 15 | 77364890 | TSPAN3 | -1.393 | 1.21E-08 | 15 | -0.051 | 0.009 | 3.74E-09 | 6.56E-02 | 8 |
| cg02931208 | 15 | 77866341 | LOC101929457(dist=39889),LINGO1(dist=39025) | 5.496 | 3.97E-17 | 33 | 0.125 | 0.014 | 6.41E-20 | 3.84E-01 | 5 |
| cg25755215 | 15 | 90441888 | ARPIN;C15orf38-AP3S2 | -1.952 | 1.74E-13 | 49 | -0.060 | 0.015 | 5.81E-05 | 1.81E-01 | 5 |
| cg06979164 | 15 | 91530190 | PRC1-AS1;PRC1 | -1.489 | 5.68E-13 | 45 | -0.067 | 0.010 | 6.30E-11 | 1.26E-01 | 3 |
| cg01959774 | 16 | 290115 | FAM234A | -3.474 | 3.83E-13 | 29 | -0.046 | 0.012 | 8.81E-05 | 6.62E-01 | 4 |
| cg01474600 | 16 | 315564 | FAM234A | -4.165 | 1.49E-10 | 13 | -0.039 | 0.009 | 8.01E-06 | 8.98E-01 | 4 |
| cg22292753 | 17 | 17655165 | RAI1 | -7.018 | 1.49E-10 | 12 | -0.068 | 0.015 | 1.15E-05 | 8.56E-01 | 3 |
| cg22116239 | 17 | 17741676 | SREBF1 | -3.639 | 9.61E-09 | 46 | -0.048 | 0.011 | 1.89E-05 | 7.12E-01 | 3 |
| cg06518535 | 17 | 46844976 | TTLL6 | 4.300 | 1.79E-10 | 15 | 0.093 | 0.013 | 1.12E-13 | 1.63E-01 | 5 |
| **cg08228953** | **19** | **7979674** | **TGFBR3L** | **-3.003** | **7.36E-11** | **39** | **-0.078** | **0.012** | **3.33E-11** | **6.73E-01** | **3** |
| cg02942825 | 19 | 46185348 | GIPR | 2.590 | 8.70E-08 | 22 | 0.086 | 0.013 | 7.63E-11 | 4.47E-01 | 3 |
| cg09118261 | 20 | 32568722 | CHMP4B(dist=126550),RALY-AS1(dist=11572) | -3.278 | 1.67E-08 | 20 | -0.057 | 0.011 | 6.68E-08 | 3.91E-01 | 5 |
| cg17113955 | 20 | 32610490 | RALY;RALY | -2.673 | 6.20E-10 | 53 | -0.060 | 0.013 | 6.87E-06 | 1.35E-01 | 8 |
| cg17746527 | 20 | 48834262 | CEBPB(dist=25035),PELATON(dist=49761) | 3.810 | 5.23E-10 | 33 | 0.110 | 0.015 | 3.84E-14 | 5.58E-01 | 6 |

[1]The number of SNPs used in the DNA methylation prediction model for the Methylome-Wide Association Study (MWAS).

[2]The number of instrumental variables in the Mendelian Randomization (MR) analysis.

CpGs highlighted in bold are those in novel genes.

**Supplmentary Table S3. Lead SNP information from the MWAS and MR analyses**

| | | | MWAS | | | | MR | | | |
|---|---|---|---|---|---|---|---|---|---|---|
| CpG | Chromosome | Position | Lead SNP | DIAMANTE P-value | weight[1] | Distance[2] | Lead SNP | FinnGen P-value | meQTL P-value | Distance[2] |
| cg03179008 | 1 | 39620300 | rs3768301 | 2.81E-25 | -4.39E-05 | -250493 | rs16837533 | 1.21E-08 | 9.23E-117 | -83547 |
| cg06697744 | 1 | 39680237 | rs16837533 | 1.03E-19 | 2.31E-03 | -23610 | rs1775654 | 9.57E-08 | 6.44E-28 | -269843 |
| cg08719237 | 1 | 39728532 | rs16826069 | 2.81E-25 | -1.10E-03 | -68523 | rs56809193 | 1.51E-08 | 3.01E-62 | 119234 |
| cg00980362 | 1 | 39820405 | rs61779275 | 1.07E-25 | -3.77E-04 | 95 | rs16837533 | 1.21E-08 | 1.76E-40 | 116558 |
| cg21619813 | 1 | 219679937 | rs2820441 | 8.56E-16 | -2.53E-04 | -55023 | rs59584436 | 6.00E-09 | 4.51E-34 | 15309 |
| cg06162668 | 2 | 638091 | rs1320330 | 5.72E-09 | 4.54E-04 | 15866 | rs4854331 | 1.20E-04 | 1.06E-43 | 20470 |
| cg01969012 | 2 | 27650506 | rs780094 | 9.59E-22 | 2.90E-04 | -90731 | rs7564625 | 2.42E-03 | 3.06E-39 | -211891 |
| cg05102552 | 2 | 27650867 | rs780110 | 1.67E-11 | 2.91E-03 | -34521 | rs4665986 | 2.66E-04 | 5.39E-73 | -104301 |
| cg22903471 | 2 | 27725779 | rs3845687 | 1.68E-08 | 8.56E-04 | 35880 | rs3749147 | 5.27E-04 | 6.67E-25 | -126139 |
| cg10337841 | 2 | 27747795 | rs780110 | 1.67E-11 | 6.31E-04 | 62407 | rs780105 | 8.27E-06 | 8.93E-46 | 69324 |
| cg25428544 | 2 | 43428588 | rs10171620 | 7.92E-10 | -4.15E-05 | -470 | rs55770424 | 3.16E-06 | 1.30E-09 | -54638 |
| cg11998686 | 2 | 43687897 | rs17030845 | 2.35E-28 | 1.19E-02 | 18 | rs17031016 | 4.89E-09 | 4.16E-95 | -76545 |
| cg05293897 | 2 | 226913713 | rs2713538 | 2.51E-24 | 1.17E-05 | -220803 | rs60271363 | 1.75E-09 | 3.96E-40 | 13391 |
| cg04891645 | 2 | 227050097 | rs10439362 | 3.28E-34 | -5.02E-03 | 23361 | rs952227 | 1.76E-19 | 1.12E-60 | -11983 |
| cg10397322 | 3 | 23244051 | rs1496653 | 2.49E-17 | -9.56E-04 | -210739 | rs1496653 | 1.81E-10 | 2.37E-65 | -210739 |
| cg18257541 | 3 | 23244062 | rs1496653 | 2.49E-17 | -9.24E-04 | -210728 | rs11926494 | 1.10E-06 | 5.83E-22 | -14552 |
| cg10632094 | 3 | 23244068 | rs1496653 | 2.49E-17 | -7.74E-04 | -210722 | rs1908475 | 1.10E-06 | 5.00E-18 | -24088 |
| cg21007371 | 3 | 23244130 | rs34513914 | 2.08E-10 | -1.54E-03 | -5383 | rs1496653 | 1.81E-10 | 1.20E-36 | -210660 |
| cg16258503 | 3 | 63850278 | rs704362 | 2.13E-11 | -2.77E-04 | -28062 | rs11922435 | 1.33E-04 | 1.18E-04 | -140529 |
| cg16021135 | 3 | 123130669 | rs6798189 | 5.28E-26 | 2.17E-03 | 35357 | rs7612096 | 1.48E-05 | 1.54E-84 | -5579 |
| cg12781915 | 3 | 185544099 | rs6444082 | 3.56E-35 | 2.25E-03 | 7876 | rs28485641 | 1.47E-08 | 2.89E-15 | 132483 |
| cg01899937 | 4 | 185723505 | rs12504993 | 9.67E-14 | 1.68E-05 | 5019 | rs6552828 | 1.03E-07 | 8.36E-12 | -1911 |
| cg20885688 | 5 | 74956618 | rs6864091 | 7.46E-13 | -1.05E-03 | -53384 | rs6881648 | 9.75E-07 | 3.06E-100 | -35231 |
| **cg27359689** | **5** | **75034394** | **rs42302** | **6.66E-12** | **-1.05E-03** | **68840** | **rs7715806** | **1.66E-04** | **8.58E-171** | **7** |
| cg17746515 | 5 | 78439992 | rs2364594 | 1.69E-10 | 7.73E-04 | -13907 | rs35893235 | 3.49E-04 | 8.34E-62 | -154825 |
| cg24959793 | 5 | 78440068 | rs2121107 | 2.05E-10 | 1.00E-03 | -27936 | rs1717565 | 4.67E-04 | 4.11E-108 | -301 |
| cg19277069 | 5 | 102089501 | rs258244 | 2.42E-08 | -1.12E-04 | -72250 | rs58786975 | 9.67E-05 | 2.17E-04 | 511391 |
| cg26671988 | 5 | 102089760 | rs17296280 | 8.02E-11 | 4.22E-03 | -270987 | rs58786975 | 9.67E-05 | 3.25E-11 | 511650 |
| cg22131612 | 6 | 20669947 | rs9350271 | 1.62E-71 | 1.71E-03 | -13217 | rs7756992 | 2.08E-42 | 3.48E-09 | -9762 |
| **cg16175906** | **6** | **31607374** | **rs2844483** | **6.21E-11** | **9.25E-05** | **70578** | **rs3096698** | **5.71E-14** | **1.94E-21** | **-499765** |
| **cg02205391** | **6** | **31762409** | **rs9267654** | **1.95E-11** | **2.18E-03** | **-78600** | **rs9271351** | **2.67E-17** | **1.06E-13** | **-821451** |
| cg25426302 | 6 | 32120826 | rs9267576 | 1.41E-14 | 5.15E-04 | 308788 | rs3132946 | 4.36E-13 | 6.09E-30 | -69202 |
| **cg12172531** | **6** | **32184296** | **rs2071590** | **3.09E-08** | **6.18E-04** | **644528** | **rs436388** | **1.04E-26** | **9.44E-37** | **-1968** |
| cg15710545 | 6 | 32578114 | rs9271423 | 5.18E-16 | 4.49E-03 | -9783 | rs9269041 | 1.79E-35 | 1.71E-187 | 139871 |
| cg06598146 | 6 | 32711632 | rs1063355 | 1.38E-13 | -1.26E-03 | 83918 | rs9270908 | 1.74E-29 | 1.12E-24 | 139484 |
| cg14255617 | 6 | 32729118 | rs9273528 | 5.38E-12 | 6.14E-03 | 100485 | rs9275163 | 2.89E-73 | 4.05E-76 | 76238 |
| cg00699601 | 6 | 32762284 | rs9271082 | 2.56E-12 | 4.87E-06 | 186198 | rs9275152 | 2.19E-73 | 4.55E-40 | 110088 |
| cg12989544 | 6 | 153429875 | rs9371676 | 2.93E-07 | -3.14E-03 | -16090 | rs9397581 | 7.32E-04 | 8.82E-64 | 46285 |

| | | | | | | | | | | |
|---|---|---|---|---|---|---|---|---|---|---|
| **cg16837680** | **8** | **8084974** | **rs2921077** | **9.14E-09** | **-3.69E-04** | **-219528** | **rs2976940** | **7.88E-03** | **2.60E-31** | **-194488** |
| **cg03033453** | **8** | **8902869** | **rs7005133** | **3.02E-09** | **-3.41E-04** | **1647** | **rs6601299** | **3.23E-05** | **2.06E-12** | **-281822** |
| cg03502219 | 8 | 9978949 | rs17689007 | 5.62E-13 | -1.02E-03 | 4125 | rs28712068 | 2.77E-05 | 3.95E-34 | -272196 |
| cg01048752 | 8 | 10001523 | rs35013996 | 3.73E-08 | 4.18E-03 | 54378 | rs35437983 | 4.52E-03 | 2.45E-22 | -3530 |
| cg18477422 | 8 | 10773493 | rs4841457 | 2.84E-11 | -1.12E-02 | -18 | rs34741518 | 4.18E-05 | 3.44E-36 | -3367 |
| cg21725652 | 8 | 10918152 | rs2409685 | 4.83E-07 | 3.59E-04 | 8959 | rs56159799 | 2.12E-03 | 2.74E-06 | 596398 |
| cg18317842 | 8 | 10921502 | rs4642600 | 4.43E-08 | -9.99E-05 | -91523 | rs11986122 | 1.24E-03 | 3.79E-18 | 911553 |
| cg09697033 | 8 | 10929599 | rs11783749 | 5.87E-11 | 5.20E-04 | 127544 | rs11986122 | 1.24E-03 | 1.80E-14 | 919650 |
| cg25014118 | 8 | 10948462 | rs4841495 | 2.20E-10 | 4.77E-04 | -10362 | rs4841660 | 1.59E-03 | 4.07E-25 | -879841 |
| **cg02034205** | **8** | **11382291** | **rs2618443** | **9.82E-08** | **-9.92E-04** | **-2265** | **rs28380506** | **6.61E-03** | **9.46E-12** | **926523** |
| cg00448191 | 8 | 41504890 | rs4317621 | 1.35E-22 | -7.50E-03 | -11691 | rs4736996 | 6.55E-07 | 3.38E-05 | 11255 |
| cg20677018 | 8 | 41508391 | rs13262861 | 3.04E-26 | 8.71E-03 | -186 | rs2977864 | 6.46E-07 | 9.77E-14 | 12077 |
| cg17274126 | 8 | 41522935 | rs508419 | 5.43E-27 | 7.85E-06 | -56 | rs3802316 | 2.25E-10 | 3.24E-49 | -5293 |
| cg09232650 | 8 | 41523757 | rs508419 | 5.43E-27 | -1.08E-02 | 766 | rs13254454 | 8.49E-03 | 1.34E-46 | 22195 |
| cg16467757 | 8 | 95888145 | rs12548874 | 3.17E-11 | 2.19E-03 | -47019 | rs6991067 | 5.36E-06 | 1.68E-55 | 12469 |
| cg22283921 | 8 | 95961819 | rs10097617 | 2.44E-14 | -3.04E-03 | 193 | rs2515140 | 9.31E-05 | 2.44E-15 | 649477 |
| cg05289649 | 10 | 80925798 | rs1613299 | 2.32E-26 | 1.00E-02 | -28987 | rs35339173 | 1.58E-05 | 6.34E-238 | 20879 |
| cg21213154 | 10 | 80927984 | rs10824724 | 2.22E-10 | -1.48E-03 | 4102 | rs703994 | 3.34E-08 | 5.70E-26 | 1948 |
| cg24590165 | 10 | 94480397 | rs1832886 | 6.67E-61 | 1.72E-03 | 2858 | rs7903302 | 5.22E-18 | 2.09E-49 | 50886 |
| cg09575421 | 11 | 17415270 | rs757110 | 1.12E-25 | 3.98E-03 | -3207 | rs4148646 | 7.65E-11 | 3.19E-57 | 80 |
| cg01366692 | 11 | 65291439 | rs11601767 | 2.49E-12 | -6.53E-04 | -24943 | rs1783541 | 5.49E-03 | 9.91E-08 | -3360 |
| cg14286243 | 11 | 68867220 | rs10896415 | 1.69E-07 | 8.52E-03 | 27886 | rs3750971 | 2.19E-05 | 8.34E-189 | 36513 |
| **cg25903143** | **11** | **68920466** | **rs1060435** | **2.35E-07** | **6.11E-03** | **64871** | **rs10896402** | **2.49E-04** | **3.93E-61** | **94149** |
| **cg14036981** | **11** | **68920648** | **rs1060435** | **2.35E-07** | **8.86E-03** | **65053** | **rs10896402** | **2.49E-04** | **2.78E-115** | **94331** |
| **cg25179853** | **11** | **68924577** | **rs1060435** | **2.35E-07** | **1.08E-04** | **68982** | **rs72932523** | **1.31E-05** | **1.23E-48** | **18457** |
| **cg23740940** | **11** | **68924746** | **rs1060435** | **2.35E-07** | **6.51E-03** | **69151** | **rs61881115** | **3.38E-03** | **2.22E-16** | **-72479** |
| cg04521279 | 11 | 72424620 | rs77464186 | 9.30E-31 | -3.72E-04 | -35778 | rs11605691 | 4.44E-17 | 1.53E-09 | 5106 |
| cg08110170 | 11 | 72474129 | rs79430446 | 6.24E-24 | -2.51E-03 | 125 | rs73541184 | 5.18E-18 | 2.44E-26 | 45981 |
| cg03878208 | 11 | 72483293 | rs76550717 | 1.05E-28 | -6.66E-04 | 55121 | rs73541184 | 5.18E-18 | 4.41E-43 | 55145 |
| cg00048149 | 12 | 26472528 | rs11048458 | 3.01E-10 | 1.49E-03 | 6943 | rs10842716 | 9.17E-05 | 9.24E-35 | -26959 |
| cg07742264 | 12 | 121419635 | rs2708081 | 1.24E-09 | -2.39E-03 | -43653 | rs1169309 | 8.72E-12 | 3.41E-09 | -19557 |
| cg25150715 | 12 | 121454808 | rs1169302 | 5.37E-16 | -7.19E-04 | 22506 | rs1169302 | 2.96E-13 | 1.00E-12 | 22506 |
| **cg21805149** | **14** | **91871665** | **rs10134560** | **1.32E-08** | **1.44E-04** | **-58487** | **rs7146002** | **6.53E-04** | **9.19E-39** | **-7** |
| cg08686008 | 15 | 41876221 | rs1023193 | 2.30E-08 | -7.01E-03 | 20485 | rs28506183 | 9.32E-03 | 1.27E-75 | -310724 |
| cg01268058 | 15 | 75738663 | rs35134156 | 3.88E-11 | 1.05E-03 | -1576769 | rs11638974 | 1.53E-04 | 1.02E-39 | -60643 |
| cg23627990 | 15 | 77364890 | rs35134156 | 3.88E-11 | 6.86E-04 | 49458 | rs62008430 | 9.65E-09 | 4.85E-32 | -202936 |
| cg02931208 | 15 | 77866341 | rs12593111 | 1.80E-17 | 4.55E-03 | 12667 | rs2682922 | 6.09E-08 | 1.15E-25 | 1335 |
| cg25755215 | 15 | 90441888 | rs7174878 | 2.27E-19 | -4.27E-03 | 7000 | rs3759831 | 1.36E-07 | 1.26E-59 | 27246 |
| cg06979164 | 15 | 91530190 | rs8036430 | 2.40E-14 | 1.52E-03 | 10530 | rs12440774 | 8.53E-08 | 8.62E-73 | 47255 |
| cg01959774 | 16 | 290115 | rs6600204 | 4.95E-12 | -2.65E-03 | -10062 | rs7189326 | 2.50E-03 | 2.28E-28 | -23291 |
| cg01474600 | 16 | 315564 | rs6600204 | 4.95E-12 | -6.69E-04 | 15387 | rs7189326 | 2.50E-03 | 1.08E-150 | 2158 |

| | | | | | | | | | | |
|---|---|---|---|---|---|---|---|---|---|---|
| cg22292753 | 17 | 17655165 | rs11655029 | 2.08E-11 | -1.55E-03 | 5993 | rs117908897 | 7.86E-04 | 9.14E-48 | 70019 |
| cg22116239 | 17 | 17741676 | rs1108646 | 6.85E-11 | -2.41E-03 | -9802 | rs75035298 | 2.28E-04 | 3.97E-59 | 268662 |
| cg06518535 | 17 | 46844976 | rs35895680 | 1.08E-15 | 3.35E-04 | -215346 | rs7217007 | 1.34E-07 | 1.06E-35 | 279 |
| **cg08228953** | **19** | **7979674** | **rs3679** | **4.61E-12** | **-4.40E-03** | **1244** | **rs2303700** | **2.29E-08** | **2.55E-44** | **3145** |
| cg02942825 | 19 | 46185348 | rs112128231 | 4.49E-09 | -1.19E-03 | -3009 | rs34642101 | 3.41E-07 | 1.13E-37 | 21358 |
| cg09118261 | 20 | 32568722 | rs4911414 | 3.33E-10 | 2.28E-04 | -160722 | rs6141433 | 3.17E-04 | 7.58E-47 | 19860 |
| cg17113955 | 20 | 32610490 | rs1007090 | 1.01E-10 | 6.76E-04 | 27619 | rs6059685 | 2.51E-05 | 9.23E-41 | -121167 |
| cg17746527 | 20 | 48834262 | rs11699802 | 8.84E-12 | 4.74E-04 | 2127 | rs4253439 | 2.15E-07 | 2.88E-46 | 26251 |

[1]Weight of the SNP in the DNA methylation prediction model

[2]Distance of the SNP to the CpG: A negative value indicates that the SNP is upstream of the CpG, while a positive value indicates that the SNP is downstream of the CpG.

CpGs highlighted in bold are those in novel genes.

**Supplmentary Table S4. CpGs that were validated using samples with measured DNA methylation levels**

| CpG | Chromosome | Position | UCSC_RefGene_Name | beta | se | P-value |
|---|---|---|---|---|---|---|
| cg06697744 | 1 | 39680237 | MACF1 | 0.053 | 0.014 | 7.96E-05 |
| cg20885688 | 5 | 74956618 | ANKDD1B | 0.030 | 0.008 | 1.49E-04 |
| cg12172531 | 6 | 32184296 | NOTCH4 | -0.039 | 0.009 | 8.07E-06 |
| cg14255617 | 6 | 32729118 | HLA-DQB2 | 0.012 | 0.004 | 4.52E-04 |
| cg16467757 | 8 | 95888145 | INTS8 | -0.022 | 0.006 | 6.92E-05 |
| cg24590165 | 10 | 94480397 | HHEX(dist=24993),EXOC6(dist=106191) | -0.028 | 0.007 | 1.35E-04 |
| cg09575421 | 11 | 17415270 | ABCC8 | 0.033 | 0.008 | 1.24E-05 |
| cg01366692 | 11 | 65291439 | SCYL1 | 0.071 | 0.010 | 3.54E-12 |
| cg08110170 | 11 | 72474129 | STARD10 | -0.044 | 0.005 | 1.01E-22 |
| cg25150715 | 12 | 121454808 | C12orf43 | -0.029 | 0.007 | 1.51E-05 |

**Supplmentary Table S5. MWAS results for the 10 cardiometabolic traits**

| | FG | | | HbA1c | | | TC | | | TG | | | LDLC | | | HDLC | | | SBP | | | DBP | | | PP | | | BMI | | |
|---|---|---|---|---|---|---|---|---|---|---|---|---|---|---|---|---|---|---|---|---|---|---|---|---|---|---|---|---|---|---|
| CpG | Z score | Effect size | P_value | Z score | Effect size | P_value | Z score | Effect size | P_value | Z score | Effect size | P_value | Z score | Effect size | P_value | Z score | Effect size | P_value | Z score | Effect size | P_value | Z score | Effect size | P_value | Z score | Effect size | P_value | Z score | Effect size | P_value |
| cg03179008 | 3.243 | 0.244 | 1.18E-03 | 2.235 | 0.133 | 2.54E-02 | -3.288 | -0.165 | 1.01E-03 | 11.104 | 0.601 | 1.20E-28 | 0.958 | 0.125 | 3.38E-01 | -20.242 | -1.231 | 4.14E-91 | 3.086 | 4.242 | 2.03E-03 | 4.099 | 3.235 | 4.14E-05 | 1.130 | 1.051 | 2.58E-01 | 5.196 | 0.410 | 2.03E-07 |
| cg06697744 | -2.592 | -0.600 | 9.55E-03 | -2.966 | -0.526 | 3.02E-03 | 3.574 | 0.663 | 3.51E-04 | -9.627 | -1.840 | 6.16E-22 | 0.125 | 0.025 | 9.01E-01 | 16.576 | 3.178 | 1.04E-61 | -2.094 | -9.050 | 3.62E-02 | -3.109 | -7.839 | 1.87E-03 | -0.551 | -1.502 | 5.81E-01 | -6.201 | -1.625 | 5.62E-10 |
| cg08719237 | 2.977 | 0.329 | 2.92E-03 | 3.363 | 0.268 | 7.71E-04 | -2.241 | -0.157 | 2.50E-02 | 12.355 | 0.990 | 4.56E-35 | 1.982 | 0.248 | 4.75E-02 | -21.189 | -1.844 | 1.22E-99 | 2.679 | 5.428 | 7.38E-03 | 4.793 | 5.554 | 1.64E-06 | -0.017 | -0.013 | 9.87E-01 | 6.348 | 0.770 | 2.17E-10 |
| cg00980362 | 3.582 | 0.356 | 3.41E-04 | 2.666 | 0.186 | 7.66E-03 | -3.663 | -0.244 | 2.49E-04 | 11.761 | 0.833 | 6.23E-32 | 0.928 | 0.142 | 3.53E-01 | -21.718 | -1.684 | 1.37E-104 | 2.390 | 4.481 | 1.68E-02 | 3.941 | 4.258 | 8.13E-05 | 0.181 | 0.214 | 8.56E-01 | 6.934 | 0.783 | 4.08E-12 |
| cg21619813 | -3.047 | -0.234 | 2.31E-03 | -2.658 | -0.176 | 7.86E-03 | -3.906 | -0.277 | 9.40E-05 | -13.399 | -0.971 | 6.13E-41 | -3.908 | -0.287 | 9.30E-05 | 12.041 | 0.879 | 2.16E-33 | -4.314 | -6.858 | 1.60E-05 | 1.293 | 1.180 | 1.96E-01 | -7.161 | -7.729 | 8.01E-13 | 4.864 | 0.442 | 1.15E-06 |
| cg06162668 | 0.924 | 0.070 | 3.56E-01 | -1.037 | -0.073 | 3.00E-01 | 3.509 | 0.278 | 4.50E-04 | -4.743 | -0.385 | 2.11E-06 | 3.739 | 0.307 | 1.85E-04 | 7.117 | 0.581 | 1.10E-12 | -1.183 | -2.099 | 2.37E-01 | 0.691 | 0.707 | 4.89E-01 | -2.128 | -2.562 | 3.34E-02 | -23.369 | -2.510 | 8.88E-121 |
| cg01969012 | -10.936 | -0.608 | 7.80E-28 | -2.349 | -0.095 | 1.88E-02 | 25.899 | 1.192 | 6.78E-148 | 40.293 | 2.475 | 0.00E+00 | 11.654 | 0.554 | 2.19E-31 | -4.799 | -0.228 | 1.59E-06 | 4.137 | 4.287 | 3.51E-05 | 2.785 | 1.652 | 5.35E-03 | 3.874 | 2.728 | 1.07E-04 | -2.216 | -0.135 | 2.67E-02 |
| cg05102552 | -10.321 | -0.854 | 5.66E-25 | -2.097 | -0.128 | 3.60E-02 | 24.320 | 1.669 | 1.19E-130 | 39.540 | 3.507 | 0.00E+00 | 10.875 | 0.773 | 1.52E-27 | -4.834 | -0.341 | 1.34E-06 | 4.047 | 6.223 | 5.19E-05 | 2.746 | 2.413 | 6.04E-03 | 3.809 | 3.981 | 1.39E-04 | -1.813 | -0.167 | 6.99E-02 |
| cg22903471 | -8.350 | -0.495 | 6.81E-17 | -2.005 | -0.091 | 4.50E-02 | 18.951 | 0.922 | 4.31E-80 | 38.673 | 2.054 | 0.00E+00 | 7.395 | 0.372 | 1.41E-13 | -3.955 | -0.198 | 7.65E-05 | 4.986 | 6.307 | 6.18E-07 | 3.303 | 2.393 | 9.57E-04 | 4.898 | 4.211 | 9.66E-07 | -0.691 | -0.050 | 4.90E-01 |
| cg10337841 | -10.018 | -1.955 | 1.28E-23 | -2.402 | -0.342 | 1.63E-02 | 19.124 | 3.489 | 1.59E-81 | 33.970 | 8.425 | 6.18E-253 | 7.899 | 1.532 | 2.81E-15 | -3.333 | -1.200 | 8.60E-04 | 4.311 | 18.027 | 1.62E-05 | 3.087 | 7.337 | 2.03E-03 | 3.919 | 11.190 | 8.90E-05 | -1.360 | -0.318 | 1.74E-01 |
| cg25428544 | 3.936 | 0.468 | 8.30E-05 | 1.915 | 0.199 | 5.55E-02 | 2.668 | 0.228 | 7.62E-03 | 0.771 | 0.067 | 4.41E-01 | 3.200 | 0.284 | 1.37E-03 | -0.202 | -0.016 | 8.40E-01 | -2.474 | -5.073 | 1.34E-02 | -5.433 | -6.262 | 5.53E-08 | 0.914 | 1.137 | 3.61E-01 | -2.810 | -0.315 | 4.96E-03 |
| cg11998686 | 6.748 | 0.212 | 1.50E-11 | 5.104 | 0.128 | 3.33E-07 | 1.789 | 0.042 | 7.36E-02 | 2.432 | 0.063 | 1.50E-02 | 0.371 | 0.006 | 7.10E-01 | 0.643 | 0.019 | 5.20E-01 | 0.572 | 0.359 | 5.67E-01 | 0.333 | 0.137 | 7.39E-01 | 0.484 | 0.190 | 6.28E-01 | -2.216 | -0.083 | 2.67E-02 |
| cg05293897 | -1.693 | -0.049 | 9.04E-02 | -1.912 | -0.045 | 5.59E-02 | -0.941 | -0.004 | 3.46E-01 | -7.041 | -0.160 | 1.91E-12 | -1.975 | -0.054 | 4.83E-02 | 9.021 | 0.228 | 1.86E-19 | -2.783 | -1.378 | 5.39E-03 | -2.227 | -0.631 | 2.59E-02 | -3.131 | -1.049 | 1.74E-03 | 0.914 | -0.012 | 3.61E-01 |
| cg04891645 | -2.652 | -0.295 | 7.99E-03 | -2.994 | -0.221 | 2.76E-03 | -1.143 | -0.128 | 2.53E-01 | -20.589 | -2.319 | 3.46E-94 | -2.673 | -0.308 | 7.52E-03 | 20.860 | 2.369 | 1.23E-96 | -8.370 | -20.831 | 5.78E-17 | -7.499 | -10.707 | 6.43E-14 | -5.542 | -9.377 | 3.00E-08 | 1.764 | 0.249 | 7.77E-02 |
| cg10397322 | -2.171 | -0.384 | 2.99E-02 | -2.032 | -0.142 | 4.21E-02 | -0.959 | -0.106 | 3.37E-01 | -0.051 | -0.026 | 9.59E-01 | 0.016 | -0.009 | 9.87E-01 | -0.640 | -0.047 | 5.22E-01 | -1.488 | -3.302 | 1.37E-01 | -0.918 | -1.124 | 3.59E-01 | -1.686 | -2.608 | 9.18E-02 | 1.219 | 0.172 | 2.23E-01 |
| cg18257541 | -2.211 | -0.503 | 2.70E-02 | -2.240 | -0.229 | 2.51E-02 | -1.576 | -0.215 | 1.15E-01 | -1.277 | -0.188 | 2.02E-01 | -0.406 | -0.070 | 6.85E-01 | -0.383 | -0.031 | 7.02E-01 | -1.241 | -3.474 | 2.15E-01 | -0.527 | -0.800 | 5.98E-01 | -1.615 | -3.135 | 1.06E-01 | 1.838 | 0.308 | 6.60E-02 |
| cg10632094 | -2.377 | -0.430 | 1.75E-02 | -2.346 | -0.191 | 1.90E-02 | -1.444 | -0.157 | 1.49E-01 | -1.291 | -0.150 | 1.97E-01 | -0.362 | -0.049 | 7.17E-01 | -0.167 | -0.003 | 8.68E-01 | -1.312 | -2.895 | 1.89E-01 | -0.305 | -0.332 | 7.61E-01 | -1.916 | -2.948 | 5.54E-02 | 1.472 | 0.193 | 1.41E-01 |
| cg21007371 | -1.990 | -0.424 | 4.66E-02 | -1.324 | -0.099 | 1.85E-01 | -0.762 | -0.110 | 4.46E-01 | 0.504 | 0.041 | 6.14E-01 | 0.016 | -0.012 | 9.88E-01 | -1.179 | -0.133 | 2.39E-01 | -2.882 | -8.497 | 3.96E-03 | -2.123 | -3.547 | 3.38E-02 | -2.536 | -5.133 | 1.12E-02 | 2.004 | 0.364 | 4.51E-02 |
| cg16258503 | 3.948 | 7.973 | 7.88E-05 | 1.729 | 2.120 | 8.39E-02 | 2.671 | 4.666 | 7.56E-03 | 2.900 | 5.164 | 3.74E-03 | 2.578 | 4.650 | 9.95E-03 | -0.127 | -0.205 | 8.99E-01 | 1.931 | 249.650 | 5.35E-02 | -1.249 | -92.581 | 2.11E-01 | 3.650 | 320.379 | 2.62E-04 | 0.795 | 5.887 | 4.27E-01 |
| cg16021135 | 12.521 | 0.949 | 5.76E-36 | 7.941 | 0.460 | 2.01E-15 | 6.819 | 0.409 | 9.19E-12 | -0.567 | -0.036 | 5.71E-01 | 4.517 | 0.281 | 6.26E-06 | 6.961 | 0.431 | 3.38E-12 | -3.860 | -5.191 | 1.13E-04 | -3.384 | -2.597 | 7.14E-04 | -2.849 | -2.603 | 4.39E-03 | -2.332 | -0.188 | 1.97E-02 |
| cg12781915 | -4.044 | -0.537 | 5.26E-05 | -2.986 | -0.283 | 2.83E-03 | 2.095 | 0.229 | 3.61E-02 | -0.172 | -0.025 | 8.64E-01 | 1.123 | 0.128 | 2.61E-01 | 3.081 | 0.352 | 2.07E-03 | 1.055 | 2.649 | 2.92E-01 | 1.778 | 2.559 | 7.54E-02 | -0.239 | -0.420 | 8.11E-01 | 2.820 | 0.399 | 4.80E-03 |
| cg01899937 | 4.137 | 0.667 | 3.52E-05 | 4.093 | 0.481 | 4.26E-05 | 0.091 | -0.074 | 9.27E-01 | 1.743 | 0.216 | 8.13E-02 | -0.732 | -0.257 | 4.64E-01 | 0.281 | -0.254 | 7.79E-01 | 1.080 | 2.915 | 2.80E-01 | -0.057 | -0.049 | 9.55E-01 | 1.395 | 2.523 | 1.63E-01 | -3.289 | -0.510 | 1.01E-03 |
| cg20885688 | 0.690 | 0.047 | 4.90E-01 | -1.429 | -0.065 | 1.53E-01 | 30.805 | 1.515 | 2.23E-208 | -1.002 | -0.050 | 3.16E-01 | 30.005 | 1.529 | 8.38E-198 | 7.708 | 0.391 | 1.28E-14 | 2.532 | 2.799 | 1.13E-02 | 8.358 | 5.297 | 6.38E-17 | -3.186 | -2.385 | 1.44E-03 | -14.417 | -0.935 | 4.06E-47 |
| cg27359689 | -1.561 | -0.116 | 1.18E-01 | 1.119 | 0.059 | 2.63E-01 | -27.400 | -1.637 | 2.71E-165 | 1.641 | 0.100 | 1.01E-01 | -25.695 | -1.591 | 1.32E-145 | -9.749 | -0.601 | 1.86E-22 | -1.775 | -2.383 | 7.59E-02 | -8.739 | -6.731 | 2.36E-18 | 4.207 | 3.828 | 2.59E-05 | 12.936 | 1.013 | 2.82E-38 |
| cg17746515 | -0.177 | 0.020 | 8.59E-01 | 1.521 | 0.173 | 1.28E-01 | -2.349 | -0.235 | 1.88E-02 | 6.309 | 0.645 | 2.80E-10 | -2.448 | -0.250 | 1.43E-02 | -5.281 | -0.549 | 1.28E-07 | -0.602 | -1.737 | 5.47E-01 | 0.789 | 1.292 | 4.30E-01 | -1.406 | -2.742 | 1.60E-01 | 3.189 | 0.543 | 1.43E-03 |
| cg24959793 | 0.853 | 0.140 | 3.94E-01 | 1.725 | 0.166 | 8.46E-02 | -2.380 | -0.275 | 1.73E-02 | 5.963 | 0.256 | 2.48E-09 | -2.253 | -0.049 | 2.43E-02 | -5.231 | -0.965 | 1.69E-07 | -0.239 | -0.452 | 8.11E-01 | 1.524 | 2.185 | 1.27E-01 | -1.374 | -2.194 | 1.70E-01 | 3.618 | 0.523 | 2.97E-04 |
| cg19277069 | 3.549 | 2.380 | 3.87E-04 | -0.350 | -0.133 | 7.26E-01 | 2.603 | 1.344 | 9.25E-03 | -1.958 | -1.036 | 5.03E-02 | 2.982 | 1.598 | 2.86E-03 | 0.738 | 0.403 | 4.61E-01 | -0.405 | -4.771 | 6.86E-01 | -1.601 | -10.632 | 1.09E-01 | 1.151 | 8.953 | 2.50E-01 | -4.730 | -3.259 | 2.25E-06 |
| cg26671988 | 4.254 | 0.252 | 2.10E-05 | 0.250 | 0.010 | 8.03E-01 | 1.654 | 0.067 | 9.82E-02 | -1.293 | -0.040 | 1.96E-01 | 1.951 | 0.085 | 5.10E-02 | -0.118 | -0.021 | 9.06E-01 | -0.592 | -0.679 | 5.54E-01 | -2.609 | -1.685 | 9.07E-03 | 1.558 | 1.174 | 1.19E-01 | -6.082 | -0.392 | 1.19E-09 |
| cg22131612 | -6.867 | -0.964 | 6.56E-12 | -4.455 | -0.449 | 8.40E-06 | 2.541 | 0.261 | 1.11E-02 | -4.824 | -0.517 | 1.41E-06 | 2.352 | 0.255 | 1.87E-02 | 4.783 | 0.507 | 1.72E-06 | -5.589 | -13.115 | 2.28E-08 | -6.066 | -8.081 | 1.31E-09 | -3.197 | -5.169 | 1.39E-03 | 1.676 | 0.208 | 9.38E-02 |
| cg16175906 | -0.442 | -0.218 | 6.58E-01 | -2.521 | -0.647 | 1.17E-02 | -2.153 | -0.656 | 3.13E-02 | -1.104 | -0.483 | 2.70E-01 | -3.358 | -1.010 | 7.85E-04 | 2.474 | 0.823 | 1.34E-02 | -4.691 | -30.173 | 2.71E-06 | -2.756 | -9.953 | 5.85E-03 | -4.762 | -21.028 | 1.92E-06 | -0.926 | -0.372 | 3.54E-01 |
| cg02205391 | 0.036 | 0.656 | 9.72E-01 | -2.290 | -1.524 | 2.20E-02 | -7.228 | -5.727 | 4.90E-13 | -7.695 | -5.971 | 1.41E-14 | -6.422 | -5.285 | 1.35E-10 | 2.261 | 1.709 | 2.38E-02 | -6.177 | -108.532 | 6.53E-10 | -4.755 | -48.110 | 1.99E-06 | -5.203 | -61.825 | 1.96E-07 | -0.527 | -0.586 | 5.98E-01 |
| cg25426302 | 0.955 | 1.927 | 3.39E-01 | -5.175 | -5.930 | 2.27E-07 | 4.888 | 6.443 | 1.02E-06 | 3.539 | 4.780 | 4.02E-04 | 1.718 | 2.239 | 8.58E-02 | 5.741 | 8.059 | 9.40E-09 | 1.379 | 37.403 | 1.68E-01 | 5.747 | 93.915 | 9.06E-09 | -2.686 | -54.541 | 7.23E-03 | -4.526 | -7.993 | 6.02E-06 |
| cg12172531 | -0.684 | -0.556 | 4.94E-01 | -3.130 | -1.405 | 1.75E-03 | 3.223 | 1.596 | 1.27E-03 | -2.406 | -1.326 | 1.61E-02 | 1.142 | 0.585 | 2.54E-01 | 7.620 | 4.053 | 2.54E-14 | -2.669 | -34.378 | 7.62E-03 | 1.780 | 13.067 | 7.51E-02 | -5.255 | -45.836 | 1.48E-07 | -3.324 | -2.351 | 8.86E-04 |
| cg15710545 | 0.334 | 0.031 | 7.39E-01 | -2.671 | -0.126 | 7.57E-03 | 4.774 | 0.282 | 1.80E-06 | -4.497 | -0.209 | 6.89E-06 | 6.371 | 0.363 | 1.87E-10 | 1.148 | 0.063 | 2.51E-01 | -5.750 | -11.690 | 8.91E-09 | -3.751 | -4.282 | 1.76E-04 | -5.248 | -7.264 | 1.53E-07 | -1.486 | -0.175 | 1.37E-01 |
| cg06598146 | 0.218 | 0.053 | 8.27E-01 | 1.909 | 0.149 | 5.62E-02 | -9.769 | -0.818 | 1.53E-22 | -7.433 | -0.638 | 1.06E-13 | -9.594 | -0.825 | 8.45E-22 | 5.138 | 0.451 | 2.78E-07 | 8.929 | 43.862 | 4.29E-19 | 3.363 | 9.324 | 7.71E-04 | 10.040 | 33.619 | 1.02E-23 | 1.868 | 0.422 | 6.18E-02 |
| cg14255617 | 1.025 | 0.066 | 3.05E-01 | 0.094 | 0.017 | 9.25E-01 | 1.847 | 0.063 | 6.48E-02 | 1.787 | 0.060 | 7.40E-02 | 5.605 | 0.189 | 2.08E-08 | -9.232 | -0.310 | 2.67E-20 | 0.972 | 4.512 | 3.31E-01 | 0.372 | 0.883 | 7.10E-01 | 0.712 | 2.164 | 4.76E-01 | -1.413 | -0.132 | 1.58E-01 |
| cg00699601 | 1.590 | 0.070 | 1.12E-01 | -3.126 | -0.090 | 1.77E-03 | -1.826 | -0.052 | 6.78E-02 | -0.108 | -0.004 | 9.14E-01 | -3.252 | -0.096 | 1.15E-03 | 3.208 | 0.094 | 1.34E-03 | 2.149 | 1.384 | 3.16E-02 | 1.589 | 0.590 | 1.12E-01 | 1.639 | 0.717 | 1.01E-01 | -0.871 | -0.036 | 3.84E-01 |
| cg12989544 | 4.711 | 0.439 | 2.46E-06 | 2.127 | 0.182 | 3.34E-02 | -1.096 | -0.080 | 2.73E-01 | 7.077 | 0.518 | 1.48E-12 | -0.829 | -0.063 | 4.07E-01 | -8.585 | -0.634 | 9.12E-18 | -3.961 | -6.387 | 7.45E-05 | -6.026 | -5.556 | 1.68E-09 | -0.611 | -0.685 | 5.42E-01 | 6.408 | 0.629 | 1.47E-10 |
| cg16837680 | -0.846 | -0.078 | 3.97E-01 | 1.178 | 0.167 | 2.39E-01 | -0.434 | -0.053 | 6.64E-01 | -10.708 | -1.354 | 9.36E-27 | 3.923 | 0.502 | 8.76E-05 | -1.271 | -0.162 | 2.04E-01 | 9.156 | 26.399 | 5.40E-20 | 7.175 | 11.850 | 7.25E-13 | 7.190 | 14.068 | 6.50E-13 | 8.750 | 1.441 | 2.14E-18 |
| cg03033453 | -1.102 | -0.221 | 2.70E-01 | 1.522 | 0.395 | 1.28E-01 | 1.443 | 0.298 | 1.49E-01 | -10.344 | -2.181 | 4.46E-25 | 6.199 | 1.324 | 5.69E-10 | -1.237 | -0.266 | 2.16E-01 | 9.920 | 44.815 | 3.40E-23 | 8.463 | 21.923 | 2.61E-17 | 7.063 | 21.630 | 1.63E-12 | 9.704 | 2.690 | 2.89E-22 |
| cg03502219 | -0.087 | -0.018 | 9.30E-01 | -1.618 | -0.143 | 1.06E-01 | 1.326 | 0.098 | 1.85E-01 | 7.016 | 0.536 | 2.28E-12 | -1.708 | -0.133 | 8.76E-02 | 2.138 | 0.164 | 3.25E-02 | -7.575 | -12.682 | 3.60E-14 | -6.939 | -6.667 | 3.95E-12 | -5.335 | -6.062 | 9.57E-08 | -7.913 | -0.762 | 2.51E-15 |
| cg01048752 | -0.909 | -0.179 | 3.63E-01 | 1.021 | 0.269 | 3.07E-01 | 2.334 | 0.421 | 1.96E-02 | -8.313 | -1.522 | 9.31E-17 | 5.582 | 1.041 | 2.38E-08 | -0.074 | -0.015 | 9.41E-01 | 9.814 | 39.042 | 9.81E-23 | 8.477 | 19.347 | 2.31E-17 | 7.214 | 19.452 | 5.44E-13 | 9.630 | 2.326 | 5.97E-22 |
| cg18477422 | 2.085 | 0.129 | 3.70E-02 | -2.134 | -0.158 | 3.28E-02 | -0.834 | -0.057 | 4.04E-01 | 8.483 | 0.546 | 2.19E-17 | -5.216 | -0.345 | 1.83E-07 | 1.409 | 0.091 | 1.59E-01 | -10.249 | -14.285 | 1.19E-24 | -8.664 | -6.921 | 4.56E-18 | -7.573 | -7.167 | 3.65E-14 | -9.162 | -0.786 | 5.11E-20 |
| cg21725652 | -2.771 | -0.642 | 5.59E-03 | 1.558 | 0.420 | 1.19E-01 | 1.389 | 0.291 | 1.65E-01 | -7.046 | -1.454 | 1.84E-12 | 5.242 | 1.109 | 1.59E-07 | -1.398 | -0.277 | 1.62E-01 | 10.204 | 46.808 | 1.90E-24 | 8.402 | 22.085 | 4.38E-17 | 7.947 | 24.772 | 1.92E-15 | 8.864 | 2.432 | 7.75E-19 |
| cg18317842 | 2.651 | 0.350 | 8.02E-03 | -1.593 | -0.227 | 1.11E-01 | -0.727 | -0.093 | 4.67E-01 | 7.301 | 0.930 | 2.86E-13 | -4.163 | -0.538 | 3.14E-05 | 1.096 | 0.137 | 2.73E-01 | -7.946 | -30.819 | 1.92E-15 | -6.638 | -14.758 | 3.18E-11 | -6.218 | -16.379 | 5.04E-10 | -7.388 | -1.752 | 1.49E-13 |
| cg09697033 | -1.753 | -0.194 | 7.97E-02 | 1.488 | 0.239 | 1.37E-01 | 0.694 | 0.086 | 4.87E-01 | -8.755 | -1.119 | 2.04E-18 | 4.600 | 0.594 | 4.23E-06 | -0.619 | -0.080 | 5.36E-01 | 9.478 | 26.571 | 2.59E-21 | 7.824 | 12.571 | 5.12E-15 | 7.248 | 13.788 | 4.23E-13 | 8.459 | 1.440 | 2.69E-17 |
| cg25014118 | -2.577 | -0.063 | 9.96E-03 | 1.789 | 0.062 | 7.36E-02 | 0.048 | 0.001 | 9.62E-01 | -9.162 | -0.261 | 5.10E-20 | 3.982 | 0.115 | 6.83E-05 | -0.884 | -0.026 | 3.77E-01 | 8.821 | 5.420 | 1.13E-18 | 7.401 | 2.602 | 1.36E-13 | 6.711 | 2.800 | 1.93E-11 | 8.841 | 0.309 | 9.49E-19 |
| cg02034205 | -1.716 | -0.269 | 8.62E-02 | 2.464 | 0.466 | 1.37E-02 | 0.796 | 0.135 | 4.26E-01 | -10.429 | -1.825 | 1.83E-25 | 5.122 | 0.905 | 3.02E-07 | -0.143 | -0.025 | 8.86E-01 | 9.681 | 36.492 | 3.63E-22 | 7.288 | 15.773 | 3.15E-13 | 7.826 | 20.003 | 5.05E-15 | 9.009 | 1.928 | 2.08E-19 |
| cg00448191 | 2.407 | 0.252 | 1.61E-02 | 2.266 | 0.159 | 2.34E-02 | 1.891 | 0.137 | 5.86E-02 | -0.274 | -0.020 | 7.84E-01 | 1.402 | 0.104 | 1.61E-01 | 0.200 | 0.014 | 8.41E-01 | -1.765 | -2.928 | 7.75E-02 | -1.282 | -1.222 | 2.00E-01 | -0.968 | -1.086 | 3.33E-01 | 1.370 | 0.139 | 1.71E-01 |
| cg20677018 | 4.230 | 0.473 | 2.34E-05 | 3.019 | 0.228 | 2.54E-03 | 0.706 | 0.056 | 4.80E-01 | -1.600 | -0.128 | 1.10E-01 | 0.288 | 0.024 | 7.74E-01 | 0.748 | 0.059 | 4.54E-01 | -0.408 | -0.826 | 6.83E-01 | 0.647 | 0.815 | 5.18E-01 | -0.896 | -1.278 | 3.70E-01 | 0.791 | 0.102 | 4.29E-01 |
| cg17274126 | -3.903 | -1.213 | 9.49E-05 | -2.517 | -0.453 | 1.18E-02 | -0.266 | -0.054 | 7.90E-01 | 1.065 | 0.242 | 2.87E-01 | 0.001 | 0.002 | 9.99E-01 | -0.612 | -0.137 | 5.41E-01 | -0.288 | -1.492 | 7.73E-01 | -1.474 | -4.194 | 1.40E-01 | 0.725 | 2.369 | 4.68E-01 | -0.320 | -0.096 | 7.49E-01 |
| cg09232650 | 3.126 | 0.067 | 1.77E-03 | 2.960 | 0.036 | 3.07E-03 | 0.605 | 0.008 | 5.45E-01 | -0.996 | -0.014 | 3.19E-01 | 0.219 | 0.003 | 8.27E-01 | 1.244 | 0.018 | 2.13E-01 | -0.461 | -0.144 | 6.45E-01 | 0.038 | 0.007 | 9.69E-01 | -0.717 | -0.153 | 4.73E-01 | 0.863 | 0.017 | 3.88E-01 |
| cg16467757 | 4.629 | 0.533 | 3.68E-06 | 1.838 | 0.132 | 6.61E-02 | -2.536 | -0.209 | 1.12E-02 | 2.334 | 0.198 | 1.96E-02 | -2.289 | -0.196 | 2.21E-02 | -1.327 | -0.113 | 1.85E-01 | 5.154 | 9.540 | 2.55E-07 | 5.278 | 5.598 | 1.30E-07 | 2.862 | 3.600 | 4.22E-03 | 1.386 | 0.145 | 1.66E-01 |
| cg22283921 | -4.745 | -0.334 | 2.08E-06 | -1.834 | -0.085 | 6.66E-02 | 2.524 | 0.127 | 1.16E-02 | -2.814 | -0.145 | 4.89E-03 | 2.101 | 0.110 | 3.56E-02 | 1.807 | 0.094 | 7.08E-02 | -5.957 | -6.838 | 2.57E-09 | -5.712 | -3.756 | 1.12E-08 | -3.987 | -3.115 | 6.69E-05 | -0.308 | -0.021 | 7.58E-01 |
| cg05289649 | 0.046 | -0.007 | 9.63E-01 | 1.935 | 0.041 | 5.30E-02 | 0.047 | 0.002 | 9.62E-01 | 1.649 | 0.046 | 9.91E-02 | 0.729 | 0.020 | 4.66E-01 | -3.789 | -0.103 | 1.51E-04 | -0.220 | -0.145 | 8.26E-01 | -0.521 | -0.187 | 6.03E-01 | 0.152 | 0.052 | 8.79E-01 | 1.078 | 0.037 | 2.81E-01 |
| cg21213154 | 0.278 | 0.016 | 7.81E-01 | 0.348 | 0.009 | 7.28E-01 | 0.768 | 0.073 | 4.42E-01 | 1.548 | 0.152 | 1.22E-01 | 1.324 | 0.133 | 1.85E-01 | -3.387 | -0.336 | 7.06E-04 | -0.693 | -1.584 | 4.88E-01 | -0.407 | -0.536 | 6.84E-01 | -0.766 | -1.186 | 4.44E-01 | 1.533 | 0.217 | 1.25E-01 |
| cg24590165 | 2.451 | 0.201 | 1.43E-02 | 2.819 | 0.160 | 4.82E-03 | 0.145 | 0.016 | 8.85E-01 | 4.460 | 0.250 | 8.21E-06 | 0.211 | 0.021 | 8.33E-01 | -3.336 | -0.179 | 8.50E-04 | 2.051 | 2.505 | 4.03E-02 | 0.024 | 0.017 | 9.80E-01 | 2.895 | 2.403 | 3.79E-03 | -1.641 | -0.117 | 1.01E-01 |
| cg09575421 | 1.007 | 0.191 | 3.14E-01 | 2.243 | 0.307 | 2.49E-02 | 2.091 | 0.257 | 3.65E-02 | 0.832 | 0.105 | 4.05E-01 | 1.247 | 0.159 | 2.12E-01 | 1.619 | 0.205 | 1.06E-01 | 10.724 | 29.766 | 7.86E-27 | 7.366 | 11.727 | 1.76E-13 | 9.228 | 17.405 | 2.76E-20 | -5.800 | -0.885 | 6.63E-09 |
| cg01366692 | 0.645 | 0.399 | 5.19E-01 | 1.047 | 0.220 | 2.95E-01 | 0.747 | 0.197 | 4.55E-01 | 2.466 | 0.659 | 1.37E-02 | 0.363 | 0.099 | 7.17E-01 | -0.041 | -0.008 | 9.68E-01 | 1.949 | 12.205 | 5.13E-02 | -2.058 | -7.292 | 3.96E-02 | 4.422 | 18.735 | 9.76E-06 | 4.600 | 1.682 | 4.22E-06 |
| cg14286243 | -2.933 | -0.150 | 3.35E-03 | -2.293 | -0.071 | 2.18E-02 | -0.054 | -0.002 | 9.57E-01 | -2.417 | -0.094 | 1.57E-02 | 0.915 | 0.036 | 3.60E-01 | 0.045 | 0.003 | 9.64E-01 | -1.381 | -1.190 | 1.67E-01 | 0.600 | 0.307 | 5.49E-01 | -2.677 | -1.578 | 7.43E-03 | 0.156 | 0.011 | 8.76E-01 |
| cg25903143 | -2.596 | -0.296 | 9.43E-03 | -1.721 | -0.112 | 8.52E-02 | 0.014 | 0.035 | 9.89E-01 | -3.156 | -0.272 | 1.60E-03 | 0.596 | 0.100 | 5.51E-01 | 1.725 | 0.146 | 8.46E-02 | -1.049 | -2.151 | 2.94E-01 | 1.388 | 1.621 | 1.65E-01 | -2.786 | -3.867 | 5.33E-03 | -1.604 | -0.178 | 1.09E-01 |
| cg14036981 | -2.345 | -0.095 | 1.90E-02 | -1.824 | -0.037 | 6.82E-02 | 0.291 | 0.009 | 7.71E-01 | -3.584 | -0.108 | 3.39E-04 | 1.244 | 0.038 | 2.13E-01 | 1.238 | 0.038 | 2.16E-01 | -0.826 | -0.607 | 4.09E-01 | 1.713 | 0.752 | 8.66E-02 | -2.737 | -1.398 | 6.19E-03 | -0.880 | -0.040 | 3.79E-01 |
| cg25179853 | -3.863 | -0.411 | 1.12E-04 | -0.793 | -0.046 | 4.28E-01 | 0.133 | 0.012 | 8.94E-01 | -1.008 | -0.079 | 3.14E-01 | 1.334 | 0.107 | 1.82E-01 | -1.281 | -0.094 | 2.00E-01 | -0.566 | -1.091 | 5.71E-01 | 2.058 | 2.328 | 3.96E-02 | -2.677 | -3.563 | 7.44E-03 | 0.766 | 0.093 | 4.43E-01 |
| cg23740940 | -3.501 | -0.086 | 4.63E-04 | -0.788 | -0.006 | 4.31E-01 | 0.153 | 0.003 | 8.78E-01 | -1.169 | -0.022 | 2.42E-01 | 1.261 | 0.024 | 2.07E-01 | -1.537 | -0.028 | 1.24E-01 | -0.650 | -0.282 | 5.15E-01 | 2.522 | 0.627 | 1.17E-02 | -3.132 | -0.922 | 1.74E-03 | -0.032 | 0.000 | 9.74E-01 |
| cg04521279 | -8.191 | -2.028 | 2.59E-16 | -3.938 | -0.771 | 8.21E-05 | -0.090 | -0.017 | 9.28E-01 | 0.828 | 0.164 | 4.08E-01 | 0.436 | 0.091 | 6.63E-01 | -2.094 | -0.423 | 3.63E-02 | 3.075 | 13.720 | 2.11E-03 | 2.582 | 6.609 | 9.83E-03 | 2.331 | 7.070 | 1.98E-02 | 4.014 | 1.088 | 5.97E-05 |
| cg08110170 | -9.880 | -3.141 | 5.09E-23 | -4.749 | -1.123 | 2.05E-06 | -0.521 | -0.134 | 6.02E-01 | 1.723 | 0.453 | 8.49E-02 | -0.032 | -0.008 | 9.74E-01 | -3.328 | -0.882 | 8.74E-04 | 1.683 | 9.785 | 9.23E-02 | 1.240 | 4.141 | 2.15E-01 | 1.545 | 6.101 | 1.22E-01 | 4.383 | 1.520 | 1.17E-05 |
| cg03878208 | -9.413 | -1.373 | 4.83E-21 | -4.385 | -0.482 | 1.16E-05 | 0.049 | 0.004 | 9.61E-01 | 1.517 | 0.186 | 1.29E-01 | 0.505 | 0.061 | 6.13E-01 | -3.162 | -0.390 | 1.57E-03 | 2.237 | 6.593 | 2.53E-02 | 1.603 | 2.697 | 1.09E-01 | 2.059 | 4.146 | 3.94E-02 | 3.683 | 0.679 | 2.31E-04 |
| cg00048149 | -1.920 | -0.598 | 5.48E-02 | -0.782 | -0.275 | 4.34E-01 | 0.710 | 0.222 | 4.77E-01 | -8.813 | -2.974 | 1.22E-18 | 1.614 | 0.539 | 1.07E-01 | 7.860 | 2.666 | 3.86E-15 | -7.400 | -54.620 | 1.37E-13 | -7.166 | -30.319 | 7.72E-13 | -4.769 | -24.128 | 1.85E-06 | 3.188 | 1.357 | 1.43E-03 |
| cg07742264 | -1.143 | -0.440 | 2.53E-01 | -1.393 | -0.409 | 1.64E-01 | -13.773 | -4.308 | 3.71E-43 | -4.023 | -1.263 | 5.75E-05 | -12.945 | -4.183 | 2.49E-38 | -1.546 | -0.562 | 1.22E-01 | 1.746 | 12.575 | 8.08E-02 | -0.065 | -0.494 | 9.48E-01 | 2.518 | 12.550 | 1.18E-02 | -2.348 | -0.972 | 1.89E-02 |
| cg25150715 | 2.923 | 0.654 | 3.47E-03 | -0.169 | 0.006 | 8.66E-01 | 19.627 | 3.552 | 9.14E-86 | 0.416 | 0.075 | 6.77E-01 | 18.878 | 3.548 | 1.74E-79 | 7.514 | 1.409 | 5.73E-14 | 3.783 | 15.336 | 1.55E-04 | 5.094 | 11.883 | 3.51E-07 | 1.402 | 3.903 | 1.61E-01 | -1.070 | -0.266 | 2.85E-01 |
| cg21805149 | -3.692 | -0.753 | 2.23E-04 | -0.530 | -0.174 | 5.96E-01 | 0.978 | 0.169 | 3.28E-01 | -2.288 | -0.408 | 2.21E-02 | 0.499 | 0.090 | 6.18E-01 | 1.556 | 0.278 | 1.20E-01 | 0.319 | 1.229 | 7.50E-01 | 1.768 | 3.967 | 7.71E-02 | -1.160 | -3.085 | 2.46E-01 | -2.663 | -0.623 | 7.73E-03 |
| cg08686008 | 1.270 | 0.019 | 2.04E-01 | 1.259 | 0.024 | 2.08E-01 | 0.432 | 0.007 | 6.66E-01 | 6.877 | 0.110 | 6.09E-12 | 1.333 | 0.021 | 1.83E-01 | -7.020 | -0.113 | 2.22E-12 | 2.541 | 0.918 | 1.11E-02 | 3.898 | 0.813 | 9.68E-05 | 0.613 | 0.151 | 5.40E-01 | 2.165 | 0.048 | 3.04E-02 |
| cg01268058 | 2.160 | 0.170 | 3.08E-02 | 3.193 | 0.220 | 1.41E-03 | 2.266 | 0.146 | 2.35E-02 | 0.452 | 0.030 | 6.51E-01 | 2.300 | 0.154 | 2.14E-02 | -1.129 | -0.076 | 2.59E-01 | 0.790 | 1.038 | 4.30E-01 | -0.414 | -0.399 | 6.79E-01 | 1.691 | 1.606 | 9.08E-02 | 0.707 | 0.060 | 4.80E-01 |
| cg23627990 | -2.456 | -0.212 | 1.41E-02 | -2.311 | -0.145 | 2.08E-02 | -3.667 | -0.189 | 2.45E-04 | -3.017 | -0.159 | 2.55E-03 | -2.771 | -0.149 | 5.59E-03 | 0.178 | 0.009 | 8.59E-01 | -0.881 | -1.030 | 3.78E-01 | -2.828 | -1.892 | 4.69E-03 | 1.294 | 1.006 | 1.96E-01 | -4.236 | -0.301 | 2.28E-05 |
| cg02931208 | 2.996 | 0.619 | 2.74E-03 | 2.560 | 0.434 | 1.05E-02 | -0.325 | -0.045 | 7.46E-01 | -0.416 | -0.058 | 6.78E-01 | -0.685 | -0.098 | 4.94E-01 | 0.345 | 0.049 | 7.30E-01 | -0.090 | -0.296 | 9.28E-01 | -0.051 | -0.102 | 9.59E-01 | -0.156 | -0.346 | 8.76E-01 | 2.874 | 0.539 | 4.06E-03 |
| cg25755215 | -1.599 | -0.106 | 1.10E-01 | -1.390 | -0.050 | 1.64E-01 | -2.330 | -0.110 | 1.98E-02 | -0.896 | -0.045 | 3.70E-01 | -1.658 | -0.082 | 9.73E-02 | -0.700 | -0.031 | 4.84E-01 | -0.603 | -0.779 | 5.46E-01 | 0.485 | 0.350 | 6.28E-01 | -1.049 | -0.920 | 2.94E-01 | 1.270 | 0.100 | 2.04E-01 |
| cg06979164 | -3.211 | -0.167 | 1.32E-03 | -2.198 | -0.067 | 2.79E-02 | 3.098 | 0.131 | 1.95E-03 | -0.117 | -0.005 | 9.07E-01 | 3.156 | 0.138 | 1.60E-03 | 2.517 | 0.109 | 1.18E-02 | 1.778 | 1.727 | 7.54E-02 | 1.833 | 1.023 | 6.68E-02 | 0.843 | 0.563 | 3.99E-01 | -0.720 | -0.040 | 4.71E-01 |
| cg01959774 | -3.518 | -0.505 | 4.34E-04 | -4.791 | -0.494 | 1.66E-06 | -2.592 | -0.258 | 9.53E-03 | -3.190 | -0.325 | 1.42E-03 | -1.578 | -0.165 | 1.14E-01 | 2.217 | 0.228 | 2.67E-02 | -0.977 | -2.213 | 3.29E-01 | -2.054 | -2.613 | 4.00E-02 | 0.117 | 0.106 | 9.07E-01 | -1.312 | -0.177 | 1.90E-01 |
| cg01474600 | -4.042 | -0.737 | 5.31E-05 | -4.391 | -0.610 | 1.13E-05 | -2.579 | -0.348 | 9.90E-03 | -2.972 | -0.414 | 2.96E-03 | -1.702 | -0.241 | 8.88E-02 | 2.069 | 0.290 | 3.86E-02 | -0.666 | -2.000 | 5.05E-01 | -2.048 | -3.590 | 4.06E-02 | 0.552 | 1.171 | 5.81E-01 | -0.238 | -0.044 | 8.12E-01 |
| cg22292753 | -0.459 | -0.264 | 6.46E-01 | -4.793 | -1.028 | 1.64E-06 | -4.944 | -1.146 | 7.67E-07 | -4.706 | -1.127 | 2.52E-06 | -4.625 | -1.113 | 3.74E-06 | 2.106 | 0.511 | 3.52E-02 | -2.026 | -10.512 | 4.28E-02 | -2.252 | -6.743 | 2.43E-02 | -1.347 | -4.774 | 1.78E-01 | 2.477 | 0.768 | 1.32E-02 |
| cg22116239 | -0.451 | -0.162 | 6.52E-01 | -4.711 | -0.597 | 2.47E-06 | -3.182 | -0.410 | 1.46E-03 | -5.012 | -0.675 | 5.38E-07 | -2.938 | -0.392 | 3.31E-03 | 2.899 | 0.396 | 3.75E-03 | -1.770 | -5.706 | 7.67E-02 | -1.856 | -3.412 | 6.34E-02 | -1.240 | -2.752 | 2.15E-01 | 2.108 | 0.383 | 3.50E-02 |
| cg06518535 | 1.528 | 0.347 | 1.27E-01 | 3.101 | 0.385 | 1.93E-03 | 3.187 | 0.451 | 1.44E-03 | 4.060 | 0.583 | 4.90E-05 | 3.886 | 0.572 | 1.02E-04 | -5.492 | -0.796 | 3.98E-08 | 5.641 | 17.761 | 1.69E-08 | 4.586 | 8.322 | 4.53E-06 | 4.138 | 8.883 | 3.50E-05 | 6.225 | 1.115 | 4.82E-10 |
| cg08228953 | -2.090 | -0.263 | 3.66E-02 | -2.031 | -0.220 | 4.23E-02 | -0.968 | -0.092 | 3.33E-01 | -10.969 | -1.040 | 5.38E-28 | -0.760 | -0.076 | 4.47E-01 | 8.408 | 0.800 | 4.18E-17 | -1.081 | -2.405 | 2.80E-01 | 0.275 | 0.271 | 7.83E-01 | -1.761 | -2.598 | 7.83E-02 | -2.451 | -0.295 | 1.43E-02 |
| cg02942825 | 2.378 | 0.315 | 1.74E-02 | 1.482 | 0.100 | 1.38E-01 | -2.573 | -0.269 | 1.01E-02 | 2.823 | 0.298 | 4.76E-03 | -1.906 | -0.208 | 5.66E-02 | -4.710 | -0.503 | 2.47E-06 | -1.800 | -4.135 | 7.19E-02 | -0.202 | -0.283 | 8.40E-01 | -2.140 | -3.330 | 3.23E-02 | 4.306 | 0.597 | 1.66E-05 |
| cg09118261 | -2.248 | -0.340 | 2.46E-02 | 0.565 | 0.032 | 5.72E-01 | 0.781 | 0.095 | 4.35E-01 | -7.798 | -0.961 | 6.30E-15 | 0.658 | 0.083 | 5.10E-01 | 8.534 | 1.060 | 1.41E-17 | -0.020 | -0.055 | 9.84E-01 | 0.877 | 1.382 | 3.80E-01 | -0.509 | -0.949 | 6.11E-01 | -6.522 | -1.050 | 6.95E-11 |
| cg17113955 | -3.016 | -0.315 | 2.56E-03 | 0.347 | 0.008 | 7.29E-01 | 1.043 | 0.093 | 2.97E-01 | -7.701 | -0.707 | 1.36E-14 | 1.159 | 0.111 | 2.47E-01 | 8.506 | 0.797 | 1.80E-17 | -0.083 | -0.165 | 9.34E-01 | 1.760 | 2.053 | 7.85E-02 | -1.390 | -1.913 | 1.65E-01 | -5.428 | -0.639 | 5.69E-08 |
| cg17746527 | 3.117 | 0.544 | 1.83E-03 | 0.542 | 0.020 | 5.88E-01 | 3.607 | 0.460 | 3.10E-04 | -2.022 | -0.268 | 4.32E-02 | 3.889 | 0.515 | 1.00E-04 | 3.168 | 0.421 | 1.53E-03 | -0.508 | -1.534 | 6.12E-01 | -1.585 | -2.674 | 1.13E-01 | 0.492 | 0.933 | 6.23E-01 | -0.618 | -0.112 | 5.37E-01 |

**Supplmentary Table S6. eQTM statistics for CpGs and the genes they regulate in alpha cells**

| CpG | | | | Regulated Gene | | | | meQTL Statistics | | Differential expression P-value |
|---|---|---|---|---|---|---|---|---|---|---|
| **CpG** | **Chromosome** | **Position** | **Gene** | **Gene** | **Chromosome** | **START** | **END** | **beta** | **p-value** | **p-value** |
| cg01969012 | 2 | 27650506 | NRBP1 | KRTCAP3 | 2 | 27666921 | 27666923 | 0.294 | 1.05E-01 | 2.20E-01 |
| cg11998686 | 2 | 43687897 | THADA | THADA | 2 | 43783473 | 43783475 | 0.320 | 9.26E-02 | 5.70E-01 |
| cg10632094 | 3 | 23244068 | UBE2E2 | UBE2E2 | 3 | 23267264 | 23267266 | 0.298 | 1.12E-01 | 7.04E-01 |
| cg21007371 | 3 | 23244130 | UBE2E2 | UBE2E2 | 3 | 23267264 | 23267266 | -0.279 | 1.50E-01 | 7.04E-01 |
| cg25426302 | 6 | 32120826 | PPT2;PRRT1 | AGPAT1 | 6 | 32137053 | 32137055 | 0.313 | 8.26E-02 | 1.12E-01 |
| cg25426302 | 6 | 32120826 | PPT2;PRRT1 | NOTCH4 | 6 | 32163214 | 32163216 | 0.271 | 1.36E-01 | 1.92E-01 |
| cg06598146 | 6 | 32711632 | HLA-DQA2 | TAP2 | 6 | 32796683 | 32796685 | 0.379 | 3.77E-02 | 5.50E-03 |
| cg03033453 | 8 | 8902869 | ERI1 | PPP1R3B | 8 | 8998304 | 8998306 | -0.406 | 2.34E-02 | 1.21E-02 |
| cg00448191 | 8 | 41504890 | NKX6-3 | ANK1 | 8 | 41513246 | 41513248 | 0.273 | 1.27E-01 | 8.83E-02 |
| cg16467757 | 8 | 95888145 | INTS8 | TP53INP1 | 8 | 95944314 | 95944316 | 0.258 | 1.49E-01 | 7.96E-01 |
| cg01366692 | 11 | 65291439 | SCYL1 | FAM89B | 11 | 65341110 | 65341112 | -0.257 | 1.90E-01 | 1.75E-01 |
| cg14286243 | 11 | 68867220 | TPCN2 - LOC338694 | LOC338694 | 11 | 68914696 | 68915208 | 0.275 | 1.26E-01 | 5.28E-01 |
| cg04521279 | 11 | 72424620 | ARAP1 | STARD10 | 11 | 72465942 | 72465944 | 0.246 | 1.80E-01 | 7.48E-02 |
| cg03878208 | 11 | 72483293 | STARD10 | FCHSD2 | 11 | 72549836 | 72549838 | -0.353 | 6.53E-02 | 7.54E-02 |
| cg07742264 | 12 | 121419635 | HNF1A | OASL | 12 | 121458364 | 121458366 | -0.320 | 1.10E-01 | 2.20E-03 |
| cg07742264 | 12 | 121419635 | HNF1A | HNF1A | 12 | 121438993 | 121438995 | -0.258 | 1.97E-01 | 1.54E-01 |
| cg25150715 | 12 | 121454808 | C12orf4 | OASL | 12 | 121458364 | 121458366 | -0.368 | 4.22E-02 | 2.20E-03 |
| cg01268058 | 15 | 75738663 | SIN3A | PTPN9 | 15 | 75761110 | 75761112 | 0.400 | 2.59E-02 | 2.92E-01 |
| cg06979164 | 15 | 91530190 | PRC1-AS1;PRC1 | VPS33B | 15 | 91542205 | 91542207 | -0.326 | 8.84E-02 | 4.89E-01 |
| cg01474600 | 16 | 315564 | FAM234A | AXIN1 | 16 | 338122 | 338124 | 0.241 | 1.98E-01 | 2.87E-01 |
| cg22292753 | 17 | 17655165 | RAI1 | TOM1L2 | 17 | 17750949 | 17750951 | -0.373 | 3.88E-02 | 5.98E-01 |
| cg08228953 | 19 | 7979674 | TGFBR3L | SNAPC2 | 19 | 7987647 | 7987649 | 0.245 | 1.73E-01 | 6.02E-02 |
| cg02942825 | 19 | 46185348 | GIPR | MEIOSIN | 19 | 46267374 | 46267376 | -0.262 | 1.47E-01 | 9.83E-01 |
| cg17113955 | 20 | 32610490 | RALY | RALY | 20 | 32666353 | 32666355 | -0.259 | 1.63E-01 | 5.83E-02 |
| cg17746527 | 20 | 48834262 | CEBPB - PELATON | LINC01271 | 20 | 48927248 | 48927786 | 0.333 | 6.79E-02 | 1.09E-01 |
| cg17746527 | 20 | 48834262 | CEBPB - PELATON | LINC01270 | 20 | 48929837 | 48931456 | 0.244 | 1.83E-01 | 2.40E-01 |

**Supplmentary Table S7. eQTM statistics for CpGs and the genes they regulate in beta cells**

| CpG | | | | Regulated Gene | | | | meQTL Statistics | | Differential expression P-value |
|---|---|---|---|---|---|---|---|---|---|---|
| CpG | Chromosome | Position | Gene | Gene | Chromosome | START | END | beta | p-value | p-value |
| cg01969012 | 2 | 27650506 | NRBP1 | KRTCAP3 | 2 | 27666921 | 27666923 | 0.568 | 2.42E-02 | 8.61E-01 |
| cg01969012 | 2 | 27650506 | NRBP1 | FNDC4 | 2 | 27715310 | 27715312 | 0.386 | 1.25E-01 | 2.76E-01 |
| cg22903471 | 2 | 27725779 | GCKR | GCKR | 2 | 27746304 | 27746306 | -0.539 | 3.11E-02 | 1.44E-01 |
| cg22903471 | 2 | 27725779 | GCKR | C2orf16 | 2 | 27805392 | 27805394 | 0.449 | 7.25E-02 | 3.39E-02 |
| cg21007371 | 3 | 23244130 | UBE2E2 | UBE2E2 | 3 | 23267264 | 23267266 | 0.442 | 8.33E-02 | 3.70E-03 |
| cg16021135 | 3 | 123130669 | ADCY5 | HACD2 | 3 | 123213735 | 123213737 | -0.378 | 1.50E-01 | 4.83E-01 |
| cg20885688 | 5 | 74956618 | ANKDD1B | LOC441087 | 5 | 75025889 | 75026795 | -0.491 | 6.57E-02 | 5.41E-01 |
| cg14255617 | 6 | 32729118 | HLA-DQB2 | TAP2 | 6 | 32796683 | 32796685 | 0.449 | 7.25E-02 | 2.95E-02 |
| cg03033453 | 8 | 8902869 | ERI1 | PPP1R3B | 8 | 8998304 | 8998306 | -0.454 | 7.81E-02 | 1.40E-03 |
| cg03033453 | 8 | 8902869 | ERI1 | SNORD3I | 8 | 8916049 | 8916140 | -0.386 | 1.36E-01 | 7.37E-02 |
| cg02034205 | 8 | 11382291 | BLK | LINC00208 | 8 | 11437979 | 11438850 | 0.381 | 1.54E-01 | 2.33E-01 |
| cg01474600 | 16 | 315564 | FAM234A | ARHGDIG | 16 | 332812 | 332814 | -0.360 | 1.51E-01 | 2.83E-01 |
| cg01474600 | 16 | 315564 | FAM234A | AXIN1 | 16 | 338122 | 338124 | -0.360 | 1.51E-01 | 1.82E-01 |
| cg22292753 | 17 | 17655165 | RAI1 | MIR33B | 17 | 17717211 | 17717230 | -0.414 | 1.03E-01 | 4.96E-01 |
| cg08228953 | 19 | 7979674 | TGFBR3L | CTXN1 | 19 | 7990175 | 7990177 | 0.449 | 7.25E-02 | 1.12E-01 |
| cg02942825 | 19 | 46185348 | GIPR | QPCTL | 19 | 46206305 | 46206307 | -0.360 | 1.51E-01 | 5.54E-01 |

**Supplmentary Table S8. Pathway analysis results for genes associated with CpGs in alpha cells**

| **gene_set_id** | **gene_set_name** | **p-value** | **FDR** |
|---|---|---|---|
| REACTOME:R-HSA-168898 | Toll-Like Receptors Cascades | 0.0002 | 0.0918 |
| REACTOME:R-HSA-8873719 | RAB geranylgeranylation | 0.0003 | 0.0918 |
| REACTOME:R-HSA-180024 | DARPP-32 events | 0.0004 | 0.0918 |
| KEGG_PARKINSONS_DISEASE | Parkinson's disease | 0.0004 | 0.0918 |
| KEGG_PPAR_SIGNALING_PATHWAY | Ppar signaling pathway | 0.0004 | 0.0918 |
| KEGG_ACUTE_MYELOID_LEUKEMIA | Acute myeloid leukemia | 0.0005 | 0.0918 |
| REACTOME:R-HSA-166016 | Toll Like Receptor 4 (TLR4) Cascade | 0.0007 | 0.1075 |
| REACTOME:R-HSA-1433557 | Signaling by SCF-KIT | 0.0009 | 0.1242 |
| REACTOME:R-HSA-70263 | Gluconeogenesis | 0.0016 | 0.1461 |
| REACTOME:R-HSA-2644603 | Signaling by NOTCH1 in Cancer | 0.0019 | 0.1461 |
| REACTOME:R-HSA-2644606 | Constitutive Signaling by NOTCH1 PEST Domain Mutants | 0.0019 | 0.1461 |
| REACTOME:R-HSA-2894858 | Signaling by NOTCH1 HD+PEST Domain Mutants in Cancer | 0.0019 | 0.1461 |
| REACTOME:R-HSA-2894862 | Constitutive Signaling by NOTCH1 HD+PEST Domain Mutants | 0.0019 | 0.1461 |
| REACTOME:R-HSA-2644602 | Signaling by NOTCH1 PEST Domain Mutants in Cancer | 0.0019 | 0.1461 |
| REACTOME:R-HSA-168181 | Toll Like Receptor 7/8 (TLR7/8) Cascade | 0.0020 | 0.1461 |
| REACTOME:R-HSA-975155 | MyD88 dependent cascade initiated on endosome | 0.0020 | 0.1461 |
| KEGG_INSULIN_SIGNALING_PATHWAY | Insulin signaling pathway | 0.0022 | 0.1461 |
| REACTOME:R-HSA-168188 | Toll Like Receptor TLR6:TLR2 Cascade | 0.0025 | 0.1491 |
| REACTOME:R-HSA-166058 | MyD88:Mal cascade initiated on plasma membrane | 0.0025 | 0.1491 |
| KEGG_MAPK_SIGNALING_PATHWAY | Mapk signaling pathway | 0.0028 | 0.1491 |
| REACTOME:R-HSA-2173796 | SMAD2/SMAD3:SMAD4 heterotrimer regulates transcription | 0.0029 | 0.1491 |
| REACTOME:R-HSA-381753 | Olfactory Signaling Pathway | 0.0029 | 0.1491 |
| KEGG_OXIDATIVE_PHOSPHORYLATION | Oxidative phosphorylation | 0.0030 | 0.1491 |
| REACTOME:R-HSA-416700 | Other semaphorin interactions | 0.0032 | 0.1535 |
| REACTOME:R-HSA-975138 | TRAF6 mediated induction of NFkB and MAP kinases upon TLR7/8 or 9 activatio | 0.0037 | 0.1618 |
| REACTOME:R-HSA-9007101 | Rab regulation of trafficking | 0.0039 | 0.1618 |
| KEGG_HUNTINGTONS_DISEASE | Huntington's disease | 0.0041 | 0.1618 |
| KEGG_OLFACTORY_TRANSDUCTION | Olfactory transduction | 0.0042 | 0.1618 |
| REACTOME:R-HSA-8854214 | TBC/RABGAPs | 0.0046 | 0.1618 |
| REACTOME:R-HSA-168138 | Toll Like Receptor 9 (TLR9) Cascade | 0.0049 | 0.1618 |
| REACTOME:R-HSA-186763 | Downstream signal transduction | 0.0049 | 0.1618 |
| KEGG_PRION_DISEASES | Prion diseases | 0.0051 | 0.1618 |
| REACTOME:R-HSA-168179 | Toll Like Receptor TLR1:TLR2 Cascade | 0.0051 | 0.1618 |
| REACTOME:R-HSA-181438 | Toll Like Receptor 2 (TLR2) Cascade | 0.0051 | 0.1618 |

| | | | |
|---|---|---|---|
| REACTOME:R-HSA-1369062 | ABC transporters in lipid homeostasis | 0.0053 | 0.1618 |
| REACTOME:R-HSA-168142 | Toll Like Receptor 10 (TLR10) Cascade | 0.0054 | 0.1618 |
| REACTOME:R-HSA-168176 | Toll Like Receptor 5 (TLR5) Cascade | 0.0054 | 0.1618 |
| REACTOME:R-HSA-975871 | MyD88 cascade initiated on plasma membrane | 0.0054 | 0.1618 |
| REACTOME:R-HSA-166520 | Signalling by NGF | 0.0056 | 0.1650 |
| REACTOME:R-HSA-2408517 | SeMet incorporation into proteins | 0.0058 | 0.1663 |
| REACTOME:R-HSA-5663202 | Diseases of signal transduction | 0.0059 | 0.1663 |
| REACTOME:R-HSA-450294 | MAP kinase activation in TLR cascade | 0.0064 | 0.1672 |
| REACTOME:R-HSA-186712 | Regulation of beta-cell development | 0.0064 | 0.1672 |
| REACTOME:R-HSA-448424 | Interleukin-17 signaling | 0.0065 | 0.1672 |
| REACTOME:R-HSA-8854691 | Interleukin-20 family signaling | 0.0067 | 0.1672 |
| REACTOME:R-HSA-6799198 | Complex I biogenesis | 0.0067 | 0.1672 |
| REACTOME:R-HSA-388844 | Receptor-type tyrosine-protein phosphatases | 0.0069 | 0.1677 |
| REACTOME:R-HSA-168638 | NOD1/2 Signaling Pathway | 0.0073 | 0.1733 |
| KEGG_TOLL_LIKE_RECEPTOR_SIGNALING_PATHWAY | Toll-like receptor signaling pathway | 0.0074 | 0.1733 |
| REACTOME:R-HSA-168164 | Toll Like Receptor 3 (TLR3) Cascade | 0.0079 | 0.1814 |
| REACTOME:R-HSA-187037 | NGF signalling via TRKA from the plasma membrane | 0.0089 | 0.1994 |

**Supplmentary Table S9. Pathway analysis results for genes associated with CpGs in beta cells**

| gene_set_id | gene_set_name | p-value | FDR |
|---|---|---|---|
| REACTOME:R-HSA-112399 | IRS-mediated signalling | 0.0751 | 0.1036 |
| REACTOME:R-HSA-5607764 | CLEC7A (Dectin-1) signaling | 0.0751 | 0.1036 |
| REACTOME:R-HSA-1168372 | Downstream signaling events of B Cell Receptor (BCR) | 0.0751 | 0.1036 |
| REACTOME:R-HSA-109704 | PI3K Cascade | 0.0751 | 0.1036 |
| REACTOME:R-HSA-8941858 | Regulation of RUNX3 expression and activity | 0.0751 | 0.1036 |
| REACTOME:R-HSA-195253 | Degradation of beta-catenin by the destruction complex | 0.0751 | 0.1036 |
| REACTOME:R-HSA-8939902 | Regulation of RUNX2 expression and activity | 0.0751 | 0.1036 |
| REACTOME:R-HSA-1169091 | Activation of NF-kappaB in B cells | 0.0751 | 0.1036 |
| REACTOME:R-HSA-8856825 | Cargo recognition for clathrin-mediated endocytosis | 0.0751 | 0.1036 |
| REACTOME:R-HSA-9013973 | TICAM1-dependent activation of IRF3/IRF7 | 0.0751 | 0.1036 |
| REACTOME:R-HSA-2428924 | IGF1R signaling cascade | 0.0751 | 0.1036 |
| REACTOME:R-HSA-8949613 | Cristae formation | 0.0751 | 0.1036 |
| REACTOME:R-HSA-180585 | Vif-mediated degradation of APOBEC3G | 0.0751 | 0.1036 |
| REACTOME:R-HSA-2404192 | Signaling by Type 1 Insulin-like Growth Factor 1 Receptor (IGF1R) | 0.0751 | 0.1036 |
| REACTOME:R-HSA-2428928 | IRS-related events triggered by IGF1R | 0.0751 | 0.1036 |
| REACTOME:R-HSA-450531 | Regulation of mRNA stability by proteins that bind AU-rich elements | 0.0751 | 0.1036 |
| REACTOME:R-HSA-8957275 | Post-translational protein phosphorylation | 0.0751 | 0.1036 |
| REACTOME:R-HSA-5607761 | Dectin-1 mediated noncanonical NF-kB signaling | 0.0751 | 0.1036 |
| REACTOME:R-HSA-5676590 | NIK-->noncanonical NF-kB signaling | 0.0751 | 0.1036 |
| REACTOME:R-HSA-4608870 | Asymmetric localization of PCP proteins | 0.0751 | 0.1036 |
| REACTOME:R-HSA-8948751 | Regulation of PTEN stability and activity | 0.0751 | 0.1036 |
| REACTOME:R-HSA-74751 | Insulin receptor signalling cascade | 0.0751 | 0.1036 |
| REACTOME:R-HSA-69242 | S Phase | 0.0751 | 0.1036 |
| REACTOME:R-HSA-5621481 | C-type lectin receptors (CLRs) | 0.0751 | 0.1036 |
| REACTOME:R-HSA-349425 | Autodegradation of the E3 ubiquitin ligase COP1 | 0.0751 | 0.1036 |
| REACTOME:R-HSA-450408 | AUF1 (hnRNP D0) binds and destabilizes mRNA | 0.0751 | 0.1036 |
| REACTOME:R-HSA-180534 | Vpu mediated degradation of CD4 | 0.0751 | 0.1036 |
| REACTOME:R-HSA-174113 | SCF-beta-TrCP mediated degradation of Emi1 | 0.0751 | 0.1036 |
| REACTOME:R-HSA-4641258 | Degradation of DVL | 0.0751 | 0.1036 |
| REACTOME:R-HSA-211733 | Regulation of activated PAK-2p34 by proteasome mediated degradation | 0.0751 | 0.1036 |
| REACTOME:R-HSA-5610785 | GLI3 is processed to GLI3R by the proteasome | 0.0751 | 0.1036 |
| REACTOME:R-HSA-8939236 | RUNX1 regulates transcription of genes involved in differentiation of HSCs | 0.0751 | 0.1036 |
| REACTOME:R-HSA-69601 | Ubiquitin Mediated Degradation of Phosphorylated Cdc25A | 0.0751 | 0.1036 |
| REACTOME:R-HSA-69610 | p53-Independent DNA Damage Response | 0.0751 | 0.1036 |
| REACTOME:R-HSA-69613 | p53-Independent G1/S DNA damage checkpoint | 0.0751 | 0.1036 |
| REACTOME:R-HSA-4641257 | Degradation of AXIN | 0.0751 | 0.1036 |
| REACTOME:R-HSA-69206 | G1/S Transition | 0.0751 | 0.1036 |
| REACTOME:R-HSA-68949 | Orc1 removal from chromatin | 0.0751 | 0.1036 |
| REACTOME:R-HSA-69052 | Switching of origins to a post-replicative state | 0.0751 | 0.1036 |
| REACTOME:R-HSA-8854050 | FBXL7 down-regulates AURKA during mitotic entry and in early mitosis | 0.0751 | 0.1036 |
| REACTOME:R-HSA-9020702 | Interleukin-1 signaling | 0.0751 | 0.1036 |
| REACTOME:R-HSA-5362768 | Hh mutants that don't undergo autocatalytic processing are degraded by ERAD | 0.0751 | 0.1036 |
| REACTOME:R-HSA-5610783 | Degradation of GLI2 by the proteasome | 0.0751 | 0.1036 |

| | | | |
|---|---|---|---|
| REACTOME:R-HSA-5627117 | RHO GTPases Activate ROCKs | 0.0751 | 0.1036 |
| REACTOME:R-HSA-69615 | G1/S DNA Damage Checkpoints | 0.0751 | 0.1036 |
| REACTOME:R-HSA-202403 | TCR signaling | 0.0751 | 0.1036 |
| REACTOME:R-HSA-400685 | Sema4D in semaphorin signaling | 0.0751 | 0.1036 |
| REACTOME:R-HSA-69300 | Removal of licensing factors from origins | 0.0751 | 0.1036 |
| REACTOME:R-HSA-69541 | Stabilization of p53 | 0.0751 | 0.1036 |
| REACTOME:R-HSA-68827 | CDT1 association with the CDC6:ORC:origin complex | 0.0751 | 0.1036 |
| REACTOME:R-HSA-8878159 | Transcriptional regulation by RUNX3 | 0.0751 | 0.1036 |
| REACTOME:R-HSA-5689603 | UCH proteinases | 0.0751 | 0.1036 |
| REACTOME:R-HSA-381426 | Regulation of Insulin-like Growth Factor (IGF) transport and uptake by Insulin-like Growth Factor Binding Proteins (IGFBPs) | 0.0751 | 0.1036 |
| REACTOME:R-HSA-453279 | Mitotic G1-G1/S phases | 0.0751 | 0.1036 |
| REACTOME:R-HSA-69017 | CDK-mediated phosphorylation and removal of Cdc6 | 0.0751 | 0.1036 |
| REACTOME:R-HSA-69481 | G2/M Checkpoints | 0.0751 | 0.1036 |
| REACTOME:R-HSA-5668541 | TNFR2 non-canonical NF-kB pathway | 0.0751 | 0.1036 |
| REACTOME:R-HSA-1234176 | Oxygen-dependent proline hydroxylation of Hypoxia-inducible Factor Alpha | 0.0751 | 0.1036 |
| REACTOME:R-HSA-69563 | p53-Dependent G1 DNA Damage Response | 0.0751 | 0.1036 |
| REACTOME:R-HSA-69580 | p53-Dependent G1/S DNA damage checkpoint | 0.0751 | 0.1036 |
| REACTOME:R-HSA-1236978 | Cross-presentation of soluble exogenous antigens (endosomes) | 0.0751 | 0.1036 |
| REACTOME:R-HSA-187577 | SCF(Skp2)-mediated degradation of p27/p21 | 0.0751 | 0.1036 |
| REACTOME:R-HSA-69229 | Ubiquitin-dependent degradation of Cyclin D1 | 0.0751 | 0.1036 |
| REACTOME:R-HSA-75815 | Ubiquitin-dependent degradation of Cyclin D | 0.0751 | 0.1036 |
| REACTOME:R-HSA-5610780 | Degradation of GLI1 by the proteasome | 0.0751 | 0.1036 |
| REACTOME:R-HSA-420029 | Tight junction interactions | 0.0751 | 0.1036 |
| REACTOME:R-HSA-169911 | Regulation of Apoptosis | 0.0751 | 0.1036 |
| REACTOME:R-HSA-68867 | Assembly of the pre-replicative complex | 0.0751 | 0.1036 |
| REACTOME:R-HSA-6807070 | PTEN Regulation | 0.0751 | 0.1036 |
| KEGG_PROTEASOME | Proteasome | 0.0751 | 0.1036 |
| KEGG_PRION_DISEASES | Prion diseases | 0.0751 | 0.1036 |
| REACTOME:R-HSA-69304 | Regulation of DNA replication | 0.0755 | 0.1042 |
| KEGG_PARKINSONS_DISEASE | Parkinson's disease | 0.0767 | 0.1058 |
| REACTOME:R-HSA-1236974 | ER-Phagosome pathway | 0.0788 | 0.1087 |
| REACTOME:R-HSA-936964 | Activation of IRF3/IRF7 mediated by TBK1/IKK epsilon | 0.0788 | 0.1087 |
| REACTOME:R-HSA-1630316 | Glycosaminoglycan metabolism | 0.0801 | 0.1105 |
| REACTOME:R-HSA-69656 | Cyclin A:Cdk2-associated events at S phase entry | 0.0801 | 0.1105 |
| REACTOME:R-HSA-1236975 | Antigen processing-Cross presentation | 0.0801 | 0.1105 |
| REACTOME:R-HSA-5658442 | Regulation of RAS by GAPs | 0.0801 | 0.1105 |
| REACTOME:R-HSA-4086400 | PCP/CE pathway | 0.0813 | 0.1122 |
| REACTOME:R-HSA-350562 | Regulation of ornithine decarboxylase (ODC) | 0.0813 | 0.1122 |
| REACTOME:R-HSA-202424 | Downstream TCR signaling | 0.0826 | 0.1139 |
| REACTOME:R-HSA-5678895 | Defective CFTR causes cystic fibrosis | 0.0826 | 0.1140 |
| REACTOME:R-HSA-8878171 | Transcriptional regulation by RUNX1 | 0.0833 | 0.1149 |
| REACTOME:R-HSA-8856828 | Clathrin-mediated endocytosis | 0.0833 | 0.1149 |
| REACTOME:R-HSA-5387390 | Hh mutants abrogate ligand secretion | 0.0867 | 0.1196 |
| REACTOME:R-HSA-69239 | Synthesis of DNA | 0.0867 | 0.1196 |
| REACTOME:R-HSA-5663205 | Infectious disease | 0.0870 | 0.1200 |

| | | | |
|---|---|---|---|
| REACTOME:R-HSA-416572 | Sema4D induced cell migration and growth-cone collapse | 0.0870 | 0.1200 |
| REACTOME:R-HSA-68874 | M/G1 Transition | 0.0872 | 0.1203 |
| REACTOME:R-HSA-69002 | DNA Replication Pre-Initiation | 0.0872 | 0.1203 |
| REACTOME:R-HSA-983705 | Signaling by the B Cell Receptor (BCR) | 0.0872 | 0.1203 |
| REACTOME:R-HSA-69202 | Cyclin E associated events during G1/S transition | 0.0872 | 0.1203 |
| KEGG_CARDIAC_MUSCLE_CONTRACTION | Cardiac muscle contraction | 0.0872 | 0.1203 |
| REACTOME:R-HSA-975577 | N-Glycan antennae elongation | 0.0873 | 0.1204 |
| REACTOME:R-HSA-5358346 | Hedgehog ligand biogenesis | 0.0873 | 0.1204 |
| REACTOME:R-HSA-162909 | Host Interactions of HIV factors | 0.0876 | 0.1208 |
| REACTOME:R-HSA-2470946 | Cohesin Loading onto Chromatin | 0.0928 | 0.1280 |
| REACTOME:R-HSA-5684264 | MAP3K8 (TPL2)-dependent MAPK1/3 activation | 0.0933 | 0.1286 |
| REACTOME:R-HSA-2871837 | FCERI mediated NF-kB activation | 0.0959 | 0.1323 |
| REACTOME:R-HSA-6804758 | Regulation of TP53 Activity through Acetylation | 0.0959 | 0.1323 |
| REACTOME:R-HSA-2559584 | Formation of Senescence-Associated Heterochromatin Foci (SAHF) | 0.0959 | 0.1323 |
| REACTOME:R-HSA-163200 | Respiratory electron transport, ATP synthesis by chemiosmotic coupling, and heat production by uncoupling proteins. | 0.0988 | 0.1363 |
| REACTOME:R-HSA-174084 | Autodegradation of Cdh1 by Cdh1:APC/C | 0.1050 | 0.1448 |
| REACTOME:R-HSA-75205 | Dissolution of Fibrin Clot | 0.1051 | 0.1450 |
| REACTOME:R-HSA-69620 | Cell Cycle Checkpoints | 0.1053 | 0.1453 |
| REACTOME:R-HSA-5687128 | MAPK6/MAPK4 signaling | 0.1053 | 0.1453 |
| REACTOME:R-HSA-3858494 | Beta-catenin independent WNT signaling | 0.1053 | 0.1453 |
| REACTOME:R-HSA-201681 | TCF dependent signaling in response to WNT | 0.1080 | 0.1490 |
| REACTOME:R-HSA-69306 | DNA Replication | 0.1089 | 0.1502 |
| REACTOME:R-HSA-376176 | Signaling by ROBO receptors | 0.1089 | 0.1502 |
| REACTOME:R-HSA-8963901 | Chylomicron remodeling | 0.1125 | 0.1552 |
| REACTOME:R-HSA-5689880 | Ub-specific processing proteases | 0.1147 | 0.1582 |
| REACTOME:R-HSA-2262749 | Cellular response to hypoxia | 0.1147 | 0.1582 |
| REACTOME:R-HSA-1234174 | Regulation of Hypoxia-inducible Factor (HIF) by oxygen | 0.1147 | 0.1582 |
| REACTOME:R-HSA-162906 | HIV Infection | 0.1182 | 0.1630 |
| REACTOME:R-HSA-446652 | Interleukin-1 family signaling | 0.1194 | 0.1647 |
| REACTOME:R-HSA-391903 | Eicosanoid ligand-binding receptors | 0.1218 | 0.1680 |
| REACTOME:R-HSA-611105 | Respiratory electron transport | 0.1242 | 0.1713 |
| REACTOME:R-HSA-1266738 | Developmental Biology | 0.1242 | 0.1713 |
| REACTOME:R-HSA-182971 | EGFR downregulation | 0.1286 | 0.1774 |
| REACTOME:R-HSA-8873719 | RAB geranylgeranylation | 0.1286 | 0.1774 |
| REACTOME:R-HSA-8878166 | Transcriptional regulation by RUNX2 | 0.1319 | 0.1819 |
| REACTOME:R-HSA-73893 | DNA Damage Bypass | 0.1319 | 0.1819 |
| KEGG_ALZHEIMERS_DISEASE | Alzheimer's disease | 0.1319 | 0.1819 |
| REACTOME:R-HSA-174154 | APC/C:Cdc20 mediated degradation of Securin | 0.1377 | 0.1899 |
| REACTOME:R-HSA-68886 | M Phase | 0.1377 | 0.1899 |
| REACTOME:R-HSA-399956 | CRMPs in Sema3A signaling | 0.1377 | 0.1899 |
| REACTOME:R-HSA-177929 | Signaling by EGFR | 0.1423 | 0.1963 |

**Supplmentary Table S10. Functional analysis of the differentially expressed genes in mouse models**

| Gene | Allelic Composition | Consequence of mutant allele | Genetic Background | Genotype ID | Qualifier | Annotated Term | Phenotype Summary Category | Reference |
|---|---|---|---|---|---|---|---|---|
| Ppp1r3b | | | | | | | | |
| | Ppp1r3b<tm1.1Arte>/Ppp1r3b<+> | Ppp1r3b<tm1.1Arte>: Y284F | involves: C57BL/6J * C57BL/6N | MGI:4431169 | | decreased circulating triglyceride level | homeostasis/metabolism | J:157291 |
| | Ppp1r3b<tm1.1Arte>/Ppp1r3b<+> | Ppp1r3b<tm1.1Arte>: Y284F | involves: C57BL/6J * C57BL/6N | MGI:4431169 | | improved glucose tolerance | homeostasis/metabolism | J:157291 |
| | Ppp1r3b<tm1.1Arte>/Ppp1r3b<tm1.1Arte> | Ppp1r3b<tm1.1Arte>: Y284F | involves: C57BL/6J * C57BL/6N | MGI:4431168 | | decreased body weight | growth/size/body region | J:157291 |
| | Ppp1r3b<tm1.1Arte>/Ppp1r3b<tm1.1Arte> | Ppp1r3b<tm1.1Arte>: Y284F | involves: C57BL/6J * C57BL/6N | MGI:4431168 | | decreased circulating glucose level | homeostasis/metabolism | J:157291 |
| | Ppp1r3b<tm1.1Arte>/Ppp1r3b<tm1.1Arte> | Ppp1r3b<tm1.1Arte>: Y284F | involves: C57BL/6J * C57BL/6N | MGI:4431168 | | improved glucose tolerance | homeostasis/metabolism | J:157291 |
| | Ppp1r3b<tm1.1Arte>/Ppp1r3b<tm1.1Arte> | Ppp1r3b<tm1.1Arte>: Y284F | involves: C57BL/6J * C57BL/6N | MGI:4431168 | | lethargy | behavior/neurological | J:157291 |
| | Ppp1r3b<tm1.1Arte>/Ppp1r3b<tm1.1Arte> | Ppp1r3b<tm1.1Arte>: Y284F | involves: C57BL/6J * C57BL/6N | MGI:4431168 | | transmission ratio distortion | reproductive system | J:157291 |
| | Ppp1r3b<tm1.1Arte>/Ppp1r3b<tm1.1Arte> | Ppp1r3b<tm1.1Arte>: Y284F | involves: C57BL/6N | MGI:5568694 | normal | homeostasis/metabolism phenotype | homeostasis/metabolism | J:208923 |
| Tap2 | | | | | | | | |
| | Tap2<jas>/Tap2<jas> | Tap2<jas>: T293P | C57BL/6JSfdAnu-Tap2<jas>/AnuApb | MGI:3612047 | | absent CD8-positive, alpha-beta T cells | hematopoietic system | J:104190 |
| | Tap2<jas>/Tap2<jas> | Tap2<jas>: T293P | C57BL/6JSfdAnu-Tap2<jas>/AnuApb | MGI:3612047 | | absent CD8-positive, alpha-beta T cells | immune system | J:104190 |
| | Tap2<jas>/Tap2<jas> | Tap2<jas>: T293P | C57BL/6JSfdAnu-Tap2<jas>/AnuApb | MGI:3612047 | | decreased CD8-positive, alpha-beta T cell number | hematopoietic system | J:166104 |
| | Tap2<jas>/Tap2<jas> | Tap2<jas>: T293P | C57BL/6JSfdAnu-Tap2<jas>/AnuApb | MGI:3612047 | | decreased CD8-positive, alpha-beta T cell number | immune system | J:166104 |
| | Tap2<m1Btlr>/Tap2<m1Btlr> | Tap2<m1Btlr>: frameshift, premature stop codon | C57BL/6J-Tap2<m1Btlr> | MGI:4442847 | | abnormal cytotoxic T cell physiology | hematopoietic system | J:159355 |
| | Tap2<m1Btlr>/Tap2<m1Btlr> | Tap2<m1Btlr>: frameshift, premature stop codon | C57BL/6J-Tap2<m1Btlr> | MGI:4442847 | | abnormal cytotoxic T cell physiology | immune system | J:159355 |
| | Tap2<m1Btlr>/Tap2<m1Btlr> | Tap2<m1Btlr>: frameshift, premature stop codon | C57BL/6J-Tap2<m1Btlr> | MGI:4442847 | | abnormal NK cell physiology | hematopoietic system | J:159355 |
| | Tap2<m1Btlr>/Tap2<m1Btlr> | Tap2<m1Btlr>: frameshift, premature stop codon | C57BL/6J-Tap2<m1Btlr> | MGI:4442847 | | abnormal NK cell physiology | immune system | J:159355 |
| | Tap2<m1Btlr>/Tap2<m1Btlr> | Tap2<m1Btlr>: frameshift, premature stop codon | C57BL/6J-Tap2<m1Btlr> | MGI:4442847 | | absent CD8-positive, alpha-beta T cells | hematopoietic system | J:159355 |
| | Tap2<m1Btlr>/Tap2<m1Btlr> | Tap2<m1Btlr>: frameshift, premature stop codon | C57BL/6J-Tap2<m1Btlr> | MGI:4442847 | | absent CD8-positive, alpha-beta T cells | immune system | J:159355 |
| | Tap2<m1Btlr>/Tap2<m1Btlr> | Tap2<m1Btlr>: frameshift, premature stop codon | C57BL/6J-Tap2<m1Btlr> | MGI:4442847 | | decreased level of surface class I molecules | immune system | J:159355 |
| | Tap2<m1Btlr>/Tap2<m1Btlr> | Tap2<m1Btlr>: frameshift, premature stop codon | C57BL/6J-Tap2<m1Btlr> | MGI:4442847 | | defective assembly of class I molecules | immune system | J:159355 |
| | Tap2<m2Btlr>/Tap2<m2Btlr> | Tap2<m2Btlr>: V422D, loss of function predicted by Polyphen-2 | C57BL/6J-Tap2<m2Btlr> | MGI:5613177 | | decreased CD8-positive, alpha-beta T cell number | hematopoietic system | J:217145 |
| | Tap2<m2Btlr>/Tap2<m2Btlr> | Tap2<m2Btlr>: V422D, loss of function predicted by Polyphen-2 | C57BL/6J-Tap2<m2Btlr> | MGI:5613177 | | decreased CD8-positive, alpha-beta T cell number | immune system | J:217145 |
| | Tap2<m2Btlr>/Tap2<m2Btlr> | Tap2<m2Btlr>: V422D, loss of function predicted by Polyphen-2 | C57BL/6J-Tap2<m2Btlr> | MGI:5613177 | | increased CD4-positive, alpha-beta T cell number | hematopoietic system | J:217145 |
| | Tap2<m2Btlr>/Tap2<m2Btlr> | Tap2<m2Btlr>: V422D, loss of function predicted by Polyphen-2 | C57BL/6J-Tap2<m2Btlr> | MGI:5613177 | | increased CD4-positive, alpha-beta T cell number | immune system | J:217145 |
| | Tap2<m3Btlr>/Tap2<m3Btlr> | Tap2<m3Btlr>: Y429* (premature stop codon) | C57BL/6J-Tap2<m3Btlr> | MGI:6113683 | | decreased CD8-positive, alpha-beta T cell number | hematopoietic system | J:254929 |
| | Tap2<m3Btlr>/Tap2<m3Btlr> | Tap2<m3Btlr>: Y429* (premature stop codon) | C57BL/6J-Tap2<m3Btlr> | MGI:6113683 | | decreased CD8-positive, alpha-beta T cell number | immune system | J:254929 |
| | Tap2<m3Btlr>/Tap2<m3Btlr> | Tap2<m3Btlr>: Y429* (premature stop codon) | C57BL/6J-Tap2<m3Btlr> | MGI:6113683 | | decreased CD8-positive, naive alpha-beta T cell number | hematopoietic system | J:254929 |
| | Tap2<m3Btlr>/Tap2<m3Btlr> | Tap2<m3Btlr>: Y429* (premature stop codon) | C57BL/6J-Tap2<m3Btlr> | MGI:6113683 | | decreased CD8-positive, naive alpha-beta T cell number | immune system | J:254929 |
| | Tap2<m3Btlr>/Tap2<m3Btlr> | Tap2<m3Btlr>: Y429* (premature stop codon) | C57BL/6J-Tap2<m3Btlr> | MGI:6113683 | | decreased effector memory CD4-positive, alpha-beta T cell number | hematopoietic system | J:254929 |
| | Tap2<m3Btlr>/Tap2<m3Btlr> | Tap2<m3Btlr>: Y429* (premature stop codon) | C57BL/6J-Tap2<m3Btlr> | MGI:6113683 | | decreased effector memory CD4-positive, alpha-beta T cell number | immune system | J:254929 |
| | Tap2<m3Btlr>/Tap2<m3Btlr> | Tap2<m3Btlr>: Y429* (premature stop codon) | C57BL/6J-Tap2<m3Btlr> | MGI:6113683 | | decreased NK T cell number | hematopoietic system | J:254929 |
| | Tap2<m3Btlr>/Tap2<m3Btlr> | Tap2<m3Btlr>: Y429* (premature stop codon) | C57BL/6J-Tap2<m3Btlr> | MGI:6113683 | | decreased NK T cell number | immune system | J:254929 |
| | Tap2<m3Btlr>/Tap2<m3Btlr> | Tap2<m3Btlr>: Y429* (premature stop codon) | C57BL/6J-Tap2<m3Btlr> | MGI:6113683 | | increased CD4-positive, alpha-beta T cell number | hematopoietic system | J:254929 |
| | Tap2<m3Btlr>/Tap2<m3Btlr> | Tap2<m3Btlr>: Y429* (premature stop codon) | C57BL/6J-Tap2<m3Btlr> | MGI:6113683 | | increased CD4-positive, alpha-beta T cell number | immune system | J:254929 |
| | Tap2<m3Btlr>/Tap2<m3Btlr> | Tap2<m3Btlr>: Y429* (premature stop codon) | C57BL/6J-Tap2<m3Btlr> | MGI:6113683 | | increased central memory CD8 positive, alpha-beta T cell number | hematopoietic system | J:254929 |
| | Tap2<m3Btlr>/Tap2<m3Btlr> | Tap2<m3Btlr>: Y429* (premature stop codon) | C57BL/6J-Tap2<m3Btlr> | MGI:6113683 | | increased central memory CD8 positive, alpha-beta T cell number | immune system | J:254929 |
| | Tap2<M4Btlr>/Tap2<M4Btlr> | Tap2<m3Btlr>: Y429* (premature stop codon) | C57BL/6J-Tap2<M4Btlr> | MGI:6286084 | | increased CD4-positive, alpha-beta T cell number | hematopoietic system | J:272847 |
| | Tap2<M4Btlr>/Tap2<M4Btlr> | Tap2<m3Btlr>: Y429* (premature stop codon) | C57BL/6J-Tap2<M4Btlr> | MGI:6286084 | | increased CD4-positive, alpha-beta T cell number | immune system | J:272847 |
| Oasl1 | | | | | | | | |
| | Oasl1<tm1b(EUCOMM)Hmgu>/Oasl1<tm1b( | Oasl1<tm1b(EUCOMM)Hmgu>: Null/knockout | C57BL/6N-Oasl1<tm1b(EUCOMM)Hmgu>/BayMmucd | MGI:6461494 | | preweaning lethality, incomplete penetrance[1] | mortality/aging | J:211773 |
| | Oasl1<tm1Yjk>/Oasl1<tm1Yjk> | Oasl1<tm1Yjk>: Null/knockout | B6.129P2-Oasl1<tm1Yjk> | MGI:5498951 | | abnormal circulating interferon level | homeostasis/metabolism | J:194467 |
| | Oasl1<tm1Yjk>/Oasl1<tm1Yjk> | Oasl1<tm1Yjk>: Null/knockout | B6.129P2-Oasl1<tm1Yjk> | MGI:5498951 | | abnormal circulating interferon level | immune system | J:194467 |
| | Oasl1<tm1Yjk>/Oasl1<tm1Yjk> | Oasl1<tm1Yjk>: Null/knockout | B6.129P2-Oasl1<tm1Yjk> | MGI:5498951 | | decreased susceptibility to Picornaviridae infection | immune system | J:194467 |
| | Oasl1<tm1Yjk>/Oasl1<tm1Yjk> | Oasl1<tm1Yjk>: Null/knockout | B6.129P2-Oasl1<tm1Yjk> | MGI:5498951 | | decreased susceptibility to Picornaviridae infection induced morbidity/mortality | immune system | J:194467 |
| | Oasl1<tm1Yjk>/Oasl1<tm1Yjk> | Oasl1<tm1Yjk>: Null/knockout | B6.129P2-Oasl1<tm1Yjk> | MGI:5498951 | | decreased susceptibility to Picornaviridae infection induced morbidity/mortality | mortality/aging | J:194467 |
| Oasl2 | | | | | | | | |
| | Oasl2<tm1a(EUCOMM)Wtsi>/Oasl2<tm1a(E | Oasl2<tm1a(EUCOMM)Wtsi>: Null/knockout | C57BL/6N-Oasl2<tm1a(EUCOMM)Wtsi>/Wtsi | MGI:5781689 | | decreased circulating glucose level[1] | homeostasis/metabolism | J:175295 |
| Ube2e2 | | | | | | | | |
| | Ube2e2<tm1b(KOMP)Wtsi>/Ube2e2<tm1b(K | Ube2e2<tm1b(KOMP)Wtsi>: Null/knockout | C57BL/6N-Ube2e2<tm1b(KOMP)Wtsi>/Ucd | MGI:6263809 | | increased mean corpuscular volume[1] | hematopoietic system | J:211773 |

[1]Phenotypes found in the International Mouse Phenotyping Consortium (IMPC)
