## Supplementary Materials for "Elucidating the Epigenetic Landscape of Type 2 Diabetes: A Multi-Omics Analysis Revealing Novel CpG Sites and Their Association with Cardiometabolic Traits"

### Methods

Overview of the PrediXcan algorithm

PrediXcan is a computational method designed to understand how genetic variations influence phenotypes by exploring the underlying mechanism of gene regulation [1]. In the study of PrediXcan, an individual's gene expression level is categorized into three components: genetically regulated expression (GReX), reflecting the inherent genetic contribution; the component altered by traits, such as diseases modifying expression levels; and components attributed to environmental factors. PrediXcan's primary goal is to determine the extent to which GReX impacts the trait or phenotype under examination. Using reference transcriptome datasets from studies like GTEx [2], GEUVADIS [3], and DGN [4], it trains additive models to estimate these genetically regulated expressions. These models subsequently allow for the prediction of gene expression levels based on multiple SNPs, and these predictions are then tested for associations with actual phenotypic data. PrediXcan uses a mechanism-centric approach, meaning that it centers on the mechanism of gene expression regulation, which is pivotal in many diseases. It also reduces the testing burden by testing around 10,000 to 20,000 genes instead of millions of SNP tests in GWAS. Most importantly, as the method revolves around predicted expression levels determined by genetic variations, there's no need for actual transcriptome data. This makes PrediXcan compatible with large-scale genome databases like dbGaP.

Overview of the S-PrediXcan algorithm

S-PrediXcan [5] is an advancement of the original PrediXcan method, developed to infer the results of PrediXcan utilizing only GWAS summary statistics, eliminating the need for individual-level data. This approach employs an analytical expression devised to compute the outcome of PrediXcan based solely on the mentioned statistics. Inputs for S-PrediXcan include weights for predicting a specific gene's expression, variance and covariances of the SNPs included in the prediction, as well as the GWAS summary statistics for each SNP. The weights for gene expression prediction, as those in PrediXcan, can be calculated from resources such as GTEx and DGN. Additionally, the variance and covariances of the SNPs can be calculated from a reference dataset such as the 1000 Genomes Project. Based on this information, a formula was derived to infer the PrediXcan results. Notably, a high concordance between PrediXcan and S-PrediXcan results has been observed. Using S-PrediXcan does not compromise the power to detect associations compared to using PrediXcan.

Overview of the Methylation Prediction Model:

DNA methylation is influenced by specific SNPs, with numerous methylation quantitative trait loci (meQTL) identified as SNPs having strong associations with methylation levels at particular CpG sites [6]. In a study by Fryett et al. [7], the potential to predict methylation levels using meQTLs was assessed by developing methylation prediction models through machine-learning algorithms. These models were trained using the Understanding Society data derived from the UK Household Longitudinal Study [8], which provided genotype and CpG methylation data for 1,120 participants post-quality control. For the creation of these models, methylation levels were regressed against the genotype dosages of SNPs situated near the CpG sites. The study employed three penalized regression techniques: ridge regression, LASSO, and the elastic net. Through the application of cross-validation and diverse testing methods, like adjusting the SNP window sizes, the models were refined. They observed that both LASSO and the elastic net typically outperformed ridge regression in prediction accuracy. However, a significant finding was that a majority of CpG methylation levels proved challenging to predict, with only about 200,000 out of 800,000 CpGs achieving a prediction accuracy of 0.1. Ultimately, the model demonstrating the highest predictive accuracy, considering the optimal method and window size, was selected.

The UK Biobank cohort

The UK Biobank is a comprehensive prospective study that initially enrolled over 500,000 individuals aged 40 to 69 years between 2006 and 2010. It systematically gathers a wide array of data on its participants, encompassing phenotypic and genotypic information, responses from questionnaires, physical measurements, sample analyses, activity tracking, diverse imaging techniques, genome-wide genotyping, and ongoing monitoring for various health outcomes [9]. Adhering to the quality control (QC) methods detailed by Bycroft et al. [10], we excluded samples that were outliers in terms of heterozygosity and missing rates (data field 22027), those putatively carrying sex chromosome configurations not classified as XX or XY (data field 22019), and focused only on unrelated individuals (data field 22020). Our analysis was particularly concentrated on the white British demographic (data field 22006), narrowing down the sample size to 339,455 for our study. The imputed genotypes, with details provided by Bycroft et al. [10], were used. Only genotypes with imputation quality scores > 0.3 were considered in our study. We defined prevalent type 2 diabetes (T2D) as individuals who, at the time of enrollment, reported a doctor's diagnosis of diabetes (data field 2443) or had diabetes diagnosis codes in their General Practitioner (GP) or hospital records up to three years before the recruitment baseline. These diagnosis codes were primarily based on the ICD-9 classification, specifically the initial three digits of 250. This approach led to the identification of 25,025 individuals with prevalent T2D at baseline and 314,430 controls for the analysis.

The Taiwan Biobank cohort

Since its inception in 2008, the Taiwan Biobank has collected samples over 150,000 individuals aged 20 and above [11]. Of these, around 129,000 individuals were genotyped using the TWB1.0 and TWB2.0 GWAS SNP arrays [12]. TWB1.0 chips, based on the customized Axiom Genome-Wide Array Plate system, comprise approximately 653,000 SNPs. TWB2.0 chips expanded upon the TWB1.0 design, encompassing about 750,000 SNPs. We adhered to the standard GWAS QC procedures, including the elimination of SNPs with Hardy-Weinberg Equilibrium p-values < $10^{-4}$, the removal of duplicate samples and closely related individuals, and the exclusion of participants with a mismatch between self-reported gender and genetic data-estimated biological sex. Post-QC samples were imputed via the Michigan Imputation Server, utilizing the 1000 Genomes Project Phase 3 data as a reference. Subsequent analyses were based on a total of 7,369,045 SNPs, each with an imputation Rsq > 0.3 and a minor allele frequency (MAF) > 1%.

In the Taiwan Biobank, approximately 2,300 individuals with GWAS data underwent genome-wide methylation profiling using the Illumina Infinium MethylationEPIC BeadChip (referred to as TWB EPIC). After standard QC procedures [13], 2,150 unrelated individuals and 761,617 CpGs remained. Briefly, the QC procedures was conducted using the “bigmelon” R package [14], which involved removing outlier samples via principal component analysis, excluding samples with low bisulfite conversion rates, and normalizing the data with the "dasen" function. Further QC steps included discarding samples with significant post-normalization changes and those with a high proportion of undetected CpG sites. After a second normalization, outlier CpG values were set to missing, and any samples or CpGs with excessive missing data were removed. Finally, probes affected by common genetic variations, cross-hybridization issues, or located on sex chromosomes were removed.

The methylation quantitative trait loci (meQTL) mapping was conducted using the software tensorQTL [15]. This tool, which utilizes GPU acceleration, significantly enhances the speed of both cis- and trans-QTL mapping—by roughly 200 to 300 times—when compared to CPU-based approaches. Our analysis focused on cis-meQTL mapping for the 87 independent and significant CpG sites identified in our European methylome-wide association study (MWAS), using samples from the Taiwan Biobank. For each CpG site, SNPs located within 1 MB were examined for their association with DNA methylation levels at these sites, employing a linear regression model that adjusted for age, sex, and smoking status. These SNPs were further refined using PLINK's --clump option [16], based on their association p-values, to ensure that the linkage disequilibrium (LD) $r^2$ between retained SNPs was less than 0.5. This process yielded 800,981 SNP-CpG pairs. SNPs that had association p-values less than $6.24 \times 10^{-8}$—to adjust for the multiple tests performed across the 800,981 pairs—were identified as meQTLs for the 87 CpG sites. In total, 19,527 meQTL were discovered, and their association statistics were subsequently utilized in Mendelian Randomization (MR) analyses focused on the East Asian population.

The PANC-DB dataset

PANC-DB serves as an open-source repository designed for the storage, dissemination, and visualization of cellular and molecular data produced by the Human Pancreas Analysis Program (HPAP), thereby enhancing the accessibility of genomic and islet function data for the diabetes research community [17]. It integrates three main facets: data management and access, computational biology and data science, and communication and outreach efforts. HPAP, a key component of the Human Islet Research Network (HIRN), focuses on the detailed phenotyping of the human endocrine pancreas to uncover the processes leading to beta-cell loss or dysfunction in both type 1 and type 2 diabetes. We acquired DNAm data, derived from whole-genome bisulfite sequencing (WGBS), and gene expression data, obtained through RNA sequencing (RNA-Seq) of pancreatic alpha and beta cells, from PANC-DB. Analysis of the DNAm data revealed DNAm levels at 82 of the 87 CpGs identified in our MWAS, which were subsequently analyzed.

For the RNA-Seq data, we adhered to the mRNA Analysis Pipeline (https://docs.gdc.cancer.gov/Data/Bioinformatics_Pipelines/Expression_mRNA_Pipeline/) from the National Cancer Institute Genomic Data Commons (GDC) . This pipeline begins with an Alignment Workflow that uses the STAR tool to implement a two-pass alignment method. Each read group is aligned separately by STAR, then merged to produce a comprehensive alignment, incorporating a splice junction detection step as recommended by the International Cancer Genome Consortium (ICGC). The result is a genomic BAM file containing both aligned and unaligned reads. Quality control is rigorously applied before alignment with FASTQC and after alignment with Picard Tools. Finally, featureCounts [18] is utilized to generate raw counts, from which transcripts per million (TPM) values are calculated for gene expression analysis.

In our study, we conducted a differential gene expression analysis by comparing the mean TPM values between T2D patients and control individuals without a history of diabetes. We first adjusted the TPM for each gene by age and sex using a linear regression model, then derived residuals for further analysis. Given the limited size of the sample, we employed a permutation test to assess the association between these residuals and T2D status. Specifically, we calculated the difference in mean TPM values between the T2D and control groups. In the permutation test, we randomly shuffled the diabetes status among the samples and recalculated the mean TPM difference for each permutation. This process was repeated

10,000 times. The permutation p-value was determined by the fraction of permutations where the absolute mean difference was greater than the absolute observed mean difference.

### Figures

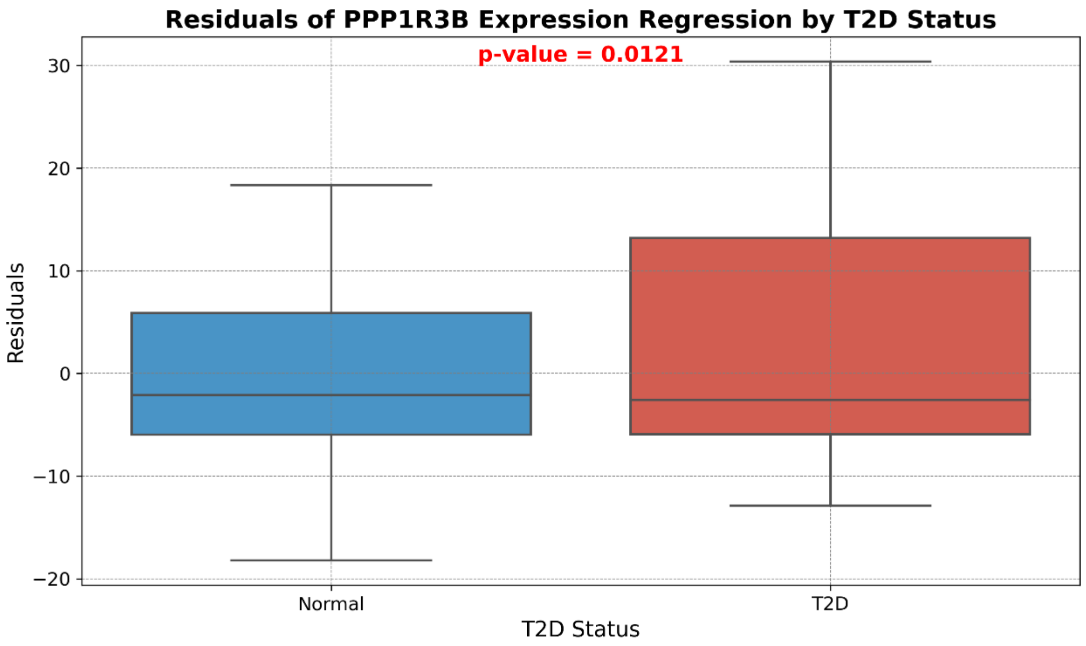


Figure S1. Box plot illustrating the residuals of PPP1R3B gene expression, adjusted for age and sex, in pancreatic alpha cells across the normal and T2D groups. The displayed p-value signifies the difference in means between the two groups, determined through a permutation test.

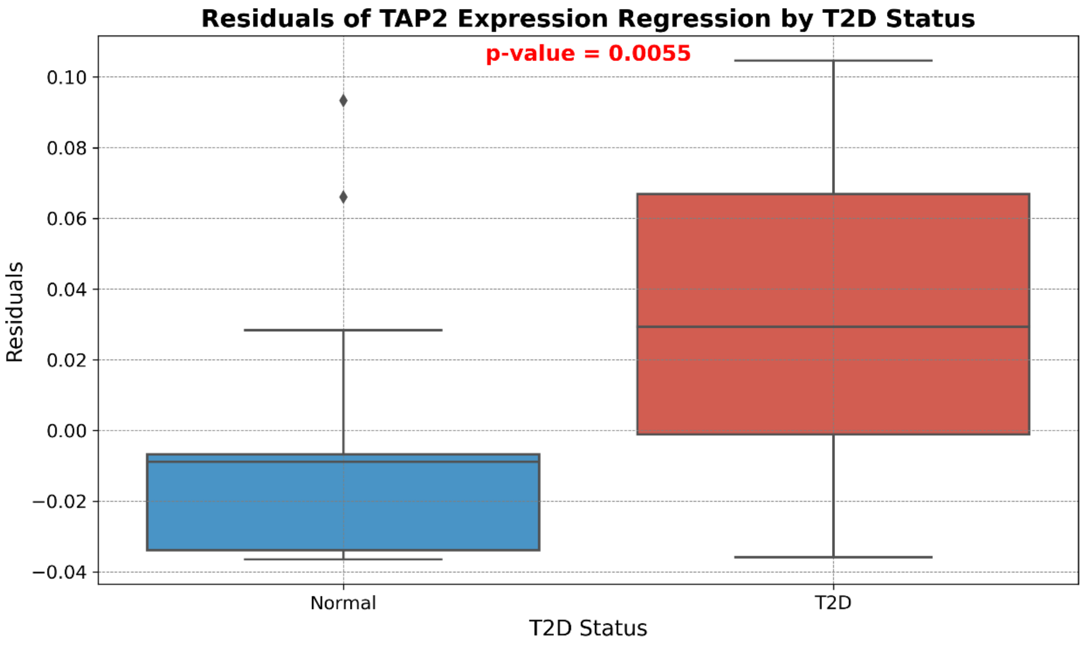


Figure S2. Box plot illustrating the residuals of TAP2 gene expression, adjusted for age and sex, in pancreatic alpha cells across the normal and T2D groups. The displayed p-value signifies the difference in means between the two groups, determined through a permutation test.

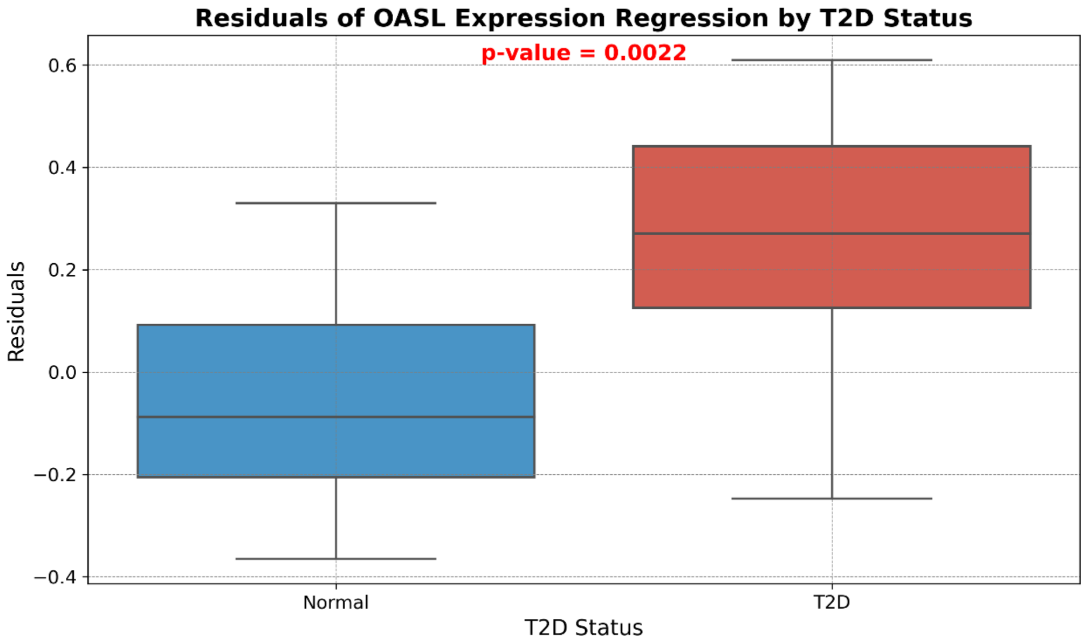


Figure S3. Box plot illustrating the residuals of OASL gene expression, adjusted for age and sex, in pancreatic alpha cells across the normal and T2D groups. The displayed p-value signifies the difference in means between the two groups, determined through a permutation test.

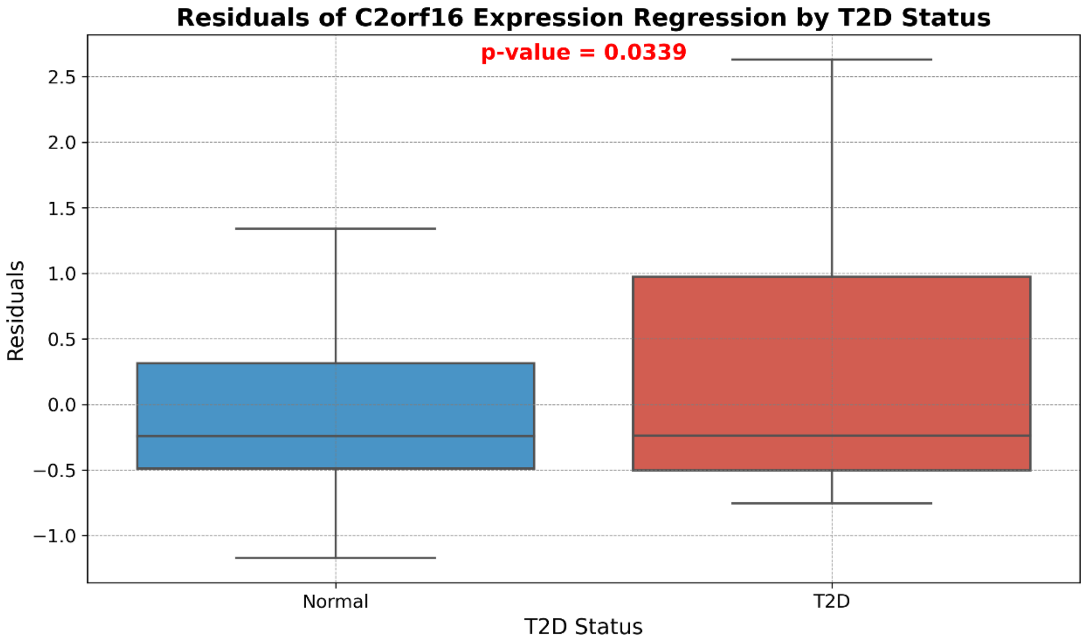


Figure S4. Box plot illustrating the residuals of C2orf16 gene expression, adjusted for age and sex, in pancreatic beta cells across the normal and T2D groups. The displayed p-value signifies the difference in means between the two groups, determined through a permutation test.

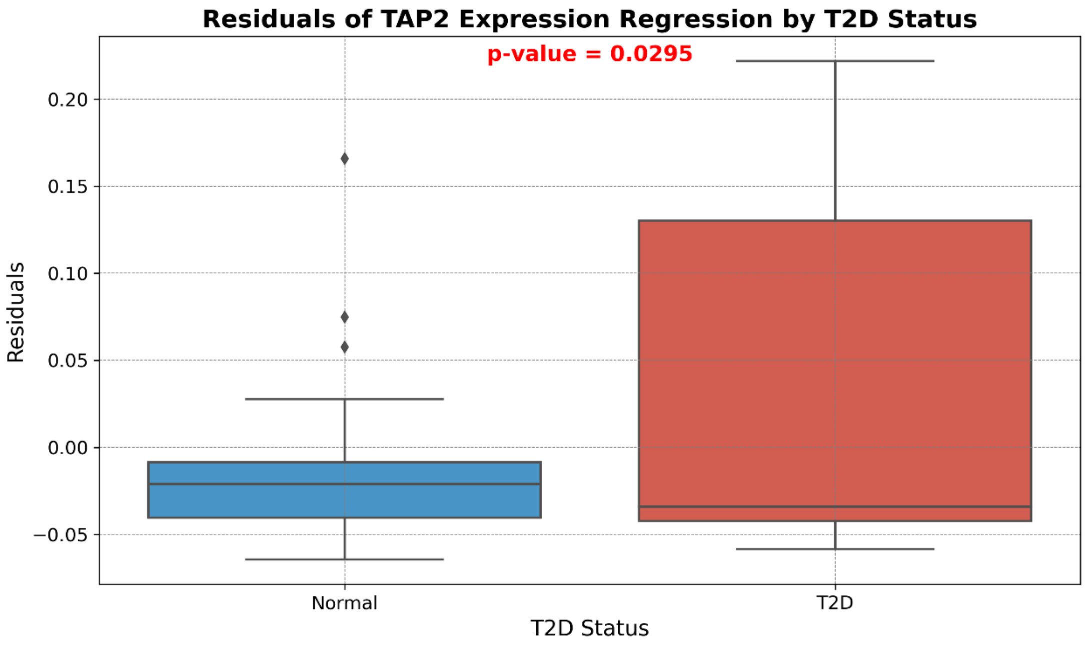


Figure S5. Box plot illustrating the residuals of TAP2 gene expression, adjusted for age and sex, in pancreatic beta cells across the normal and T2D groups. The displayed p-value signifies the difference in means between the two groups, determined through a permutation test.

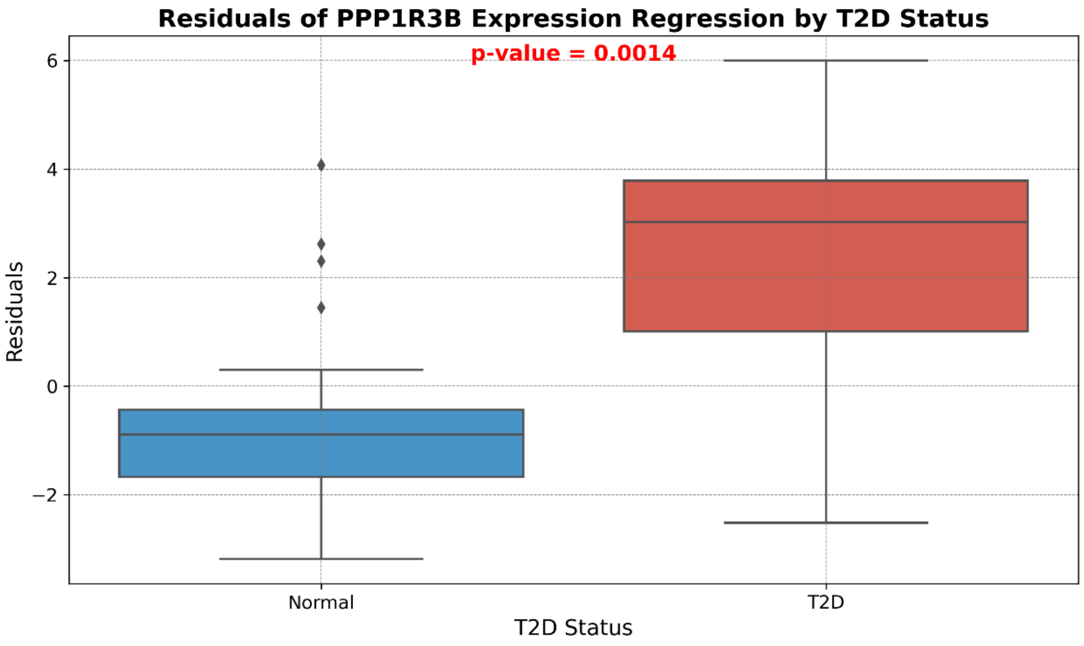


Figure S6. Box plot illustrating the residuals of PPP1R3B gene expression, adjusted for age and sex, in pancreatic beta cells across the normal and T2D groups. The displayed p-value signifies the difference in means between the two groups, determined through a permutation test.

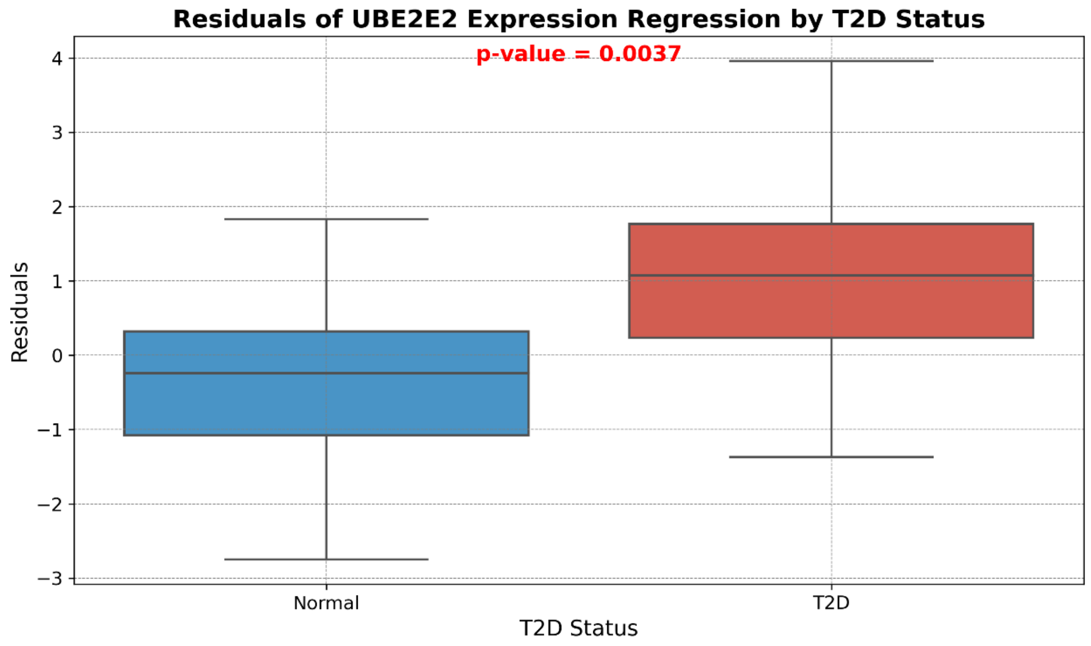


Figure S7. Box plot illustrating the residuals of UBE2E2 gene expression, adjusted for age and sex, in pancreatic beta cells across the normal and T2D groups. The displayed p-value signifies the difference in means between the two groups, determined through a permutation test.
